# Improved multi-ancestry fine-mapping identifies *cis*-regulatory variants underlying molecular traits and disease risk

**DOI:** 10.1101/2024.04.15.24305836

**Authors:** Zeyun Lu, Xinran Wang, Matthew Carr, Artem Kim, Steven Gazal, Pejman Mohammadi, Lang Wu, Alexander Gusev, James Pirruccello, Linda Kachuri, Nicholas Mancuso

## Abstract

Multi-ancestry statistical fine-mapping of *cis*-molecular quantitative trait loci (*cis*-molQTL) aims to improve the precision of distinguishing causal *cis*-molQTLs from tagging variants. However, existing approaches fail to reflect shared genetic architectures. To solve this limitation, we present the Sum of Shared Single Effects (SuShiE) model, which leverages LD heterogeneity to improve fine-mapping precision, infer cross-ancestry effect size correlations, and estimate ancestry-specific expression prediction weights. We apply SuShiE to mRNA expression measured in PBMCs (n=956) and LCLs (n=814) together with plasma protein levels (n=854) from individuals of diverse ancestries in the TOPMed MESA and GENOA studies. We find SuShiE fine-maps *cis*-molQTLs for 16*%* more genes compared with baselines while prioritizing fewer variants with greater functional enrichment. SuShiE infers highly consistent *cis*-molQTL architectures across ancestries on average; however, we also find evidence of heterogeneity at genes with predicted loss-of-function intolerance, suggesting that environmental interactions may partially explain differences in *cis*-molQTL effect sizes across ancestries. Lastly, we leverage estimated *cis*-molQTL effect-sizes to perform individual-level TWAS and PWAS on six white blood cell-related traits in AOU Biobank individuals (n=86k), and identify 44 more genes compared with baselines, further highlighting its benefits in identifying genes relevant for complex disease risk. Overall, SuShiE provides new insights into the *cis*-genetic architecture of molecular traits.

## Introduction

Characterizing the functional consequences of genetic variation remains a crucial task in deciphering the mechanisms underlying complex disease risk^1,2^. To this end, *cis*-molecular quantitative trait loci (*cis*-molQTL) mapping seeks to identify genetic variants associated with genomically proximal molecular features measured across diverse cellular, tissue, and environmental contexts^3–14^. However, due to linkage disequilibrium (LD), it is challenging to distinguish causal *cis*-molQTLs from tagging variants within a genomic region^3,5^. Statistical fine-mapping aims to resolve precisely this issue^15–19^, yet pervasive LD signals limit the resolution of these approaches. Previous efforts have demonstrated that leveraging the heterogeneity of LD and minor allele frequency (MAF) across diverse ancestries improves the precision of statistical fine-mapping and therefore enhances our biological understanding of complex diseases^20–25^ and molecular traits^26–32^.

While existing multi-ancestry fine-mapping frameworks have been proposed for the analysis of complex traits and diseases^30,33–41^, they have several limitations in the context of large-scale *cis*-molQTL data. First, many approaches do not model the correlation of causal variant effect sizes across ancestries or assume that they are a-priori independent across ancestries, which fails to reflect shared or similar genetic architectures^33,35,37,38^. Second, existing multi-ancestry approaches scale poorly, which precludes their application to thousands of molecular traits commonly measured in *cis*-molQTL studies^33,35,40^. Third, current fine-mapping approaches lack ancestry-specific effect size estimates^33,35,37^, which neglects their potential use in post-Genome-wide Association Studies (GWASs) frameworks (e.g., Transcriptome- and Proteome-wide Association Studies (TWASs/PWASs)^42–47^. Last, while recent approaches address some of these limitations, existing software implementations are capable of analyzing only two ancestries, which excludes datasets consisting of ever-increasing diverse ancestries^39^.

Here, we describe the Sum of Shared Single Effects (SuShiE) approach to fine-map genetic variants shared across diverse ancestries for thousands of molecular traits. SuShiE integrates genotypic and molecular data from multiple ancestries to identify *cis*-molQTLs while modeling and learning the covariance structures of shared/non-shared signals. SuShiE leverages four key insights. First, SuShiE improves fine-mapping precision of the shared *cis*-molQTLs by leveraging LD across different ancestries. Second, it estimates ancestry-specific effect sizes at shared *cis*-molQTLs. Third, it infers the prior effect size correlation across ancestries to shed light on genetic similarities and differences. Lastly, SuShiE is implemented using a scalable variational inference algorithm that runs seamlessly on CPUs, GPUs, or tensor-processing units (TPUs).

Through extensive simulations, we show that SuShiE outputs higher posterior inclusion probabilities (PIPs) at causal *cis*-molQTLs, outputs smaller credible set sizes, and exhibits better calibration compared with current approaches^15,38^. Using bulk mRNA expression levels measured in peripheral blood mononuclear cells (PBMCs) and lymphoblastoid cell lines (LCLs) together with protein abundance measured in plasma, we fine-map 36,911 molecular phenotypes across American European, African, and Hispanic ancestries from TOPMed-MESA^48,49^ (n_mRNA_=956 and n_protein_=814) and GENOA^26^ (n_mRNA_=854). SuShiE fine-maps significantly more *cis*-molQTLs with smaller credible sets and greater enrichment in relevant functional annotations compared with existing methods. In addition, SuShiE infers shared genetic architecture of *cis*-molQTL in significantly heritable genes and shows the heterogeneity across ancestries of signals associated with multiple measures of loss-of-function (LOF) intolerance. Last, we integrate ancestry-specific *cis*-molQTL effects inferred by SuShiE with six white blood cell-related traits to perform individual-level TWAS and PWAS in the All of Us Biobank (average n=86,345)^50^ and observe that SuShiE-based prediction models identified 44 additional associated genes compared with the baseline approach. Overall, our approach sheds light on understanding the genetic *cis*-architecture of molecular data across multiple ancestries.

## Results

### SuShiE overview

Here, we briefly introduce the SuShiE model (for a detailed description, see **Methods** and **Supplementary Note**). SuShiE assumes *cis*-molQTLs are present in *all* ancestries (defined as shared *cis*-molQTLs) while allowing for effect sizes at causal *cis*-molQTLs to covary across ancestries a-priori, in contrast to previous multi-ancestry approaches^15,33,35,37,38^. These assumptions provide enough flexibility to model a variety of *cis*-genetic architectures across ancestries, including cases when effects are present only in a subset of ancestries. For instance, when effects are observed only in a subset of ancestries, prior variances can be shrunk towards zero to effectively allow for *ancestry-specific* causal *cis*-molQTLs.

Focusing on the *i*^*th*^ out of *k* ancestries, SuShiE models the normalized levels of a molecular trait ***g***_*i*_ measured in *n*_*i*_ individuals as a linear combination of *p* genotyped variants ***X***_*i*_ as

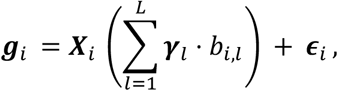

where *L* is the number of shared effects, ***γ***_*l*_ is a *p ×* 1 binary vector selecting the *l*^*th*^ causal *cis*-molQTL shared across ancestries, *b*_*i,l*_ is the *l*^*th*^ effect size in the *i*^*th*^ ancestry, and environmental noise distributed as 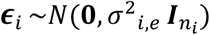 **Fig. 1A**. Following previous work^15,51,52^, we place a Multi(1, ***π***) prior over ***γ***_*l*_ where ***π*** is a *p ×* 1 vector representing prior probability for each SNP to be shared *cis*-molQTLs; however, unlike existing approaches^33,35,37,38^, we organize ancestry-specific effect sizes under a multivariate normal prior [*b*_1,*l*_, …, *b*_*k,l*_] ∼ *MVN*(**0, *C***_*l*_) where ***C***_*l*_ is the *l*^*th*^ *k × k* prior effect size covariance matrix. To perform scalable inference, we use a variational Bayesian approach and compute, for each of the *L* shared effects, the posterior probability of a shared causal *cis*-molQTL (***α***_*l*_), the ancestry-specific posterior effect sizes, and covariances, in addition to prior effect-size correlations (**Fig. 1B**) inferred through a procedure analogous to Empirical Bayes. Through learning prior effect-size correlations, SuShiE can quantify genes’ heterogeneity in *cis*-molQTL effects across ancestries.

**Fig. 1:**
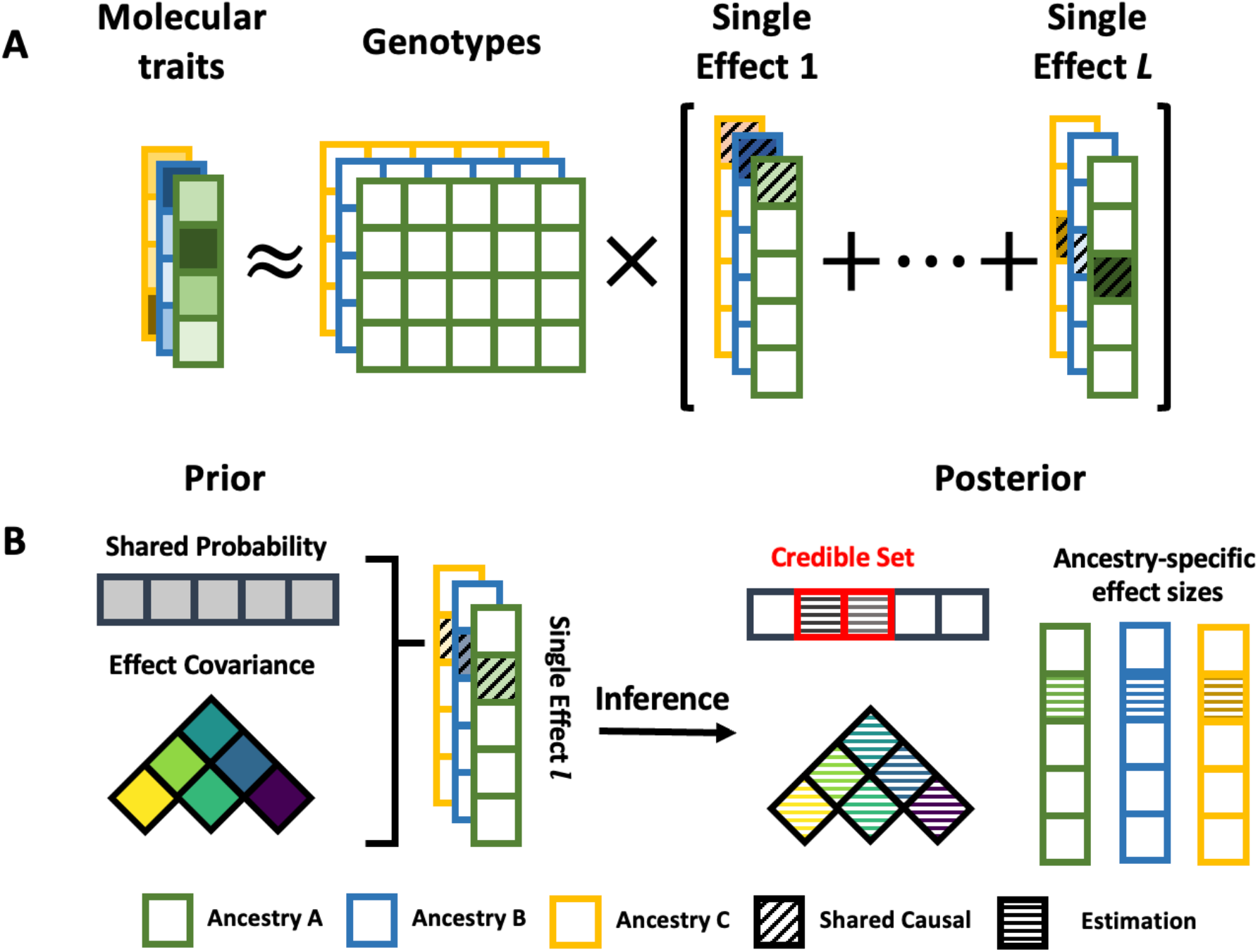
SuShiE infers ancestry-specific effect sizes, PIPs, and credible sets by leveraging shared genetic architectures and LD heterogeneity. **A)**.SuShiE takes individual-level phenotypic and genotypic data as input and assumes the shared *cis*-molQTL effects as a linear combination of single effects. **B)**.For each single shared effect, SuShiE models the *cis*-molQTL effect size follows a multivariate normal prior distribution with a covariance matrix, and the probability for each SNP to be moQTL follows a uniform prior distribution; through the inference, SuShiE outputs a credible set that includes putative causal *cis*-molQTLs, learns the effect-size covariance prior, and estimates the ancestry-specific effect sizes.

SuShiE constructs a 90*%* credible set for each of the *L* effects along with a posterior inclusion probability (PIP) for each SNP to be putative causal *cis*-molQTL (see **Methods**). SuShiE is implemented in an open-source command-line Python software with JAX (see **Methods** and **Code Availability**) using *Just-In-Time* compilation to achieve high-speed inference that runs seamlessly on CPUs, GPUs, or TPUs at https://github.com/mancusolab/sushie.

### SuShiE outperforms other methods in realistic simulations

First, to recapitulate the benefits of multi-ancestry study design^33,35,37–41^, we performed simulations varying the number of contributing ancestries under a fixed total sample size (see **Methods**). As the number of ancestries increased, SuShiE produced higher PIPs at causal *cis*-molQTLs, smaller credible set sizes, and better calibration (**Fig. S1**), reaffirming that increasing genetic diversity refines fine-mapping results compared with expanding the sample size of a single ancestry. Next, we evaluated the performance of SuShiE in simulations by varying different parameters and compared against three baselines: SuShiE-Indep (i.e., SuShiE assuming no a-priori correlation of effect sizes across ancestries), meta-SuSiE (i.e., a meta-analysis on single-ancestry SuSiE), and SuSiE (i.e., SuSiE performed over data aggregated across ancestries; see **Methods**). For all simulations, SuShiE output higher PIPs at causal *cis*-molQTLs (∼0.06 on average; all P<3.1e-4; **Fig. 2A, S2**), smaller credible set sizes (∼0.73 on average; 2 out of 3 comparisons P<0.05; **Fig. 2B, S3**), and better calibration (∼0.08 on average; all P<1.51e-7; **Fig. 2C, S4**). SuShiE similarly outperformed competing methods under simulations with differential power (**Fig. S5**) and genetic architectures across ancestries (**Fig. S6**). Next, we evaluated the ability of SuShiE to infer prior effect size correlations from data (see **Methods**). SuShiE accurately estimated primary effect size correlations (**Fig. 2D**) with higher-order effects having diminishing accuracies. This result was likely due to decreasing statistical power, as evidenced by simulations under increased sample sizes (**Fig. 2D, S7**).

**Fig. 2:**
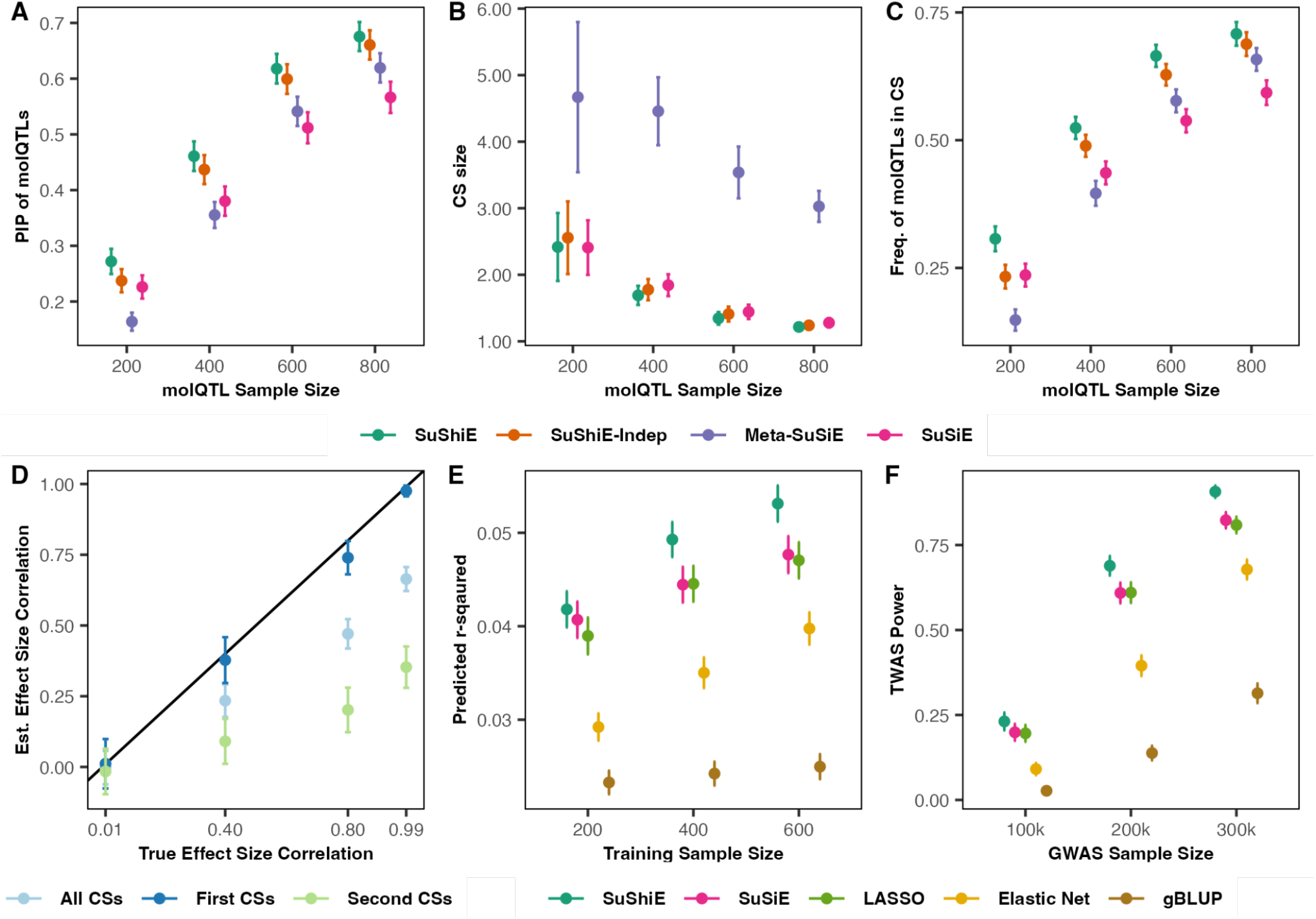
SuShiE outperforms other methods, estimates accurate effect-size correlation, and boosts higher power of TWAS in realistic simulations. **A-C**) SuShiE outputs higher posterior inclusion probabilities (PIPs; A), smaller credible set sizes (B), and higher frequency of *cis*-molQTLs in the credible sets (calibration; C) compared to SuShiE-Indep (2.60e-4, 1.5e-1, and 1.30e-11), Meta-SuSiE (P=9.67e-43, 9.35e-231, and 1.17e-76), and SuSiE (P=6.98e-63, 6.65e-2, and 1.58e-104). **D)** SuShiE accurately estimates the true effect-size correlation across ancestries using the primary effect (First credible sets; CSs) while exhibiting an underestimation using the secondary effects (Second CSs) or combined (All CSs) because the variance explained by the secondary effect decreases, thus requiring higher statistical power. The error bar is a 95*l* confidence interval. **E)** SuShiE outputs higher ancestry-specific prediction accuracy compared against SuSiE, LASSO, Elastic Net, and gBLUP (all P<9.57e-8) with the fixed sample size. The plots are aggregation across two ancestries. **F)** SuShiE induces higher TWAS power compared to SuSiE, LASSO, Elastic Net, and gBLUP (all P<4.34e-14) with the fixed sample size. The plots are aggregation across two ancestries. By default, the simulation assumes that there are 2 causal *cis*-molQTLs, the per-ancestry training sample size is 400, and the testing sample size is 200, *cis*-SNP heritability is 0.05, the effect size correlation is 0.8 across ancestries, and the proportion of *cis*-SNP heritability of complex trait explained by gene expression is 1.5e-14. The error bar is a 95*%* confidence interval.

Next, we assessed the robustness of SuShiE when there exist genetic variants causal for only a subset of ancestries in addition to shared causal *cis*-molQTLs (see **Methods**). As the number of ancestry-specific *cis*-molQTLs increased, the performance of all approaches decreased compared with previous simulations. However, SuShiE continued producing higher PIPs at shared causal *cis*-molQTLs (**Fig. S8A**), smaller credible set sizes (**Fig. S8B**), and better calibrated credible sets (**Fig. S8C**), demonstrating SuShiE’s robustness when ancestry-specific *cis*-molQTLs are present. We also evaluated performance in simulations where the number of causal effects (i.e., *L*) differs from the number specified at inference and observed that SuShiE similarly outperformed alternative approaches (**Fig. S9**).

Last, we evaluated the use of SuShiE-derived ancestry-specific effect sizes in *cis*-molQTL data as a means to predict the genetic component of gene expression for downstream TWAS^42–44^. Briefly, we performed simulations under a model in which gene expression mediates disease risk and compared SuShiE predictions with commonly used approaches for prediction-based TWAS (e.g., LASSO^53^, Elastic Net^54^, and gBLUP^55^) to identify susceptibility genes (see **Methods**). SuShiE-derived prediction models more accurately recapitulated gene expression levels compared with existing approaches and exhibited higher statistical power for TWAS with various study sample sizes and proportion of trait heritability mediated by gene expression (**Fig. 2E-F, S10**).

Overall, SuShiE outperforms existing approaches in realistic parameter settings, remains robust under model misspecifications, and improves statistical power in post-GWAS analyses.

### SuShiE identifies more functionally relevant *cis*-molQTL signals

Having verified that SuShiE outperforms other methods under realistic simulations, we next sought to perform fine-mapping on 36, 911 molecular phenotypes from diverse ancestries. Specifically, from the Trans-Omics for Precision Medicine program Multi-Ethnic Study of Atherosclerosis^48,49^ (TOPMed-MESA), we analyzed mRNA expression data of 21,747 genes measured in PBMCs (visit-1; n=956) and protein expression data of 1,274 genes measured in plasma (visit-1; n=854) for American European, African, and Hispanic ancestries (EUR, AFR, and HIS), together with mRNA expression data of 13,890 genes measured in LCLs (n=814) for EUR and AFR from the Genetic Epidemiology Network of Arteriopathy study^26^ (GENOA; see **Methods**; **Table S1**).

Focusing on 1Mb windows for each gene (i.e., *cis*-region), SuShiE fine-mapped *cis*-molQTLs for 21,088 phenotypes (e/pGenes), representing an average increase of 3,378 (16%) compared with existing methods (i.e., SuShiE-Indep, Meta-SuSiE, and SuSiE; all P<2.94e-110; see **Methods**). For example, SuShiE fine-mapped 21% more e/pGenes compared to single-ancestry SuSiE followed by meta-analysis (i.e., Meta-SuSiE; P=7.01e-238), again highlighting the benefit of multi-ancestry study design. SuShiE-based credible sets maintained higher average PIPs (∼0.07 on average) and higher frequency of *cis*-molQTLs with PIPs > 0.9 (∼0.02 on average), as well as smaller credible sets in most cases (∼6.24 on average; **Table S2**). We found the performance advantage slightly diminished in TOPMed-MESA protein and GENOA mRNA datasets, likely due to lower statistical power. Using the number of credible sets identified after purity pruning (see **Methods**), SuShiE estimated most (90.4%) molecular phenotypes to exhibit 1-3 *cis*-molQTL signals (**Fig. 3A**) with PIPs localizing near the transcription start site (TSS; **Fig. 3B**), consistent with previous studies^3,4,26,56,57^.

**Fig. 3:**
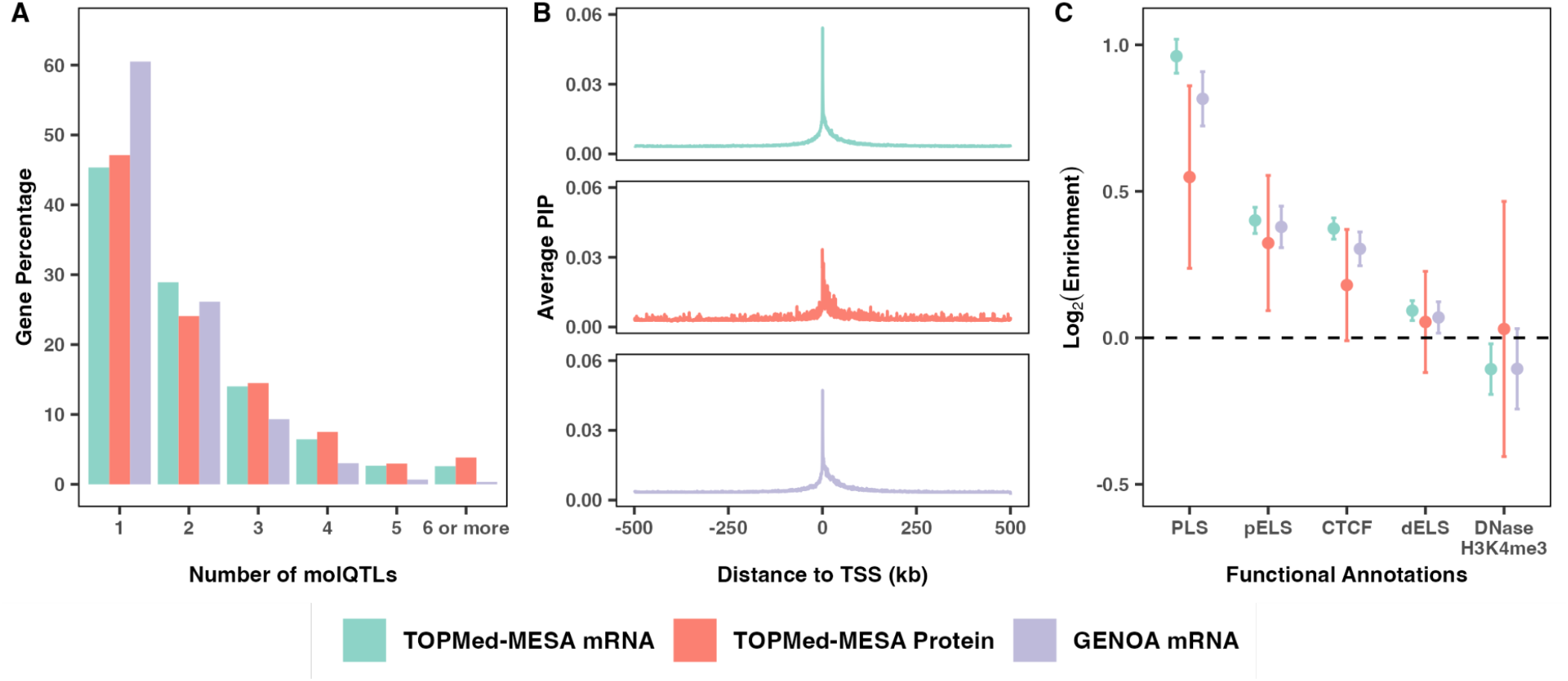
SuShiE reveals cis-regulatory mechanisms for mRNA and protein expression. **A)** SuShiE identified *cis*-molQTLs for 14,590, 573, and 5,925 genes whose 88*%*, 86*%*, and 96*%* contain 1-3 *cis*-molQTLs for the TOPMed-MESA mRNA, TOPMed-MESA protein, and GENOA mRNA dataset, respectively. **B)** Posterior inclusion probabilities (PIPs) of *cis*-molQTLs inferred by SuShiE are mainly enriched around the TSS region of genes. We grouped SNPs into 500-bp-long bins and computed their PIP average. There are 2,000 bins to cover a one-million-bp-long genomic window around the genes’ TSS. **C)** Across all three studies, *cis*-molQTLs identified by SuShiE are enriched in four out of five candidate *cis*-regulatory elements (cCREs) from ENCODE^58^, with the promoter (PLS) as the most enriched category. Specifically, the mRNA expression from TOPMed-MESA and GENOA showed enrichment in the promoter, proximal enhancer (pELS), CTCF, and distal enhancer (dELS) but depletion in DNase-H3K4me3. Protein expression from TOPMed-MESA showed enrichment in PLS and pELS but non-significant enrichment in CTCF and dELS because of the low number of genes identified with pQTLs (n=573). The error bar is a 95*%* confidence interval.

To characterize the regulatory function of identified *cis*-molQTL signals, we performed enrichment analysis using PIPs with 89 genomic functional annotations (see **Methods**). We observed that PIPs inferred by SuShiE were enriched in 83/89 annotations across all three datasets, with the highest enrichment occurring in promoter regions (**Table S3**). For example, PIPs were enriched in 4/5 candidate cis-regulatory elements (cCREs) from ENCODE Registry v3^58^ (**Fig. 3C**) and in all 10 cell-type/tissue-specific cCREs using single-nucleus(sn) or single-cell(sc) ATAC-Seq^59,60^ (**Fig. S11**). Importantly, PIPs inferred by SuShiE were more enriched across functional annotations compared with those computed from existing fine-mapping methods (all P<8.13e-3; **Table S4**), highlighting SuShiE’s ability to better prioritize functionally relevant *cis*-molQTLs. Next, to explore how potential regulatory function may differ among *cis*-molQTLs contributing to the same gene, we repeated the above analyses using per-effect posterior probabilities (***α***_*l*_), rather than overall inclusion probabilities (i.e., PIPs). First, the initial three shared effects were similarly localized near the TSS (**Fig. S12**) and were more enriched in promoter regions compared to the PIP-based analyses (**Fig. S13; Table S5**), echoing the previous finding that most genes are regulated by 1-3 *cis*-molQTLs^3,4,26,57^. Second, we found *cis*-molQTLs with weaker effects were further away from the TSS on average (**Fig. S14**), likely due to statistical power. For example, we observed the expected distance to TSS for the initial three shared effects was 84.7kb compared with 144.5 kb for the remaining shared effects (i.e., from L=6 to L=10; P=8.39e-113).

Last, we sought to validate our fine-mapping results by applying SuShiE on molecular phenotypes from three independent datasets: mRNA expression measured in PBMCs of EUR, AFR, and HIS ancestries from TOPMed-MESA^48,49^ (visit-5, ten-year after visit-1; n=875), mRNA expression measured in LCLs (n=462) of EUR and Yoruba (YRI) ancestries from GEUVADIS study^61^, and protein expression measured in plasma of EUR ancestry (N=3,301; single-ancestry SuSiE) from INTERVAL study^5^ (see **Methods**; **Table S1**). First, we confirmed SuShiE identifies 4,361 (21%; all P<2.89e-112) more e/pGenes on average compared with existing methods while obtaining higher average PIPs (∼0.07 on average), smaller credible set sizes (∼6.54 on average), and more *cis*-molQTLs with PIPs > 0.9 (∼0.04 on average) for TOPMed-MESA visit-5 and GEUVADIS (**Table S6**). Second, focusing on 20,502 e/pGenes identified by SuShiE that were also measured in validation datasets, 34% (41%, 32%, and 13% for TOPMed-MESA visit-5, INTERVAL, and GEUVADIS, respectively) *cis*-molQTLs replicated in the validation datasets with an average cosine similarity of 0.70 (0.72, 0.63, and 0.45 for the three mentioned studies; P<2e-200 for all), which increased to 73% and 0.75 respectively after conditioning on significantly heritable genes and the primary signal (see **Methods**). The diminished replication performance of GEUVADIS likely resulted from a combination of significantly reduced sample sizes, admixture differences between African YRI and American Africans in GENOA, and genotyping differences (see **Methods**). Furthermore, SuShiE exhibited similar replication ratios and cosine similarities compared to existing methods, suggesting the higher number of e/pGenes identified by SuShiE were not likely due to false positives (**Table S7**; see **Methods**).

Overall, by jointly modeling multi-ancestry data, SuShiE identifies additional *cis*-regulatory mechanisms for molecular traits.

### SuShiE identifies putative eQTL for *URGCP*

Here, we showcase a putative eQTL for *URGCP*, a gene on chromosome 7 that has been implicated in tumor growth and progression^62–66^. SuShiE fine-mapped a single SNP in TOPMed-MESA mRNA (*rs2528382*; GRCh38: 7:43926148; PIP=0.94; **Fig. 4A**), while alternative methods did not produce credible sets for this gene. Importantly, SuShiE replicated *rs2528382* in TOPMed-MESA visit-5 mRNA data. We found *rs2528382* was reported as significant in whole blood eQTL data from the eQTLGen Consortium^4^, the Study of African Americans, Asthma, Genes, and Environments (SAGE), and the Genes-Environments and Admixture in Latino Asthmatics (GALA II) study^31^, further supporting its role in regulating *URGCP* expression levels. Investigating the functional consequences of *rs2528382* using genomic annotations, we found *rs2528382* represents a non-coding exon variant within the 5’ UTR^67^, and localizes within a proximal enhancer region (pELS), as evidenced by strong signals of H3K27ac in PBMCs^58^ falling within 2kb of the TSS (**Fig. 4B**). Lastly, through snATAC-seq^59^ and scATAC-seq^60^, we found *rs2528382* localizes within an open chromatin accessibility region measured in different cell types, such as PBMCs, naive T cells, naive B cells, cytotoxic NK cells, and monocytes. Altogether, these results suggest that *rs2528382* regulates *URGCP* expression levels in PBMCs through disruption of regulatory activity.

**Fig. 4:**
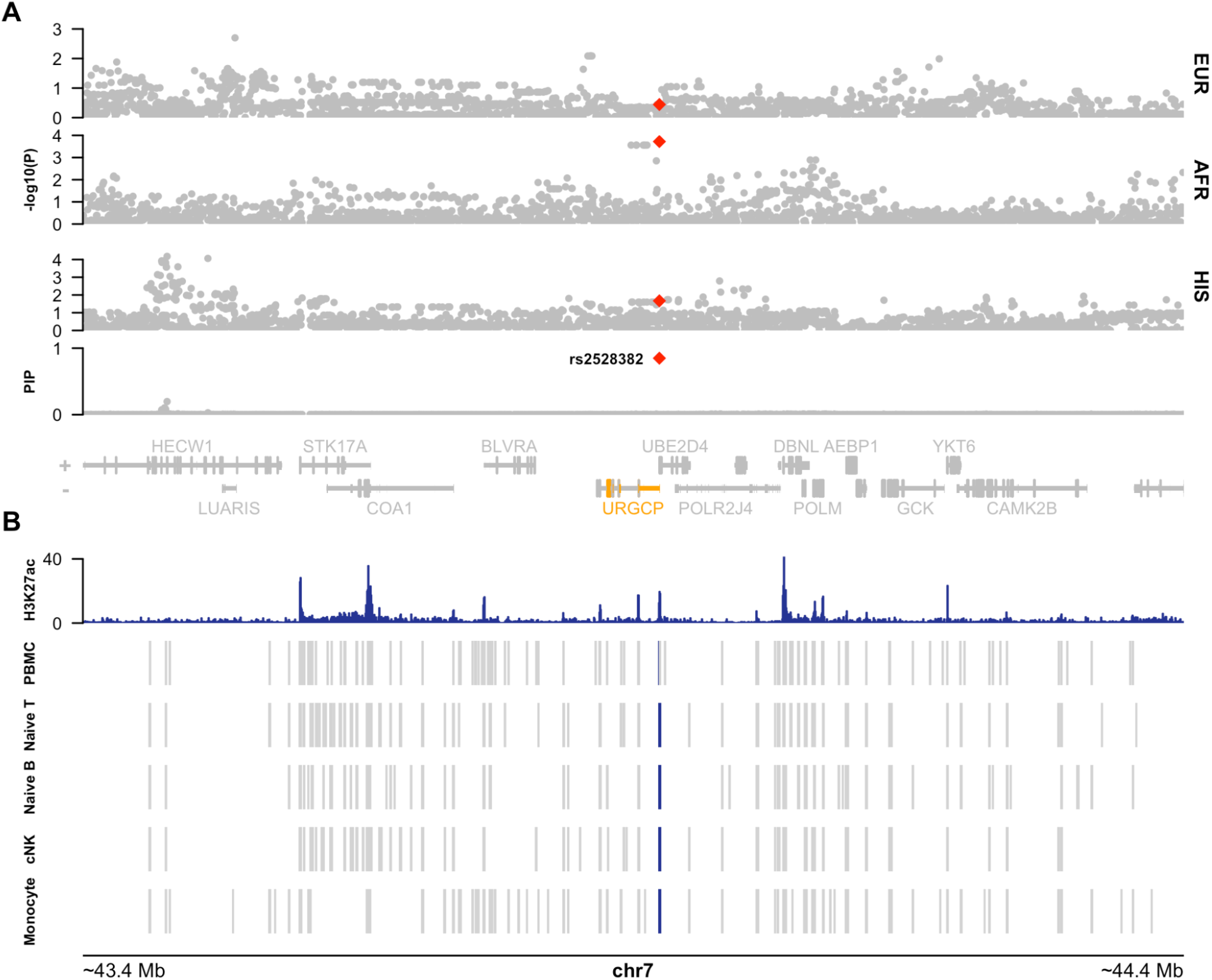
SuShiE identifies eQTL *rs2528382* for *URGCP* with functional support. **A)** Manhattan plot of *cis*-eQTL scans of *URGCP* (denoted in orange) for each ancestry (above) with SuShiE fine-mapping results (below). SuShiE was the only method to output credible sets for *URGCP* and prioritized a single SNP (*rs2528382*; denoted in red). **B)** Functional annotations at *URGCP* locus show colocalization of active enhancer activity and chromatin accessibility with *rs2528382*. H3K27ac CHIP-seq peaks measured in PBMCs (intensity denoted in blue) and 0/1 accessibility annotations determined from scATAC-seq measured in PBMCs and snATAC-seq measured in naive T cells, naive B cells, cytotoxic NK (cNK) cells, and monocytes. Blue rectangles denote a putative cCRE called from sc/snATAC-seq data that colocalize with *rs2528382* (gray no colocalization).

### SuShiE reveals heterogeneity of *cis*-molQTL effect sizes at the loss-of-function intolerant genes

After validating *cis*-molQTLs identified by SuShiE, we next sought to characterize genetic architectures of molecular traits across ancestries. First, we computed *cis*-SNP heritability for all e/pGenes of each ancestry and observed 87% significant heritable genes (in at least one ancestry) across studies (**Fig. S15**), which resulted in highly correlated estimates across ancestries (**Fig. S16**). Next, using SuShiE-derived estimates of *cis*-molQTL correlation across ancestries (see **Methods**), we found highly consistent effect-size correlations on average (0.81, 0.86, and 0.87 for EUR-AFR, EUR-HIS, and AFR-HIS, respectively), which further increased when focusing on genes whose heritabilities are significant in all ancestries (0.94, 0.98 and 0.99, respectively; 9,885 genes; 46.9%; **Figs. S17-S18**). Altogether, these results further affirm previous results^20,21,23,68–74^ demonstrating primarily shared genetic architectures for molecular traits across ancestries.

Despite this evidence, we observed a long tail of heterogeneous effect sizes (i.e., SuShiE-estimated effect size correlation <1), suggesting the presence of ancestry-specific *cis*-molQTL effects (**Fig. S19**), which is consistent with previous multi-ancestry *cis*-molQTL studies^27,31,72^. To characterize this apparent heterogeneity across ancestries, we correlated the estimated correlation signals with multiple measures of constraint (pLI^75^, LOEUF^76^, EDS^77^, RVIS^78^, and s_het_^79^) and found highly significant associations (**Table 1**; see **Methods**). Overall, genes with lower effect-size correlations across ancestries exhibited higher intolerance to loss-of-function mutations on average. For example using TOPMed-MESA mRNA dataset, we observed an average *cis*-molQTL effect size correlation of 0.81 (when L=1; SE=0.02) between EUR and AFR individuals at genes that exhibited pLI >0.9, which increased to 0.86 (when L=1; SE=0.01) when focusing on genes with pLI <0.1. Genes with high constraint exhibited lower estimates of *cis*-SNP heritability on average (**Table S8**), which may result in apparent heterogeneity arising from low statistical power. Given this, we re-analyzed putative relationships using estimated covariances, only primary signals (L=1), and bootstrapped standard errors and found broadly consistent results (**Table 1**). In addition, we observed our results were robust to adjusting for Wright’s fixation index (F_st_; **Table 1**; see **Methods**), suggesting heterogeneity/constraint associations are not driven solely by allele frequency differences across ancestries.

**Table 1:** Across-ancestry *cis*-molQTL effect size correlations are negatively associated with gene constraint scores. The estimates and corresponding P-value in the regression framework testing associations between inferred effect size correlations across ancestries and constraint scores (see **Methods** for the base model). “Bootstrap SE” is to re-estimate standard error using bootstrap. “Primary Effect” is to only use estimates from L=1. “Effect Covariance” is to replace estimated correlation with estimated effect size covariance across ancestries. “Adjusted F_st_” is to additionally adjusted for F_st_ from the base model. A higher value of pLI, s_het_, and, EDS is taken to indicate stronger constraint, while a lower value of LOEUF and RVIS is suggestive of more constraint. The reported P-value is one-sided.

|  | pLI | LOEUF | S<sub>het</sub> | RVIS | EDS |
| --- | --- | --- | --- | --- | --- |
| **Base Model** | -0.022 (4.13e-33) | 0.021 (5.92e-20) | -0.007 (4.15e-40) | 0.043 (2.04e-14) | -0.002 (1.25e-02) |
| **Bootstrap SE** | -0.022 (5.84e-32) | 0.021 (4.92e-20) | -0.007 (3.13e-37) | 0.043 (1.68e-17) | -0.002 (1.56e-02) |
| **Primary Effect** | -0.034 (3.51e-23) | 0.027 (7.45e-11) | -0.011 (7.69e-29) | 0.055 (4.09e-09) | -0.004 (1.27e-03) |
| **Effect Covariance** | -0.339 (7.59e-177) | 0.334 (1.77e-109) | -0.089 (1.33e-154) | 0.537 (9.93e-49) | -0.053 (3.10e-25) |
| **Adjusted F<sub>st</sub>** | -0.022 (2.00e-32) | 0.021 (9.90e-20) | -0.007 (2.22e-39) | 0.042 (5.63e-14) | -0.002 (1.08e-02) |

To investigate the relationship between *cis*-molQTLs identified by SuShiE and gene constraint, we first observed inverse associations between the number of fine-mapped *cis*-molQTLs per gene and constraint (**Fig. S20**), consistent with several previous studies showing the depletion of *cis*-molQTLs for high constraint genes^56,77,80^. However, we also observed positive associations between expected *cis*-molQTLs’ distance to TSS and constraint, affirming previous results that high constraint genes tended to have more complex regulatory regions^56,77^ (**Fig. S21**; see **Methods**). In addition, we correlated gene enrichment scores from ENCODE^58^ cCREs with constraint scores. We found that putative causal *cis*-molQTLs for high constraint genes tended to be enriched for distal enhancers (dELS) and depleted for promoter (PLS) and proximal enhancers (pELS) compared with weakly constrained genes, consistent with several previous studies^56,77^ (**Fig. S22**). We found these associations remained significant after accounting for F_st_, suggesting average allele frequency differences across ancestries cannot solely explain the observed heterogeneity.

Overall, SuShiE recapitulates the findings of primarily shared genetic architectures of molecular traits and show that effect size heterogeneity is consistent with gene LOF intolerance.

### Posterior *cis*-molQTL effect sizes improve T/PWAS power in white blood cell traits

Lastly, to showcase the downstream benefits of SuShiE, we performed TWAS and PWAS^42–44^ on six white blood-cell-related traits in AOU biobank^50^ (average n=86,336; **Table S9**). First, we assessed the predictive performance of SuShiE-based weights compared to alternative expression-prediction methods. Specifically, SuShiE obtained better cross-validation estimates (cv-*r*^2^) compared to SuShiE-Indep, Meta-SuSiE, SuSiE, Elastic Net and gBLUP (2 out of 5 comparisons P<0.05) and comparable estimates relative to LASSO (P=0.64; **Fig. S23A**). When focusing on genes with estimated *cis*-molQTL effect size correlation <0.9 across ancestries, we find SuShiE consistently outperformed other approaches (4 out of 6 comparisons P<0.05; **Fig. S23B**), suggesting the benefits in modeling and learning the prior effect size covariances. We observed significantly decreased prediction performance when evaluating cross-ancestry prediction (e.g., predicting mRNA expression of AFR using EUR weights; see **Methods**; P=1.71e-53; **Fig. S24**), consistent with previous works^22,27,36,81^ and further motivating ancestry-matched analyses. Given this, we predicted the expression levels of 20,515 genes (mRNA) and 573 proteins using ancestry-matched SuShiE *cis*-molQTL prediction weights from the above analyses and AOU genotypes. Overall, we identified 221 T/PWAS significant associations in white blood count (WBC), eosinophil count (EOS), and monocyte count (MON; **Table S10**; **Fig. S25**). Of these associations, ∼90% were identified in WBC due to substantially increased statistical power (i.e., 21,476 more participants on average). We found no significant associations in lymphocyte count (LYM), neutrophil count (NEU), and basophil count (BAS), likely due to low detected cell counts, similar to previous studies^36,82^ that identified fewer associations compared to models based on WBC.

Consistent with our simulation results (**Fig. 2F**), SuShiE demonstrated higher T/PWAS chi-square statistics and identified 44 more T/PWAS associations compared to results driven by SuSiE prediction weights (**Fig. 5A**). In addition, we observed that the SuShiE T/PWAS signals associated with multiple measures of LOF intolerance (**Table S11**), in contrast to previous work demonstrating that high LOF intolerance genes are typically depleted in TWAS models due to weak eQTL signals^56,77,80^ (**Fig. 5B**; see **Methods**). We found less support for a relationship between SuSiE-based TWAS signals and LOF intolerance (P=9.21e-10; **Table S11**), further demonstrating SuShiE’s advantage. To validate our results, we compared our TWAS statistics with multiple independent white blood cell-related TWASs^31,36,82–84^. Overall, we found SuShiE-based TWAS replicated at rates similar to SuSiE, suggesting that its improved power is unlikely due to false positives and further highlighting its benefit in identifying disease-related genes.

**Fig. 5:**
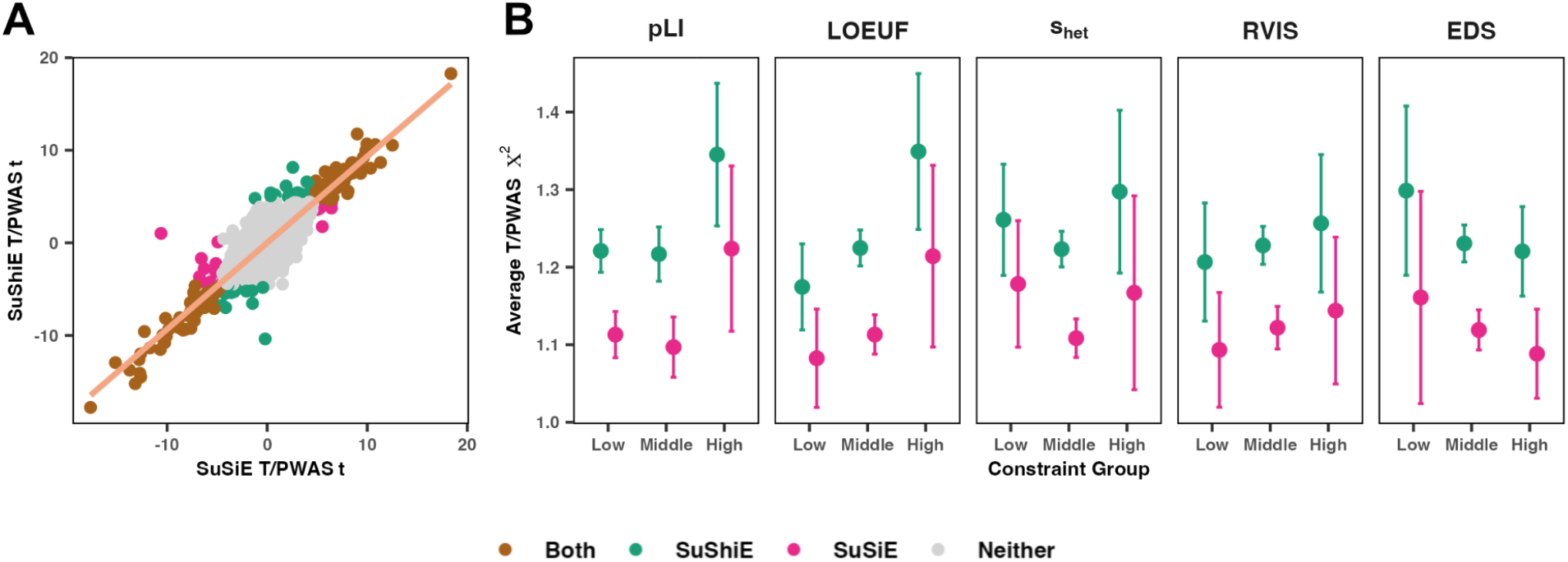
SuShiE identifies more T/PWAS genes compared with SuSiE. **A)** Scatter plot of T/PWAS t-statistics between SuShiE (y-axis) and SuSiE (x-axis) across all phenotypes and contributing *cis*-molQTL studies. **B)** Average T/PWAS chi-square statistics within low, middle, and high constraint scores (see **Methods**). Error bars represent 95*%* confidence intervals.

Overall, our work has shown that by jointly modeling the molecular data across different ancestries while allowing effect sizes to differ, SuShiE outputs more accurate *cis*-molQTL prediction weights, thus boosting the downstream statistical power for integrative analyses with GWASs.

## Discussion

Here, we present the Sum of Shared Single Effect approach (SuShiE), a novel approach for multi-ancestry SNP fine-mapping of molecular traits using a scalable variational approach. SuShiE assumes the joint *cis*-molQTL effects arise as a linear combination of per-ancestry effect sizes across shared causal variants. Through extensive simulations, SuShiE first improved the fine-mapping precision in disentangling the causal *cis*-molQTLs from tagging SNPs by leveraging LD heterogeneity across diverse ancestries. Second, SuShiE accurately learned prior effect size correlations across ancestries employing a procedure analogous to Empirical Bayes. Third, SuShiE estimated ancestry-specific cis-molQTL prediction weights, boosting findings in the post-GWAS framework (e.g., TWAS and PWAS), compared to the baselines that did not model effect size covariance across ancestries or ignored ancestry altogether. We applied SuShiE to 36,911 molecular phenotypes of diverse ancestries from three datasets: mRNA and protein expression from TOPMed-MESA and mRNA expression from GENOA. SuShiE fine-mapped 16% more genes on average compared to the existing methods, exhibiting smaller credible set sizes and higher enrichment in relevant functional annotations. SuShiE inferred highly correlated *cis*-molQTL effect sizes across ancestries on average in significantly heritable genes, reflecting primarily shared *cis*-molQTL architectures. In addition, we observed *cis*-molQTL effect size heterogeneity across ancestries associated with multiple constraint measurements, consistent with environmental interactions may partially drive differences in effect sizes across ancestries. Last, we performed TWAS and PWAS on six white blood cell-related traits from AOU biobank using SuShiE-derived ancestry-specific *cis*-molQTL prediction weights and identified 44 more significant genes compared to the existing method. We also observed that SuShiE T/PWAS signals are associated with multiple measures of LOF intolerance, further showing the benefit of multi-ancestry approaches in identifying genes relevant to complex disease risk.

Next, we describe caveats in our real data analysis. First, SuShiE approximates ancestry as a discrete category, allowing us to model *cis*-molQTL effect sizes using a multivariate normal distribution (see **Methods**). While this simplifies modeling and inference tasks, we emphasize that this is a heuristic approach that neglects the complex and shared demographic histories underlying all humans. Indeed, recent work has demonstrated the importance of viewing genetic ancestries as a continuous spectrum rather than discrete categories^85^. Relatedly, previous studies^33,35,37–41^ and our simulation results (**Fig. S1**) have shown that increasing the number of ancestries within a multi-ancestry framework improves fine-mapping precision. However, SuShiE and similar frameworks perform inference on variants present after filtering on MAF thresholds (e.g., 1%) within each ancestry. As a result, this requirement can exclude *cis*-molQTLs from analysis due to small sample sizes within an ancestry, suggesting a trade-off in practice between increasing overall sample size versus excluding informative genetic variants. For instance, we obtained mRNA expression data measured in EUR (n=402), AFR (n=175), HIS (n=277), and East Asian ancestry (EAS; n=96) individuals from TOPMed MESA study visit-1. From two-ancestry fine-mapping (EUR and AFR) to three-ancestry (+HIS), we filtered an additional 29 SNPs per gene on average. However, this number increased to 501 SNPs by including the additional 96 participants of EAS ancestry. As a result, we opted to not include EAS participants in our analysis in order to maximize the genetic variants analyzed. Modeling genetic ancestry continuously can potentially avoid this type of *cis*-molQTL loss, thus improving the fine-mapping precision with a larger sample size.

Second, we note that our data consist of African-(AFR) and Hispanic-American (HIS) individuals, which contain recent admixture events. To account for complex diversity within ancestries, we included genotyping PCs as a covariate in our models. Several works have suggested that admixture can be sufficiently corrected for using global ancestry information (i.e., genotyping PCs) in association testing^73,86–91^, especially when causal effect sizes are largely consistent across ancestries^86,87,89^ (**Fig. S16-S18**). On the other hand, accounting for local ancestry may increase the associating testing power when causal effects are highly different across ancestries^86,87,92^ or aid fine-mapping in post-GWAS analysis^87,89,93^, which can be one of the future directions for SuShiE.

Third, we observed significant associations between gene LOF intolerance and several SuShiE-estimated metrics, including effect size heterogeneity across ancestries, the number of *cis*-molQTLs, *cis*-molQTL distance to TSS, and functional enrichments. The relationship remained significant after adjusting for F_st_, suggesting allele frequency differences across ancestries are not sufficient to fully explain estimated heterogeneity. As a result, we hypothesized that *cis*-molQTL effect size heterogeneity could be in part due to gene-by-environment (GxE) interactions^69,77,94–96^. Highly constrained genes exhibit more complex regulatory landscapes with fewer *cis-* molQTLs (or apparent *cis-*molQTLs due to smaller effect sizes)^56,77^. As a result, these genes may be less resilient to environmental perturbations^77^, which may induce effect-size heterogeneity across different ancestries. On the other hand, it is possible that our F_st_ estimates are underpowered to detect subtle allele frequency differences across ancestries. Therefore, these associations may provide indirect evidence for natural selection partially driving *cis*-molQTL effect size heterogeneity across ancestries. To explicitly investigate the role of selection in molecular differences across ancestries, we likely require a more principled modeling procedure based in population genetics together with higher-resolution molecular data measured in diverse ancestries^56,80,97–100^. For instance, recent work has shown the promise of using single-cell data to demonstrate how selection impacts genes expressed differentially across ancestries^101^.

Fourth, SuShiE assumes causal *cis*-molQTLs are shared across ancestries. Our simulations show that SuShiE remains robust when ancestry-specific *cis*-molQTLs are present (**Fig. S8**). However, in situations where there exist shared *cis*-molQTLs but ancestries have different sample sizes, SuShiE may prioritize shared *cis*-molQTLS along with SNPs tagged in LD of the ancestry with larger sample sizes, evidenced through simulations (**Fig. S5B**). However, through our case study in *URGCP* (**Fig. 4**), we observed relatively higher signals in AFR but not in EUR and HIS, despite AFR having the smallest sample size, suggesting this limitation may be minimal overall.

Lastly, in our T/PWAS analysis, we selected six white blood-cell related traits to best match PBMC and LCL contexts. However, alternative cell-types not included in our analyses may better capture relevant contexts. For example, PBMCs and LCLs do not contain neutrophils, basophils, and eosinophils, and LCLs additionally do not include monocytes, which may result in a loss in statistical power. As single-cell RNA-seq datasets become more available^102^, one possible direction would be to perform TWAS in fine-grained cellular contexts and backgrounds. In addition, after predicting expression levels using ancestry-matched weights for each individual, we performed individual-level T/PWAS by concatenating the predicted expression levels across ancestries rather than perform ancestry-specific TWAS followed by meta-analysis^103^. The premise of the meta-analysis approach is that researchers obtain ancestry-specific GWAS and then integrate with corresponding eQTL weights. Because the causal genes for complex traits are likely shared across ancestries^20,21,23,36,68–74^, a regression framework with individual-level data concatenated across ancestries (the largest sample size) can maximize power.

We briefly discuss potential directions for future work. First, recent studies have shown that incorporating functional annotation in the prior distribution can improve the fine-mapping precision^33,79,104^. SuShiE currently employs a uniform distribution for prior causal probability. Including functionally-informed priors is likely to improve its performance further. Second, SuShiE fine-maps individual-level molecular and genotypic data in a prespecified locus flanking the TSS and TES regions of a gene. In theory, users can apply SuShiE on individual-level complex trait data, however, this likely will require additional analyses (e.g., pre-specifying GWAS significant loci) and care in controlling for genome-wide backgrounds and population structure. In addition, the limited accessibility to the individual-level complex trait data allows the extension of SuSiE-like models to be compatible with summary statistics^38,39,41,51^, which typically requires external LD reference panels. As more *cis*-molQTL summary statistics are available to the community^4,102^, we foresee a potential demand to implement this compatibility in SuShiE. Last, SuShiE currently cannot model molecular data in their original read-count format, which is usually transformed to a continuous scale (i.e., inverse normal transformation). Extending SuShiE to a GLM-like model naturally would encompass this scenario and present an exciting direction for SuShiE.

Overall, SuShiE, together with its application on large-scale molecular data of diverse ancestries, identifies more *cis*-regulatory mechanisms and reveals its genetic architecture. We anticipate considerable demand for our approach in the genetics field characterized by forthcoming multi-ancestry and multi-omics research.

## Online Methods

### Sum of Shared Single Effects Model

Here, we describe the statistical model underlying SuShiE (see **Supplementary Note** for a detailed description). SuShiE assumes *cis*-molQTLs are present in *all* ancestries, defined as shared *cis*-molQTLs while allowing for effect sizes at causal *cis*-molQTLs to covary across ancestries a-priori. For the *i*^*th*^ of total *k* ancestries, SuShiE models the centered and standardized levels of a molecular trait ***g***_*i*_ measured in *n*_*i*_ individuals as a linear combination of *p* genotyped variants ***X***_*i*_ as

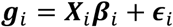

where ***β***_*i*_ is a *p ×* 1 vector of ancestry-specific *cis*-molQTL effects, and 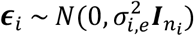 is environmental noise. In addition, we model 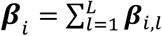 as the sum of *L* effects ***β***_*i,l*_ = ***γ***_*l*_ · *b*_*i,l*_ where ***γ***_*l*_ is a *p ×* 1 binary vector indicating which variant is the shared *cis*-molQTL for the *l*^*th*^ effect while allowing ancestry-specific effect sizes *b*_*i,l*_. Furthermore, we model ***γ***_*l*_ ∼ Multi(1, ***π***) where ***π*** is a *p ×* 1 vector representing prior probability for each SNP to be a *cis*-molQTL, and model *b*_*l*_ = [*b*_1,*l*_ *⋯ b*_*i,l*_ *⋯ b*_*k,l*_ P ∼*N*(**0, *C***_*l*_) where

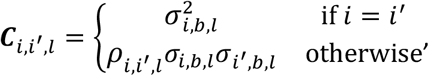

***C***_*l*_ is the *l*^*th*^ prior *k × k* effect size covariance matrix with 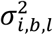 as variance, and *𝜌*_*l*_ as correlation.

### Variational inference of model parameters

To infer the *cis*-molQTL effects, we seek to estimate the posterior distribution of 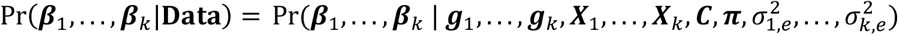 where ***C*** = {***C***_1_, …, ***C***_*L*_}. We regard ***β***_∗_ as latent variables, ***g***_∗_, and ***X***_∗_, as observed data, and ***C, π***, and 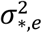 are the hyperparameters. However, inferring the exact distributions of latent variables is computationally intractable due to non-conjugacy with the prior distribution. Therefore, we seek a surrogate distribution *Q*(***β***_1_, …, ***β***_*k*_), which minimizes the Kullback–Leibler (KL) divergence with Pr(***β***_1_, …, ***β***_*k*_|**Data**). Specifically, we have:

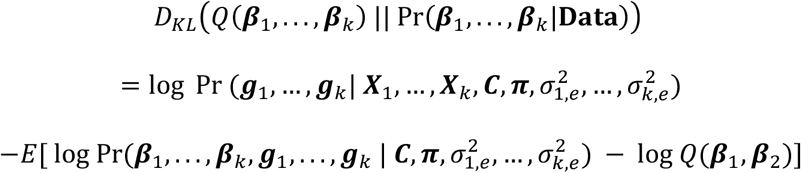

where the first term is the log evidence, and the expectation term is the evidence lower bound (ELBO). Since the log evidence is constant with respect to model variables, minimizing the KL divergence is equivalent to maximizing the ELBO. Furthermore, to limit the universe of possible forms that the surrogate distribution *Q*(***β***_1_, …, ***β***_*k*_) may take, we impose an additional mean-field assumption^105^. Namely, SuShiE assumes that each of the *L* shared effects *β*_*l*_ are mutually independent under *Q*:

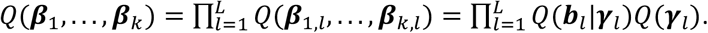

Therefore, to approximate the posterior distributions *Q*(·) for latent variables ***b***_*l,j*_ (a *k ×* 1 vector) and ***γ***_*l,j*_ (a scaler) at SNP *j ∈* [1, *p*] of *l*^*th*^ shared effect, we need to compute the expectation of complete data log-likelihood *L*(***β***_1_, …, ***β***_*k*_, ***g***_1_, …, ***g***_*k*_ | ***C, π***, 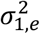, …, 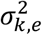) (i.e., the joint distribution) while holding other variables constant.

Through the principles of coordinate-ascent variational inference (CAVI)^105^, we can identify each *Q*(·) surrogate as,

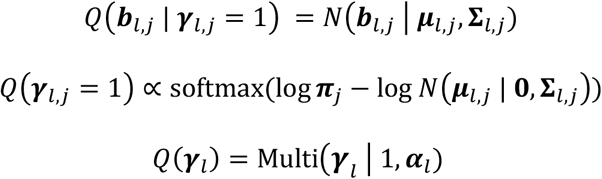

Where. ***μ***_*i,j*_ *∈* ℝ^*k×*1^ and ***∑***_*l,j*_ *∈* ℝ^*k×k*^ are the corresponding posterior mean and covariance, and ***α***_*l*_ *∈* ℝ^*p×*1^ is each SNP’s posterior probability to explain the *l*^*th*^ effect. We provide the complete mathematical derivations, inference algorithms, and detailed definitions in the **Supplementary Note**

### Computing posterior inclusion probability and *η*-credible sets

We define the posterior inclusion probability (PIP) for SNP *j* with 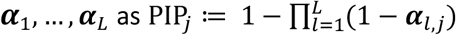 To compute an *η* -credible set for each *L*, where *η* represents the desired probability that the set contains *cis*-molQTLs, we decreasingly sort ***α***_*l*_ and take a greedy approach to include SNPs until their cumulative sum exceeds *η*.

In the case that the inferred number of effects *L* surpasses the actual number of *cis*-molQTLs, the unnecessary credible sets will contain most SNPs with low posterior probability close to ***α***_*l,j*_ = 1/*p*, where *p* is the number of SNPs. To refine the final inference results, we remove the credible sets whose lowest absolute pairwise correlation, which is defined as “purity”^15^ and weighted by sample size across all ancestries, among SNPs is less than 0.5. In practice, following the previous work^15^, we empirically specify *L* as 10.

### Inferring cross-ancestry effect size correlations

SuShiE features the capability to estimate the correlation of *cis*-molQTL effect sizes across multiple ancestries. For some gene *t*, SuShiE by default outputs *L* estimates of the effect size correlation 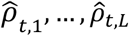 for each credible set. If we apply SuShiE to *T* genes in total, we empirically recommend computing effect size correlation across ancestries with 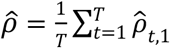

### Simulating genotypes and quantitative molecular traits

To evaluate SuShiE’s performance in simulations, we first simulated genotypes and quantitative molecular traits to mimic the real-world scenarios using our previous simulation frameworks^36,106,107^. To simulate genotype data, we used LD estimates from individuals of European (EUR; n=489), African (AFR; n=639), and East Asian (EAS; n=481) ancestries from the 1000 Genomes Project (1000G) phase three data^108^. We limited LD to biallelic HapMap SNPs^109^, discarded those with missingness (>1*%*), MAF (<1*%*), and violated Hardy-Weinberg equilibrium (HWE mid-adjusted P< 1e-6). We obtained chromosome, transcription start site (TSS), and transcription end site (TES) information for 19,279 protein-coding autosomal genes using GENCODE release 26 (GRCh37)^110^. We extended each gene 500,000 base pairs (bp) upstream of TSS and 500,000 bp downstream of TES, and randomly selected 500 genes that have at least 500 common SNPs across EUR, AFR, and EAS genotypes.

We first focused on simulations using EUR and AFR (*k* = 2). At each gene, we simulated centered and standardized genotype matrix 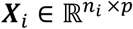 for *i*^*th*^ ancestry using a multivariate normal distribution *N*(0, ***V***_*i*_) where *n*_*i*_ *∈* {200, 400, 600, 800} is the *cis*-molQTL study sample size, *p* is the number of common SNPs across ancestries in the locus, and ***V***_*i*_ *∈* ℝ^*p×p*^ is the ancestry-specific LD matrix estimated from 1000G genotypes^108^. Next, we uniformly chose *m ∈* {1, 2, 3} out of *p* common SNPs as *cis*-molQTLs and simulated their ancestry-specific effect sizes 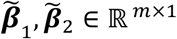 under a bivariate normal distribution as

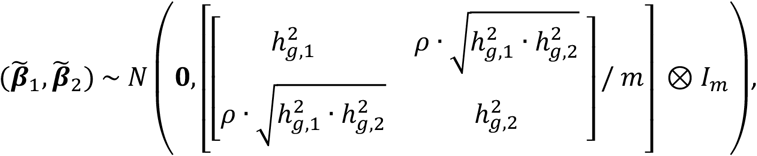

where 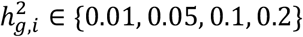 is the proportion of variance in gene expression explained by *cis*-molQTLs (i.e., *cis*-SNP heritability of the molecular trait) and *𝜌 ∈* {0.01, 0.4, 0.8,0.99} is the effect size correlation. Then, we constructed effect-size vectors ***β***_1_ and ***β***_2_, where 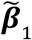 and 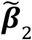 are the *m* non-zero entries at the same index representing shared *cis*-molQTLs and the rest *p* ™ *m* entries are zero representing the null SNPs. Next, we computed the quantitative molecular traits ***g***_*i*_ using ***X***_*i*_***β***_*i*_ + *ϵ*_*i*_ where 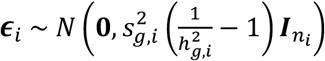 is the random environmental noise and 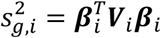 is the genetic variance after accounting for LD. To reflect cases where heterogeneity exists in the genetic architecture of molecular traits across ancestries^31,72^, we allowed *cis*-SNP heritability to be ancestry-specific with 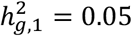 and 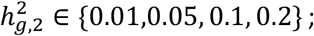 we also evaluated the performance under different statistical power where *n*_1_ = 400 and *n*_*_ *∈* {200, 400, 600, 800}. To determine whether incorporating additional ancestry improves SuShiE’s performance, we simulated the genotypic and phenotypic data for EAS with the same total sample sizes and genetic architecture.

In addition, we simulated two cases under model misspecification. We first evaluated SuShiE’s performance when ancestry-specific *cis*-molQTLs exist, we simulated *m*_*i,AS*_ *∈* {1, 2, 3} *cis*-molQTLs for both ancestries in addition to shared *cis*-molQTLs *m* = 2 while fixing 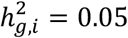 for ancestry *i*. Second, to reflect cases where the number of shared *cis*-molQTL (*m*) is different from inferred *L* by fixing *m* = 2 and varying the inferred *L ∈* {2, 5, 10}.

### Default parameters and performance metrics

We performed SNP fine-mapping using SuShiE on simulated genotypes and molecular data across EUR and AFR individuals. In terms of variational inference parameters, we specified *L ∈* {1, 2, 3} to match the actual number of simulated effects and initialized *cis*-molQTL effects 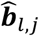 as 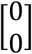 their covariance matrix *Ĉ*_*l*_ as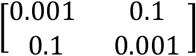, the prior estimates of environmental noises 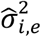 as 0.001, the prior probability for SNPs to be *cis*-molQTLs as 1/*p* where *p* is the number of common SNPs.

To evaluate the gain in parametrizing the effect size correlation across ancestries, we compared our method SuShiE to “SuShiE-Indep” which assumes the *cis*-molQTL effect sizes are independent across ancestries; that is, we fixed the effect size correlation prior *𝜌* = 0, and did not learn it through the Empirical-Bayes-like procedure. To demonstrate that SuShiE’s improvement does not result from the accumulation of samples across ancestries, we compared SuShiE’s performance to two “baseline” methods: first, we performed single-ancestry SuSiE and then meta-analyzed the resulting PIPs by PIP_meta_ = 1 ™ (1 – PIP_EUR_) · (1 – PIP_AFR_); we refer to this method as “meta-SuSiE”. Second, we row-stacked the genotype matrices and molecular trait vectors across ancestries and then performed single-ancestry SuSiE as “SuSiE.” Overall, we performed four methods (SuShiE, SuShiE-Indep, meta-SuSiE, and SuSiE) on 500 genes’ simulated genotypes and molecular traits to output corresponding PIPs, credible sets, and ancestry-specific effect size estimates. We varied four parameters: per-ancestry *cis*-molQTL study sample size (*n*_*i*_), the number of *cis*-molQTLs (*m*), the *cis*-SNP heritability of molecular traits (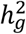) for each ancestry, and the effect size correlation (*𝜌*). To reflect a practical study design, the default parameters were fixed at *n*_*i*_ = 400, *m* = 2, 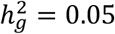, and *𝜌* = 0.8 unless stated otherwise. Furthermore, we evaluated the fine-mapping performance with three metrics across 500 simulated genes: PIPs at causal *cis*-molQTLs, credible set size, and frequency that causal *cis*-molQTLs are contained in 90*%* credible sets (calibration). We computed the metrics of meta-SuSiE based on the union of the credible sets across two single-ancestry SuSiE. As different methods may or may not prune credible sets at the same simulated gene, to show a fair comparison, we computed the credible set size metric only using the credible set that none of the four methods pruned out. To compare metrics across methods, we ran linear regression adjusted for relevant simulation parameters and reported one-sided Wald test P values.

### Simulating GWAS and TWAS

Transcriptome-wide Association Studies (TWASs) leverage GWAS summary statistics, eQTL prediction weights, and LD reference to identify genes whose predicted expression levels are associated with complex traits^42–44^. A more accurate eQTL prediction weight will increase the power of the TWAS framework. Therefore, we compared the prediction weights inferred by SuShiE to other methods: SuShiE-Indep, Meta-SuSiE, SuSiE, least absolute shrinkage and selection operator (LASSO)^53^, elastic net regularization (Elastic Net)^54^, and genomic best linear unbiased prediction (gBLUP)^55^. We simulated the expression and genotype data for the training and testing set separately, with the same method mentioned in the previous sections. For the training set, we varied the per-ancestry sample size *n*_*t*_ *∈* {200, 400, 600, 800} and set the out-of-sample testing set sample size *n*_v_ = 200. Then, we predicted the expressions using ancestry-matched fitted weights on testing genotype data, and computed the coefficients of determination (*r*^2^) between the predicted and simulated expression. For Meta-SuSiE, we trained the prediction weights for each ancestry using per-ancestry sample size. For SuSiE, LASSO, Elastic Net, and gBLUP, we trained the prediction weights after concatenating data across ancestries to guarantee that the total sample sizes were the same as SuShiE as fair comparisons.

To showcase that SuShiE’s prediction weights introduce more power in TWAS, we simulated GWAS summary statistics and computed TWAS statistics using different prediction weights. First, because individuals in GWASs are usually different from ones in the eQTL studies, we re-simulated the genotype matrix 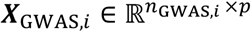 where *n*_GWAS,*i*_ is the GWAS sample size for ancestry *i* using the same generating approach above. Then, we used the eQTL effect size vectors ***β***_*i*_ generated in the previous section to simulate a complex trait 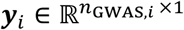 as a linear combination of expression levels 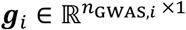 as

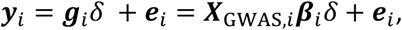

where *δ*∼*N*(0,1) is the gene expression effect on the complex trait, 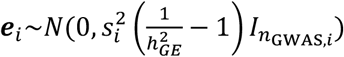 is the random noises for the complex traits, 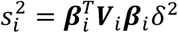, ***V***_*i*_ is the LD matrix generated from 1000G^108^, and 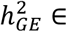 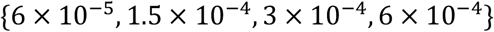 is the proportion of variation of the complex trait explained by the expression of a single gene^111^. Then, we regressed the complex trait ***y***_*i*_ on each SNP in ***X***_GWAS,*i*_ marginally to compute the GWAS summary statistics ***z***_GWAS,*i*_ *∈* ℝ^*p ×*1^. Last, we computed TWAS summary statistics with 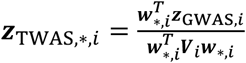 along with its P value where ***w***_∗,*I*_ is the prediction weights fitted by different methods. We define the TWAS power as the frequency of the Bonferroni-corrected P value is less than 0.05.

### Overview of real-data analyses

We applied SuShiE and other methods (e.g., SuShiE-Indep, Meta-SuSiE, and SuSiE) to three datasets: mRNA (visit-1) measured in peripheral blood mononuclear cells (PBMCs) and protein expression measured in plasma of three EUR, AFR, and HIS ancestries from Trans-Omics for Precision Medicine program Multi-Ethnic Study of Atherosclerosis (TOPMed MESA)^48,49^ and mRNA expression measured in lymphoblastoid cell lines (LCLs) of EUR and AFR ancestries from the Genetic Epidemiology Network of Arteriopathy (GENOA) study^26^. We excluded the mRNA expression levels data measured in T cells and monocytes from TOPMed MESA study due to relatively smaller sample sizes. We explain the detailed quality control (QC) procedure in the sections below. We conducted pairwise comparisons of methods on four basic summary statistics, focusing on the genes for which both methods output credible sets; the summary statistics included the number of genes identified with *cis*-molQTLs (e/pGenes), the average PIPs of the SNPs in the credible sets, the average single-effect-specific credible set sizes, and the frequency of having genes whose credible sets contained SNPs with PIPs greater than 0.9. We defined the number of *cis*-molQTLs as the number of credible sets output after pruning for purity (see previous section for the definition). Next, we performed enrichment analyses using 89 functional annotations and a case study focusing on a gene that was only identified by SuShiE, and missed by other methods. Last, using SuShiE-derived ancestry-specific *cis*-molQTL effect sizes, we performed individual-level TWAS and PWAS with All Of Us (AOU) biobank^50^ individuals and compared to the results derived from SuSiE.

To validate SuShiE’s results on the three main datasets mentioned above, we applied SuShiE and other methods to three separate datasets: mRNA expression (visit-5) measured in PBMC of EUR, AFR, and HIS ancestries from TOPMed MESA, protein expression measured in plasma of EUR ancestry from INTERVAL study^5^, and the mRNA expression measured in LCL of EUR and Yoruba in Ibadan (YRI) ancestries from the GEUVADIS study^61^. We computed two statistics to evaluate validation performance: first, focusing on the *cis*-molQTLs of e/pGenes identified by SuShiE, the percentage for which SuShiE identified *cis*-molQTLs in the validation datasets. Second, focusing on the credible sets for which we identified the same *cis*-molQTLs in both main and validation studies, we computed the cosine similarity of posterior probabilities (***α***_*l*_) to see whether they prioritized the same SNPs. For SNPs that are not in the overlap between main and validation studies, we manually assigned them a value of 0 for cosine similarity calculation. For each credible set, we randomly shuffled the ***α***_*l*_ in validation studies 500 times to construct the null distribution of the cosine similarity and compute its z score. We computed the average cosine similarity and z scores across all credible sets as an aggregation estimate and its corresponding significance. For all the fine-mapping analysis, we used the SNPs that are shared across ancestries on the genomic window of each gene that is 500,000 bp upstream and downstream of each gene’s TSS and TES (one million bp in total), respectively, based on the GENCODE v34^110,112^. In addition, we only included genes that are located on the autosomes, do not overlap with the major histocompatibility complex (MHC) region, have more than 100 SNPs on the genomic window present in all ancestries, and whose ENSEMBL gene IDs match the records in GENCODE v34^110,112^. We adjusted for covariates by regressing them from both mRNA/protein levels and each SNP. In addition, we computed the *cis*-SNP heritability using the limix python package (see **Code Availability**) for each analyzed molecule within each ancestry. We used PLINK2.0, vcftools, and bcftools for genotype manipulation^113–116^.

### Genotype data in the TOPMed MESA study

We obtained the whole-genome sequencing (WGS) data (freeze 9; GRCh 38) of 5,379 individuals from the TOPMed MESA^48,49^. Specifically, we removed the SNPs with the following criteria: both duplicate genotype discordance and mendelian genotype discordance are greater than 2*%*, genotype missing rate at depth 10 is greater than 2*%*, Milk-SVM score for variant quality is less than -0.5, variants that overlap with centromeric regions, HWE p-value is less than 1e-6, and MAF is less than 1*%*, resulting in a total of 125,089,612 SNPs. In addition, we computed the genotype principal components (PCs) with SNPs that are pruned for LD using PLINK2.0 (--indep-pairwise 200 1 0.3)^113,114,117^. Last, we retained individuals who are self-identified as EUR, AFR, or HIS ancestries and have measurements in mRNA (both visits 1 and 5) and protein datasets, resulting in a total of 1,292 individuals.

### mRNA expression data in the TOPMed MESA study

We obtained RNA-seq data in gene-level read counts and reads per kilobase of transcript per million mapped reads (RPKM) of 57,615 genes for 2,137 samples (both visits-1 and visit-5) measured in PBMC using RNA-SeQC v2.0.0 from the TOPMed MESA study. The data was pre-processed based on the TOPMed RNA-seq harmonization pipeline (see **Code Availability**). We first calculated the gene expression PCs on all samples’ read counts using the PCA function of the scikit-learn package^118^, and normalized it across all samples within each PC. Then, focusing on the samples measured in visit-1, we followed the GTEx^3^ eQTL analysis preparation script to select gene whose transcript per million (TPM) is >0.1 and raw read counts >6 reads in at least 20*%* of samples (see **Code Availability**). For individuals with replicate samples, we only kept one sample with the greatest sum of reads across all genes; we also removed individuals with whom we did not have self-identified ancestry information, resulting in 402 EUR, 175 AFR, and 277 HIS individuals. Then, within each ancestry, we normalized expression levels between samples using edger_cpm function in the pyqtl package, (see **Code Availability**) with normalized_lib_sizes=True, which is a Python implementation of edgeR^119^ ; we next performed inverse-rank normalization using the inverse_normal_transform function. Last, focusing on 21,747 genes filtered based on inclusion criteria and using SNPs whose MAF >1*%* and HWE mid-adjusted P>1e-6 within each ancestry, we ran SuShiE and other methods using SNPs on the genomic windows of each gene, adjusting for 15 gene expression PCs, 10 genotype PCs, age, sex, and the assay lab. We did not include individuals who self-identified as East Asian in TOPMed MESA study due to the small sample size (n=96). We removed SNPs based on MAF <1*%*, and including EAS participants would exclude 501 more SNPs on average per gene from downstream analyses.

### Protein expression data in the TOPMed MESA study

We obtained the protein expression levels of 1,317 target proteins for 1,966 samples (both visits-1 and -5) from the TOPMed MESA study using SOMAscan, an aptamer-based technology. First, we computed the protein expression PCs on all samples using the PCA function of the scikit-learn package^118^, and normalized it across all samples for each PC. Then, focusing on the samples measured in visit-1, we removed individuals with whom we did not have self-identified ancestry information, resulting in 398 EUR, 297 AFR, and 261 HIS individuals. Within each ancestry, we inverse-rank normalized the protein expression data using the inverse_normal_transform function in the pyqtl package (see **Code Availability**). As some proteins may be targeted by multiple aptamers, which correspond to different isoforms of proteins^120^, we regarded each isoform as a unique protein. As a result, we obtained 1,274 proteins based on gene inclusion criteria and performed fine-mapping using SuShiE and other methods on the genomic windows adjusted for 15 protein expression PCs, 10 genotype PCs, sex, and age, using SNPs whose MAF > 1*%* and HWE mid-adjusted P>1e-6 within each ancestry.

### Genotype and mRNA expression data in the GENOA study

From the GENOA study^26^, we obtained paired genotype and LCL mRNA expression data of 373 EUR and 441 AFR individuals, together with corresponding covariates, processed by previous works^26,36^. Briefly, we restricted TOPMed-imputed^121^ genotype data on biallelic SNPs with imputation score *r*^2^ > 0.6, MAF >1*%*, and HWE mid-adjusted P>1e-6 within each ancestry. Focusing on 14,797 genes based on gene inclusion criteria, we performed fine-mapping on the genomic window, adjusted for 30 gene expression PCs, five genotype PCs, age, sex, and genotyping platform.

### Genotype and molecular data in three validation datasets

To validate SuShiE’s results of PBMC mRNA expression (visit-1) in TOPMed MESA^48,49^, we used the mRNA expression data measured in PBMC of the same study but collected from visit-5, a 10-year-later follow-up visit. We performed the identical pipeline mentioned in the previous section, resulting in 21,695 genes (21,240 overlapped with visit-1) from 422 EUR, 168 AFR, and 285 HIS individuals.

To validate the plasma protein expression results in TOPMed MESA, we obtained the inverse-rank normalized protein expression levels of 3,301 EUR individuals measured in plasma from the INTERVAL study^5^. The genotype data was pre-processed, imputed, and annotated with dbSNP v153 by previous studies^5,122,123^. We obtained 3,187 ENSEMBLE-UniProt-SOMAmer ID triplets (1,313 overlapped with the TOPMed MESA) based on gene selection criteria and performed singe-ancestry SuSiE fine-mapping on the genomic window, adjusted for sex, age, duration between blood draw and process, 3 genotype PCs, and subcohort, and 5 expression PCs, using SNPs whose MAF >1*%* and HWE mid-adjusted P>1e-6.

To validate the mRNA expression data measured in LCLs from the GENOA study, we obtained paired genotype and gene expression data measured in LCLs in gene-level RPKM of 23,722 genes for 373 EUR and 89 YRI individuals from the GEUVADIS study^61^. First, we computed the expression PCs on all the individuals using the PCA function of the scikit-learn package^118^. Then, we kept high-expressed genes whose TPM >0.1 in at least 20% of all the individuals^3^ and filtered based on gene selection criteria, resulting in a total of 19,882 genes (10,439 overlapped with GENOA). Last, using SNPs whose MAF >1*%* and HWE mid-adjusted P>1e-6 within each ancestry, we performed SuShiE fine-mapping on the genomic window, adjusted for sex, five expression PCs, and five genotype PCs, which is calculated on the LD-pruned pipeline defined in the previous section.

### Functional enrichment analyses and case study

We ran functional enrichment analysis only on the genes identified with *cis*-molQTLs (i.e., SuShiE outputs credible sets; e/pGenes). To visualize the relationship between the PIPs inferred by SuShiE and their distance to the TSS, we grouped fine-mapped SNPs into 2,000 bins that are 500 bp long to cover the one-million-bp window around the TSS for each gene and computed the average PIPs within each bin. To visualize the relationship between single effects’ posterior probabilities and their distance to the TSS, we performed the same procedure focusing on the shared effects that had credible set output (i.e., passed the purity threshold; see previous method section).

We performed enrichment analysis using 89 functional annotations. First, we downloaded 5 candidate cis-regulatory elements (cCREs) from ENCODE Registry v3^58^. Then, we obtained 9 cell-type specific cCREs measured in PBMC using snATAC-Seq^59^ and one cCRE measured in frozen PBMC using scATAC-seq^60^. Last, we obtained the 74 categorical functional annotations from LDSC baseline annotations v2.2^124,125^, and remapped to GRCh38 using LiftOver (see **Code Availability**). To compute the functional enrichment scores, we employed an approach that is similar to TORUS^126^. Briefly, for each functional annotation and each gene, we performed the logistic regression *g*(***P***) = ***a****ω* where *g*(*⋅*) is the logit link function, ***P*** is the vector for the PIPs of all the SNPs, ***a*** is the binary vector indicating whether the SNPs fall into the annotation, and *ω* is the desired log-enrichment scores. After removing the genes on which logistic regression does not converge, we meta-analyzed the log-enrichment scores across genes by 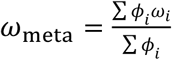 and 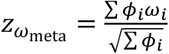 where *ϕ*_*i*_ is the inverse of the squared standard error for gene *i*. When comparing enrichment results across methods, we focused on e/pGenes fine-mapped by both methods. We computed the comparison z score as 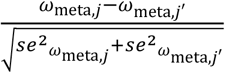 for method *j* and *j*′. For the enrichment analyses focusing on individual shared effect using ***α***_*l*_, rather than PIPs, we limited analyses to those single effects that had corresponding credible sets (i.e., were not pruned).

To perform a case study, we selected *URGCP*, which was fine-mapped by SuShiE, but missed by other methods. To annotate the genomic region around *URGCP*, we downloaded the ChiP-Seq H3K27ac data of ENCODE^58^ from WashU Epigenome Browser^127^ (see **Code Availability**) and proximal enhancer (pELS) cCREs from ENCODE Registry v3, PBMC annotation using scATAC-seq in Satpathy et al.^60^, naive T cells, naive B cells, cytotoxic natural killer (cNK) cells, and monocytes annotations using snATAC-seq in Chiou et al.^59^

### Prior *cis*-molQTL correlation analyses

To shed light on the relationship between heterogeneity of effect-sizes across ancestries and genes’ constraint, using all the credible sets output by SuShiE, we tested for association between SuShiE-inferred effect size correlations across ancestries (***𝝆***_*l*_) and five measures of constraint (***s***) using all the fine-mapped e/pGenes: probability of being Loss-of-Function Intolerant (pLI)^75^, loss-of-function observed/expected upper bound fraction (LOEUF)^76^, enhancer-domain score (EDS)^77^, the Residual Variation Intolerance Score (RVIS)^78^, and s_het_^79^. We downloaded pLI and LOEUF from gnomAD browser v4.0 (see **Code Availability**), we downloaded EDS, RVIS, and s_het_ from their original papers. Our base model is according to:

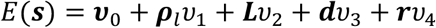

where ***v***_0_ is the intercept term, ***L*** is the ordered and categorical single effect index representing the order of variance explained, ***d*** is the corresponding ancestry pair indicator (e.g., the correlation of EUR-AFR, EUR-HIS, or HIS-AFR), ***r*** is the study indicator (e.g., TOPMed MESA mRNA, TOPMed MESA proteins, or GENOA mRNA), *v*_*i*_s are the corresponding coefficients. We test the significance of *v*_1_ in a linear regression framework. A negative value of *v*_1_for pLI, EDS, and s_het_ is taken to indicate stronger associations between *cis*-molQTL effect size heterogeneity across ancestries and gene constraint, while a lower value of LOEUF and RVIS is suggestive of stronger associations. In addition, to show robustness, we re-tested these associations using estimated covariance by replacing ***𝝆***_*l*_ by ***σ***_*b*_^2^. We also only focused on correlations estimated only from the primary effect (i.e., L=1); in this case, we removed ***L*** from the base model. We also re-computed the standard error using bootstrap. Specifically, for each study, each ancestry pair, and each *L*, we sampled the genes with replacement and computed the *v*_1_. We repeated 100 times to construct the null distributions for *v*_1_ and used its standard deviation as a new standard error. In addition, to adjust for allele frequency differences across ancestries, we added Wright’s fixation index (F_st_) as an additional term. To compute F_s,_, we only used the fine-mapped SNPs to compute the F_st_ using PLINK2^113,114^ with the “Hudson” method^128,129^ for each gene. To investigate the relationship between expected *cis*-molQTLs’s distance to TSS and genes’ constraint, we computed the expected distance to TSS for each gene according to 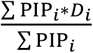 where *D* is the distance (absolute value) to the TSS for SNP *i*.

### TWAS and PWAS analyses in All Of Us biobank

We performed individual-level Transcriptome- and Proteome-wide Association Studies (TWASs and PWASs)^42–44,47^ on 6 white blood cell-related traits: basophil count (BAS), eosinophil count (EOS), lymphocyte count (LYM), monocyte count (MON), neutrophil count (NEU), and white blood cell count (WBC; **Table S9**) measured in AOU biobank^50^. We excluded individuals who had acute abdomen, acute appendicitis, acute cholangitis, acute cholecystitis, acute pancreatitis, anemia due to and following chemotherapy, bone marrow transplant present, chemotherapy-induced nausea and vomiting, cirrhosis of liver, clostridium difficile colitis, complication of chemotherapy, congenital anemia, congenital hemolytic anemia, convalescence after chemotherapy, dermatosis resulting from cytotoxic therapy, diverticulitis of intestine, end-stage renal disease, fatigue due to chemotherapy, hereditary hemolytic anemia, human immunodeficiency virus infection, leukemia, mucositis following chemotherapy, myelodysplastic syndrome (clinical), neutropenia due to and following chemotherapy, pancytopenia due to antineoplastic chemotherapy, peripheral neuropathy due to and following antineoplastic therapy, post-splenectomy disorder and post-splenectomy thrombocytosis. For WBC, we only included measurements <200e9/L. For all the traits, we also excluded measurements that were 3 standard deviations away from the mean, resulting in a total of 86,345 individuals on average. We identified individual ancestry information based on AOU precomputed information (i.e., “eur”, “afr”, and “amr” labels), resulting in 53,268 EUR, 16,748 AFR, and 16,329 HIS individuals on average.

From our previous analysis, we obtained the eQTL prediction weights of EUR, AFR, and HIS in the TOPMed MESA mRNA dataset, the pQTL prediction weights of EUR, AFR, and HIS in the TOPMed MESA protein dataset, and the eQTL prediction weights of EUR and AFR in the GENOA mRNA dataset. We evaluated the prediction accuracy for SuShiE SuShiE-Indep, Meta-SuSiE, SuSiE, LASSO^53^, Elastic Net^54^, and gBLUP^55^ with five-fold cross-validation. For Meta-SuSiE, we trained the prediction weights for each ancestry. For SuSiE, LASSO, Elastic Net, and gBLUP, we trained the prediction weights after concatenating genotype and phenotype data across ancestries to ensure the equal sample sizes as SuShiE (i.e., the same prediction weights for all ancestries). We computed cross validation *r*^2^ (cv-*r*^2^) between the measured expression levels and predicted expression levels concatenated across each fold and each ancestry. We also used the SuShiE-based ancestry-specific prediction weights to evaluate the prediction performance using cross-ancestry weights. Specifically, we predicted the expression levels of EUR individuals using AFR weights (of AFR individuals using HIS weights and of HIS individuals using EUR weights).

To perform T/PWAS, we first predicted expression levels (either mRNA or proteins) for EUR, AFR, and HIS individuals in AOU using each ancestry-matched e/pQTL prediction weights with the score function in PLINK2^113,114^. Then, we standardized the expression vector (centered by mean and scaled by standard deviation) within each ancestry and then concatenated them into a single vector across ancestries. Then, we regressed out sex, age, squared age, and ten genotype PCs from the trait measurements. Last, we regressed the inverse-rank normalized residuals on the predicted expression levels to compute the TWAS or PWAS statistics. We re-performed the procedure using SuSiE-derived e/pQTL prediction weights as comparisons. We applied the Bonferroni correction to adjust the reported P-values with n=23,000. To validate our TWAS results, we compared them to five independent TWAS studies: Lu and Gopalan et al.^36^, Kachuri et al.^31^, Tapia et al.^82^, Rowland et al.^84^, and Wen et al.^83^ We released our *cis*-molQTL prediction weights to the public, which can be found at the Data Availability section. To test the association between T/PWAS chi-square statistics and genes’ constraint scores: pLI^75^, LOEUF^76^, EDS^77^, RVIS^78^, and s_het_^79^, we used linear regression adjusted for phenotype and study and reported one-sided P values. To compare significance of these associations between SuShiE and SuSiE, we computed the z score as 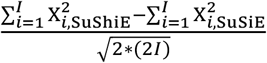 where 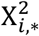 is the chi-square statistics for constraint score *i*. We classified genes into three groups: Low, Middle, and High based on different scores, respectively. For pLI, we labeled genes with pLI >0.9 as High, <0.1 as Low, and otherwise middle. For other scores, we labeled genes whose value is greater than 90*%* quantile as High, smaller than 10*%* quantile as Low, and otherwise middle.

### High-speed inference of SuShiE using JAX

We implemented SuShiE in an open-sourced command-line Python software *sushie*, which can read individual-level genotype data in three formats: PLINK1.9^113,114^, bgen1.3^130^, and vcf^116^, together with phenotypic and covariates data in tab-separated-values format. We leveraged *Just In Time* compilation in JAX (see **Code Availability**) to facilitate high-speed inference on CPUs, GPUs, or TPUs. This technique allows users to process, in a scalable fashion, thousands of molecular phenotypes with the backgrounds of diverse ancestries specified by the user. Not only can *sushie* perform our method, but it can also perform single-ancestry SuSiE^15^, effect size correlation estimation, *cis*-SNP heritability estimation, cross-validation for the *cis*-molQTL prediction weights, and contain the script to convert the *cis*-molQTL prediction results to FUSION format^42^, thus can be used in TWAS framework. We also implemented basic QC on the input data. Users can also customize the *sushie* inference function according to their preferences. We have compiled comprehensive documentation about the software at https://mancusolab.github.io/sushie/.

## Data Availability

SuShiE-derived prediction models for TWAS/PWAS, fine-mapping, and other analyzed results across cis-molQTL datasets can be found at https://zenodo.org/records/10963034.

https://github.com/mancusolab/sushie

https://github.com/mancusolab/sushie-project-codes

https://zenodo.org/records/10963034

## Code availability

SuShiE: https://github.com/mancusolab/sushie

The analysis codes for simulation and real-data analysis of this manuscript: https://github.com/mancusolab/sushie-project-codes

TOPMed RNA-seq Harmonization pipeline: https://github.com/broadinstitute/gtex-pipeline/blob/master/TOPMed_RNAseq_pipeline.md

gnomAD v4.0: https://gnomad.broadinstitute.org/news/2023-11-gnomad-v4-0/

GTEx eQTL analysis pipeline: https://www.gtexportal.org/home/methods

pyqtl software: https://github.com/broadinstitute/pyqtl

PLINK: https://www.cog-genomics.org/plink/2.0

BCFTOOLS: https://samtools.github.io/bcftools/bcftools.html

JAX: https://github.com/google/jax

scikit-learn: https://scikit-learn.org/stable/

FUSION: http://gusevlab.org/projects/fusion/

limix: https://github.com/limix/limix

LiftOver: https://genome.ucsc.edu/cgi-bin/hgLiftOver

WashU Epigenome Browser: https://epigenomegateway.wustl.edu/

## Acknowledgements

The authors would like to thank members of the Mancuso and Gazal labs for fruitful discussions regarding this manuscript. The authors would also like to specially thank Dr. Michael D. Edge for his thoughtful comments and suggestions. This work was funded in part by National Institutes of Health (NIH) under awards R01HG012133, R01CA258808, R01GM140287, R35GM142783, R01GM140287, U54HG013243, R35GM147789, and K08HL159346.

MESA phenotypes (dbGaP: phs000209.v13.p3): MESA and the MESA SHARe project are conducted and supported by the National Heart, Lung, and Blood Institute (NHLBI) in collaboration with MESA investigators. Support for MESA is provided by contracts HHSN268201500003I, N01-HC-95159, N01-HC-95160, N01-HC-95161, N01-HC-95162, N01-HC-95163, N01-HC95164, N01-HC-95165, N01-HC-95166, N01-HC-95167, N01-HC-95168, N01-HC-95169, UL1-TR-001079, UL1-TR000040, UL1-TR-001420, UL1-TR-001881, and DK063491. Funding for SHARe genotyping was provided by NHLBI Contract N02-HL-64278. TOPMed MESA WGS genotype, mRNA, and protein expression data (dbGaP: phs001416.v3.p1): Molecular data for the Trans-Omics in Precision Medicine (TOPMed) program was supported by the National Heart, Lung and Blood Institute (NHLBI). WGS genotype data for NHLBI TOPMed: MESA (phs001416.v3.p1) was performed at Broad Genomics (HHSN268201600034I). mRNA expression data for NHLBI TOPMed: MESA (phs001416.v3.p1) was performed at NWGC (HHSN268201600032I). SOMAscan proteomics for NHLBI TOPMed: Multi-Ethnic Study of Atherosclerosis (MESA) (phs001416.v1.p1) was performed at the Broad Institute and Beth Israel Proteomics Platform (HHSN268201600034I). Core support including centralized genomic read mapping and genotype calling, along with variant quality metrics and filtering were provided by the TOPMed Informatics Research Center (3R01HL-117626-02S1; contract HHSN268201800002I). Core support including phenotype harmonization, data management, sample-identity QC, and general program coordination were provided by the TOPMed Data Coordinating Center (R01HL-120393; U01HL-120393; contract HHSN268201800001I). We gratefully acknowledge the studies and participants who provided biological samples and data for TOPMed.

GENOA genotype (dbGaP: phs001238.v2.p1) and gene expression (GEO: GSE138914) data were supported by grants from NIH NHLBI (HL054457, HL054464, HL054481, HL119443, and HL087660). The authors would like to acknowledge Drs. Sharon Kardia and Jennifer Smith in preparing GENOA eQTL data.

The All of Us Research Program is supported by the National Institutes of Health, Office of the Director: Regional Medical Centers: 1 OT2 OD026549; 1 OT2 OD026554; 1 OT2 OD026557; 1 OT2 OD026556; 1 OT2 OD026550; 1 OT2 OD 026552; 1 OT2 OD026553; 1 OT2 OD026548; 1 OT2 OD026551; 1 OT2 OD026555; IAA *i*: AOD 16037; Federally Qualified Health Centers: HHSN 263201600085U; Data and Research Center: 5 U2C OD023196; Biobank: 1 U24 OD023121; The Participant Center: U24 OD023176; Participant Technology Systems Center: 1 U24 OD023163; Communications and Engagement: 3 OT2 OD023205; 3 OT2 OD023206; and Community Partners: 1 OT2 OD025277; 3 OT2 OD025315; 1 OT2 OD025337; 1 OT2 OD025276. In addition, the All of Us Research Program would not be possible without the partnership of its participants.

## Author contributions

Z.L. and N.M. developed the model and study design. Z.L. performed simulations and fine-mapping analyses. Z.L., X.W., J.P., and L.K. performed TWAS and AoU analyses. Z.L., M.C., and N.M. developed the model and inference scheme. Z.L. and A.K. prepared functional genomic annotations and enrichment analyses. Z.L. and N.M. wrote the initial manuscript. All authors edited the final manuscript.

