## Supplementary Figures for "Improved multi-ancestry fine-mapping identifies *cis*-regulatory variants underlying molecular traits and disease risk"

1. Center for Genetic Epidemiology, Keck School of Medicine, University of Southern California, Los Angeles, CA, USA
2. Department of Population and Public Health Sciences, Keck School of Medicine, University of Southern California, Los Angeles, CA, USA
3. Department of Quantitative and Computational Biology, University of Southern California, Los Angeles, CA
4. Center for Immunity and Immunotherapies, Seattle Children's Research Institute, Seattle, WA, USA
5. Department of Pediatrics, University of Washington School of Medicine, Seattle, WA, USA
6. Department of Genome Sciences, University of Washington, Seattle, WA, USA
7. Cancer Epidemiology Division, Population Sciences in the Pacific Program, University of Hawaiʻi Cancer Center, University of Hawaiʻi at Mānoa, Honolulu, HI, USA
8. Harvard Medical School and Dana-Farber Cancer Institute, Boston, MA, USA
9. Division of Cardiology, University of California San Francisco, San Francisco, CA, USA
10. Department of Epidemiology and Population Health, Stanford University School of Medicine, Stanford, CA, USA
11. Stanford Cancer Institute, Stanford University School of Medicine, Stanford, CA, USA
12. Corresponding Author

Contacts:

1. Zeyun Lu
2. Nicholas Mancuso

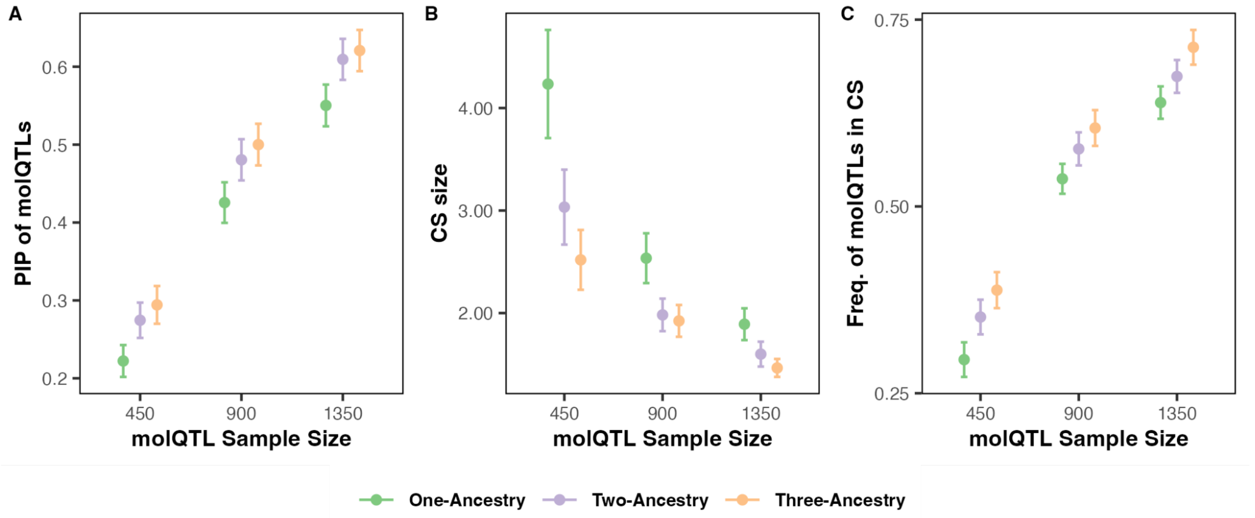


**Fig. S1: Multi-ancestry study designs improve fine-mapping results compared to a single-ancestry study design with fixed sample sizes.**

Three-ancestry study design outputs higher posterior inclusion probabilities (PIPs; A), smaller credible set size (B), and better calibration (C) than two-ancestry (P=5.46e-2, 1.95e-2, and 1.50e-4, respectively) and one-ancestry (P=3.27e-12, 4.62e-18, and 9.93e-17, respectively). The simulation assumes 2 causal *cis*-molQTLs, *cis*-SNP heritability is 0.05, and the *cis*-molQTL effect size correlation across ancestries is 0.8. The error bar is a 95% confidence interval.

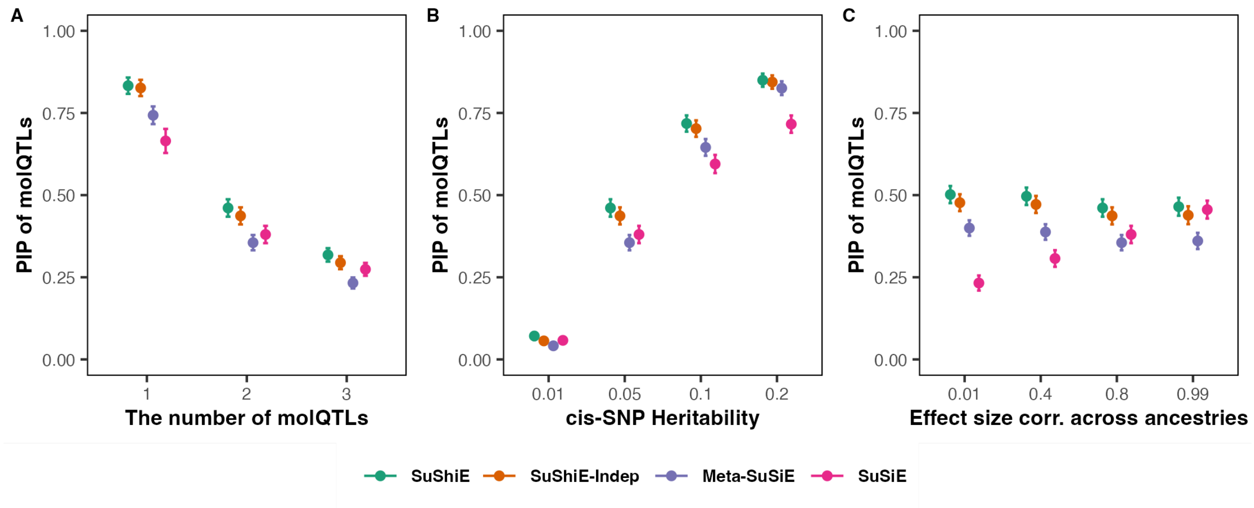


**Fig. S2: SuShiE outputs higher PIP of *cis*-molQTLs when varying different simulation parameters**

SuShiE outputs higher posterior inclusion of probability (PIPs) when varying the number of *cis*-molQTLs (A), the cis-SNP heritability (B), and the *cis*-molQTL effect size correlation across ancestries (C) compared against SuShiE-Indep (P=3.10e-4), Meta-SuSiE (1.68e-45), and SuSiE (2.26e-80). The P value is computed across all simulations (see **Methods**). By default, the simulation assumes a per-ancestry sample size of 400, 2 causal *cis*-molQTLs, *cis*-SNP heritability of 0.05, and the *cis*-molQTL effect size correlation across ancestries of 0.8. The error bar is a 95% confidence interval.

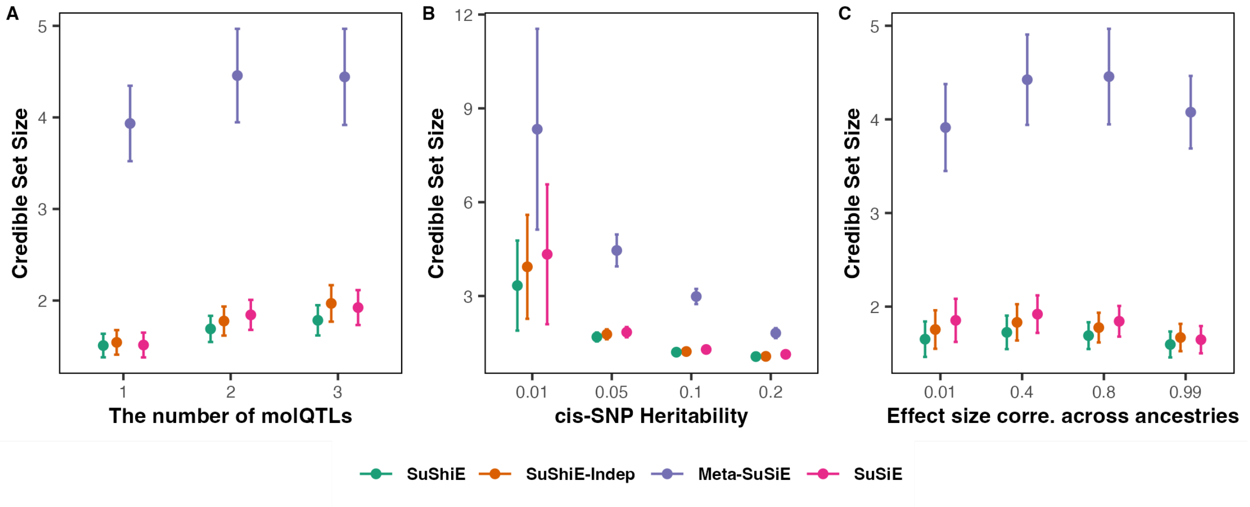


**Fig. S3: SuShiE outputs smaller credible set sizes when varying different simulation parameters**

SuShiE outputs smaller credible set sizes when varying the number of *cis*-molQTLs (A), the cis-SNP heritability (B), and the *cis*-molQTL effect size correlation across ancestries (C) compared against SuShiE-Indep (P=1.18e-1), Meta-SuSiE (5.20e-291), and SuSiE (2.94e-2). The P value is computed across all simulations (see **Methods**). By default, the simulation assumes a per-ancestry sample size of 400, 2 causal *cis*-molQTLs, *cis*-SNP heritability of 0.05, and the *cis*-molQTL effect size correlation across ancestries of 0.8. The error bar is a 95% confidence interval.

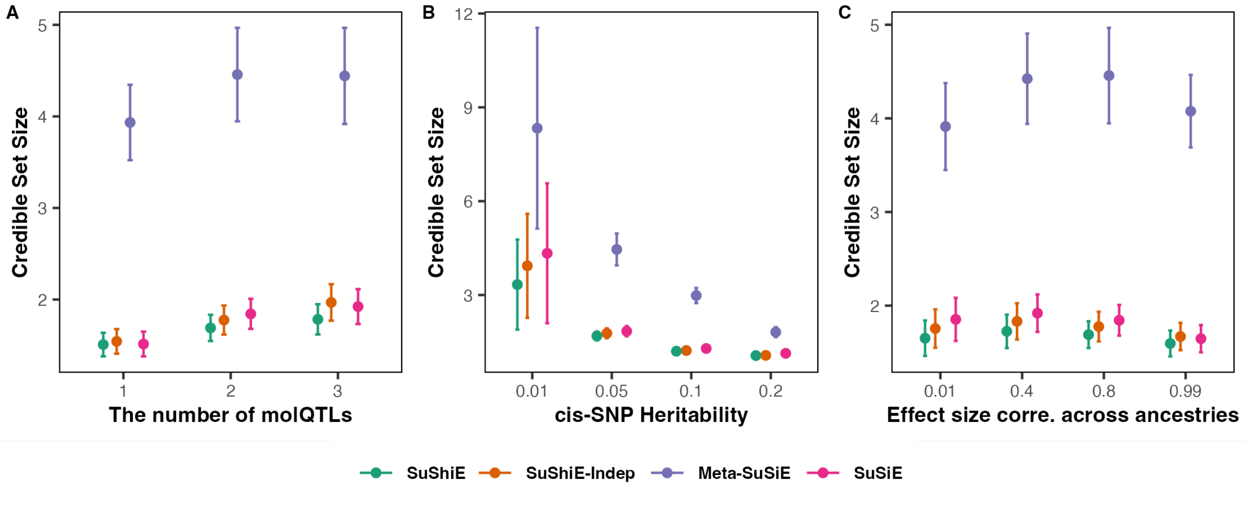


**Fig. S4: SuShiE outputs better calibration when varying different simulation parameters**

SuShiE outputs higher frequency of *cis*-molQTLs in the credible set (calibration) when varying the number of *cis*-molQTLs (A), the cis-SNP heritability (B), and the *cis*-molQTL effect size correlation across ancestries (C) compared against SuShiE-Indep (P=1.51e-7), Meta-SuSiE (9.41e-70), and SuSiE (1.03e-131). The P value is computed across all simulations (see **Methods**). By default, the simulation assumes a per-ancestry sample size of 400, 2 causal *cis*-molQTLs, *cis*-SNP heritability of 0.05, and the *cis*-molQTL effect size correlation across ancestries of 0.8. The error bar is a 95% confidence interval.

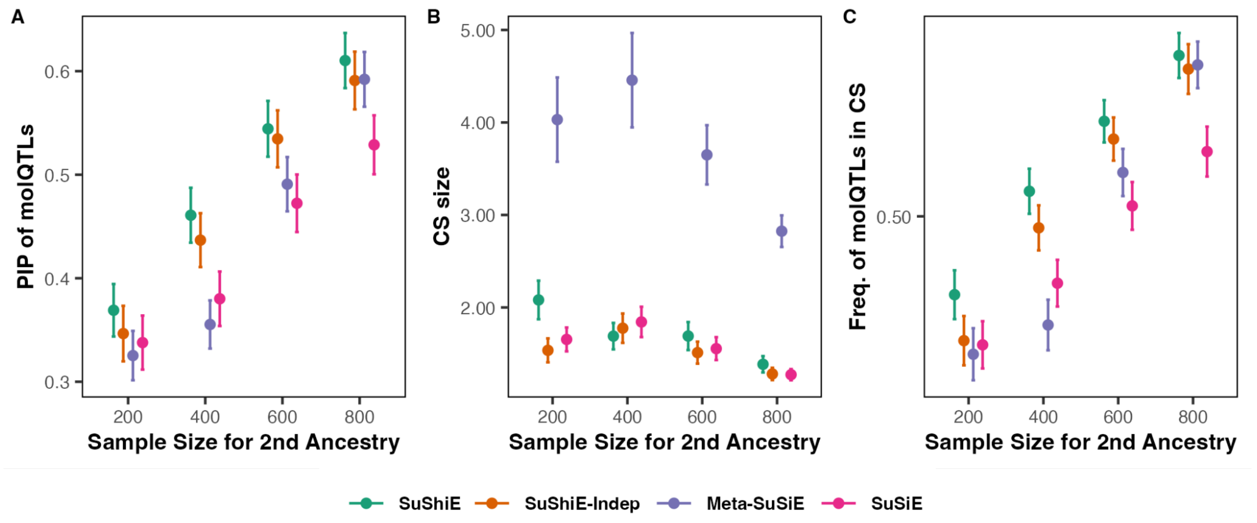


**Fig. S5: SuShiE outputs higher PIPs and better calibration when varying the sample size for one ancestry and fixing for the other**

SuShiE outputs higher posterior inclusion probability (PIPs; A) and higher frequency of *cis*-molQTLs in the credible sets (Calibration; C) compared against SuShiE-Indep (P=2.38e-2 and 4.45e-4), Meta-SuSiE (P=3.49e-9 and 6.88e-14), and SuSiE (P=1.81e-12 and 2.77e-21) when varying the sample size for one ancestry and fixing for the other. However, SuShiE's performance is not consistent in credible set size compared against SuShiE-Indep (larger; P=7.23e-3; B), Meta-SuSiE (smaller; P=2.79e-144), and SuSiE (larger; P=3.61e-2). The P value is computed across all simulations (see **Methods**). By default, the simulation assumes the first ancestry's sample size of 400, 2 causal *cis*-molQTLs, *cis*-SNP heritability of 0.05, and the *cis*-molQTL effect size correlation across ancestries of 0.8. The error bar is a 95% confidence interval.

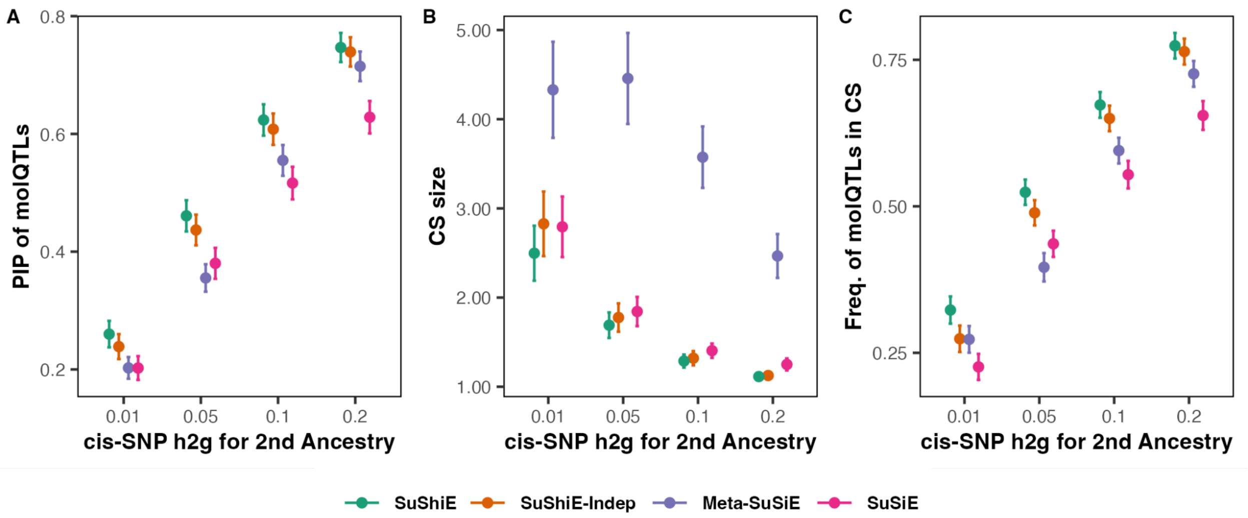


**Fig. S6: SuShiE outputs higher PIPs, smaller credible set size, and better calibration when varying the *cis*-SNP heritability for one ancestry and fixing for the other**

SuShiE outputs higher posterior inclusion probability (PIPs; A), smaller credible set size (B), and higher frequency of *cis*-molQTLs in the credible sets (Calibration; C) compared against SuShiE-Indep (P=2.77e-2, 1.83e-1, and 1.64e-4), Meta-SuSiE (P=8.46e-14, 1.92e-140, and 6.30e-21), and SuSiE (P=1.36e-24, 2.37e-2, 1.65e-38) when varying the *cis*-SNP heritability for one ancestry and fixing for the other. The P value is computed across all simulations (see **Methods**). By default, the simulation assumes the first ancestry's *cis*-SNP heritability of 0.05, 2 causal *cis*-molQTLs, the per-ancestry sample size of 400, and the *cis*-molQTL effect size correlation across ancestries of 0.8. The error bar is a 95% confidence interval.

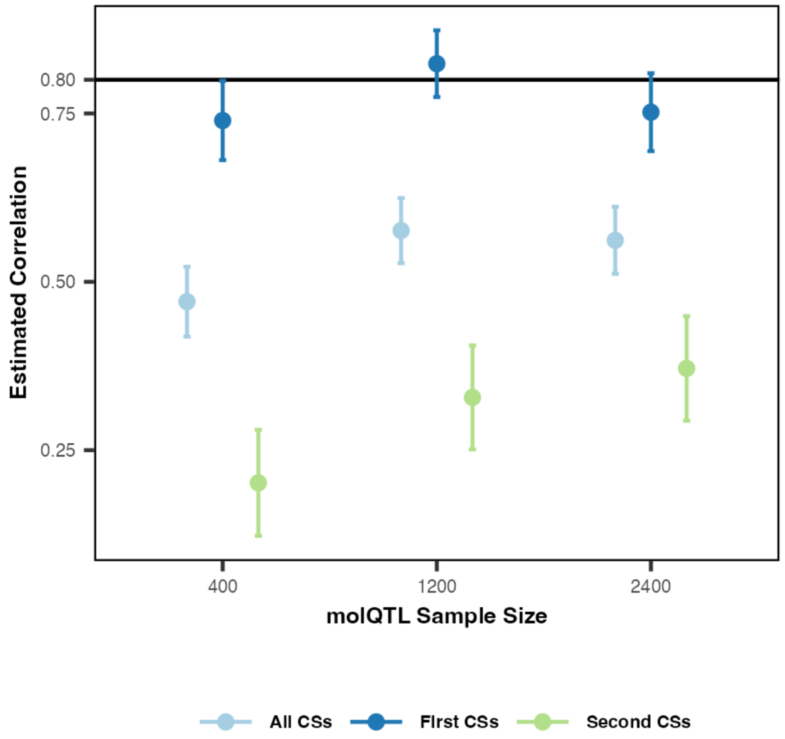


**Fig. S7: The effect-size correlation estimation over the second and all credible sets becomes more accurate as increasing the *cis*-molQTL sample size**

As we increase the per-ancestry sample size (from 400 to 2,400), the estimation using the primary effects (First credible sets) remains robust, and the performance by all and second credible sets gets closer to the true value (0.8). By default, the simulation assumes a per-ancestry sample size of 400, 2 causal *cis*-molQTLs, *cis*-SNP heritability of 0.05, and the *cis*-molQTL effect size correlation across ancestries of 0.8. The error bar is a 95% confidence interval.

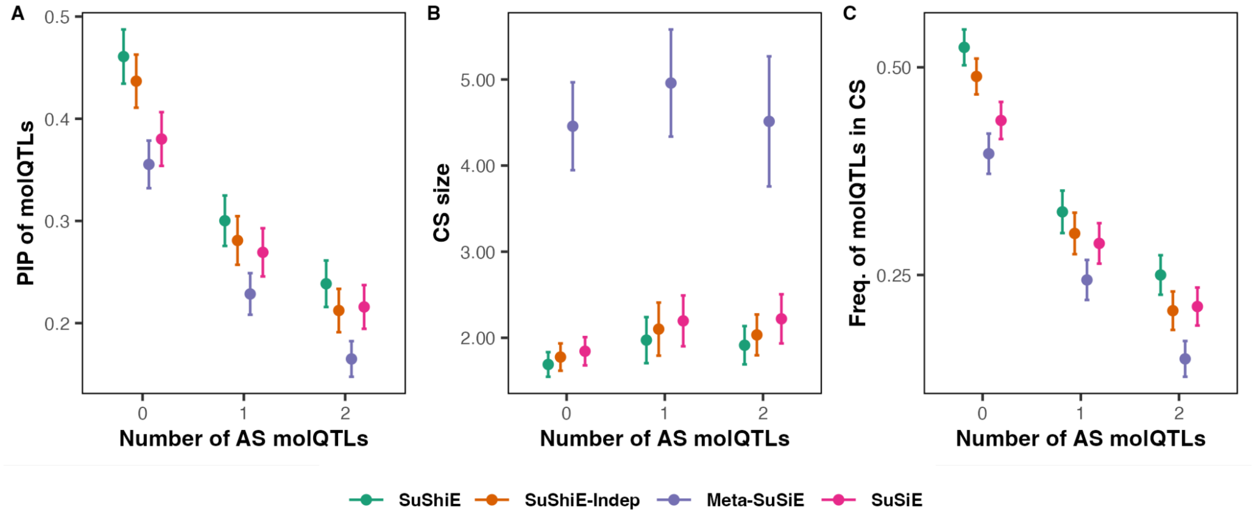


**Fig. S8: SuShiE outputs higher PIPs, smaller credible set size, and better calibration when ancestry-specific *cis*-molQTLs are present**

SuShiE outputs higher posterior inclusion probability (PIPs; A), smaller credible set size (B), and higher frequency of *cis*-molQTLs in the credible set (Calibration; C) compared against SuShiE-Indep (P=8.38e-3, 2.38e-1, and 1.95e-4), Meta-SuSiE (P=3.82e-18, 6.03e-74, and 2.22e-26), and SuSiE (P=1.97e-6, 8.17e-2, and 1.14e-8) when ancestry-specific *cis*-molQTLs are present. The P value is computed across all simulations (see **Methods**). By default, the simulation assumes the first ancestry's *cis*-SNP heritability of 0.05, 2 shared causal *cis*-molQTLs, the per-ancestry sample size of 400, and the *cis*-molQTL effect size correlation across ancestries of 0.8. The error bar is a 95% confidence interval.

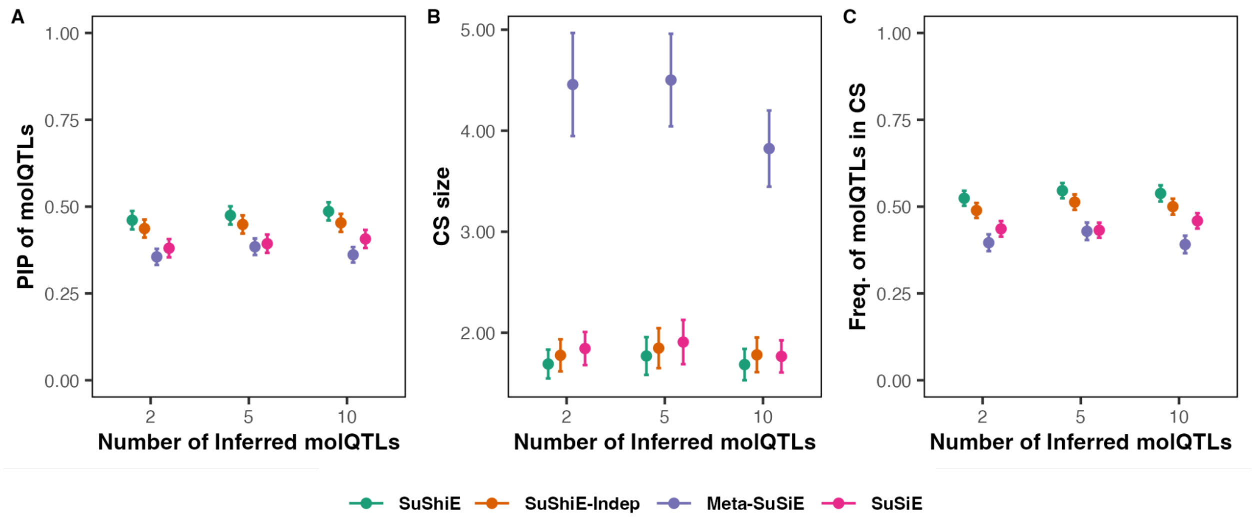


**Fig. S9: SuShiE's out-performance remains when the number of inferred *cis*-molQTLs is greater than the generative *cis*-molQTLs**

SuShiE's out-performance remains the same when the number of inferred *cis*-molQTLs exceeds the generative *cis*-molQTLs. SuShiE outputs higher posterior inclusion probability (PIPs; A), smaller credible set size (B), and higher frequency of *cis*-molQTLs in the credible set (Calibration; C) compared against SuShiE-Indep (P=4.51e-3, 2.23e-1, and 1.77e-3), Meta-SuSiE (P=4.55e-24, 1.18e-105, and 1.09e-26), and SuSiE (P=1.82e-14, 1.37e-1, and 3.26e-15). The P value is computed across all simulations (see **Methods**). By default, the simulation assumes the first ancestry's *cis*-SNP heritability of 0.05, 2 shared causal *cis*-molQTLs, the per-ancestry sample size of 400, and the *cis*-molQTL effect size correlation across ancestries of 0.8. The error bar is a 95% confidence interval.

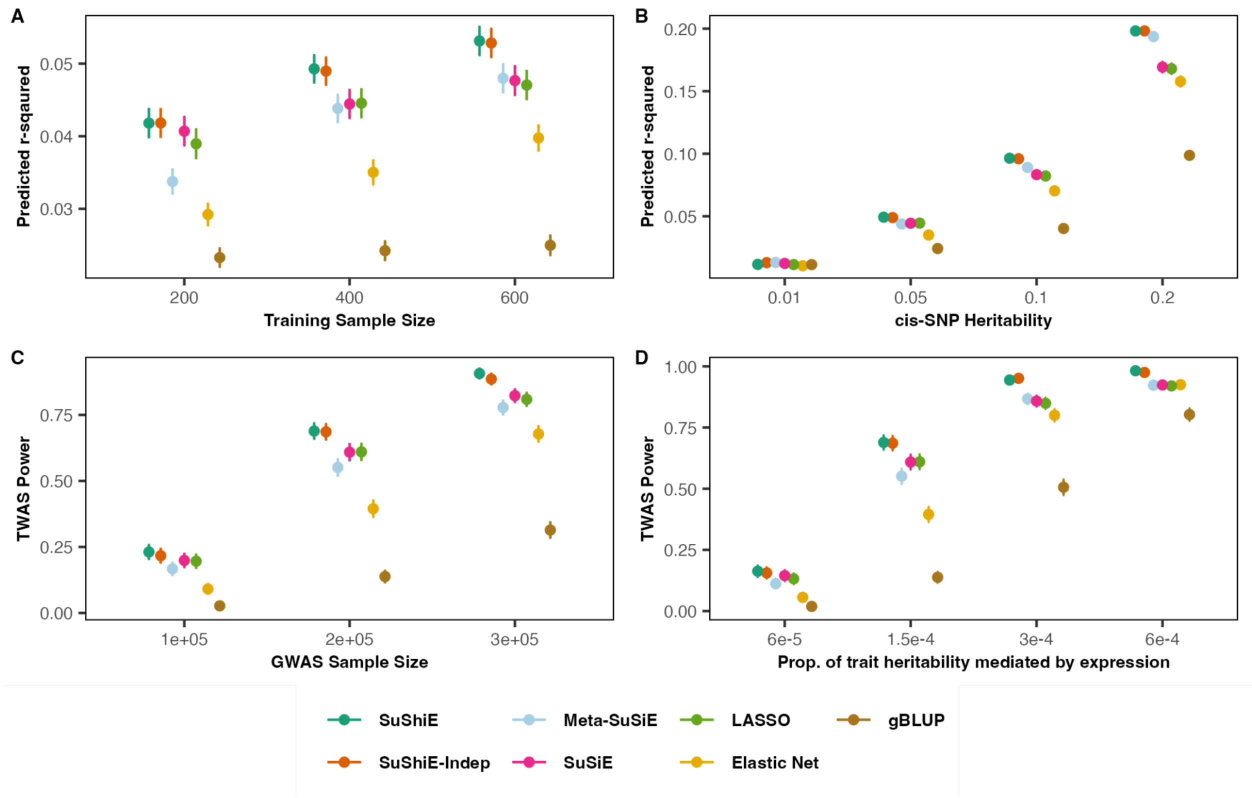


**Fig. S10: The ancestry-specific *cis*-molQTL effect sizes estimated by SuShiE output higher prediction accuracy, thus boosting TWAS power**

**A**) Extension of the main figure 2E. SuShiE outputs higher ancestry-specific prediction accuracy compared to SuShiE-Indep, Meta-SuSiE, SuSiE, LASSO, Elastic Net, and gBLUP (P=4e-1, 3.42e-17, 1.56e-7, 5.32e-10, 1.69e-72, and 5.24e-222) with the fixed sample size.
**B**) SuShiE outputs higher ancestry-specific prediction accuracy compared to Meta-SuSiE, SuSiE, LASSO, Elastic Net, and gBLUP (P=4.77e-6, 2.79e-38, 1.21e-42, 1.45e-115, and <1.45e-115) with the fixed sample size. SuShiE outputs similar prediction accuracy compared to SuShiE-Indep with the fixed sample size (P=4.21).
**C**) Extension of the main figure 2F. SuShiE induces higher TWAS power compared to SuShiE-Indep, Meta-SuSiE, SuSiE, LASSO, Elastic Net, and gBLUP (P=1.93e-1, 6.93e-38, 1.19e13, 8.89e-18, 6.40e-161, and <6.40e-161) with the fixed sample size.
**D**) SuShiE induces higher TWAS power compared to SuShiE-Indep, Meta-SuSiE, SuSiE, LASSO, Elastic Net, and gBLUP (P=1.25e-1, 6.51e-56, 1.06e19, 1.46e-25, 2.01e-226, and <2.01e-226) with the fixed sample size.

The plots are aggregation across two ancestries. The per-ancestry training sample size is 400, and the testing sample size is 200. The simulation assumes 2 causal *cis*-molQTLs, the default *cis*-SNP heritability is 0.05, the *cis*-molQTL effect size correlation is 0.8 across ancestries, and the default proportion of *cis*-SNP heritability of complex trait explained by gene expression is 1.5e-14. The error bar is a 95% confidence interval.

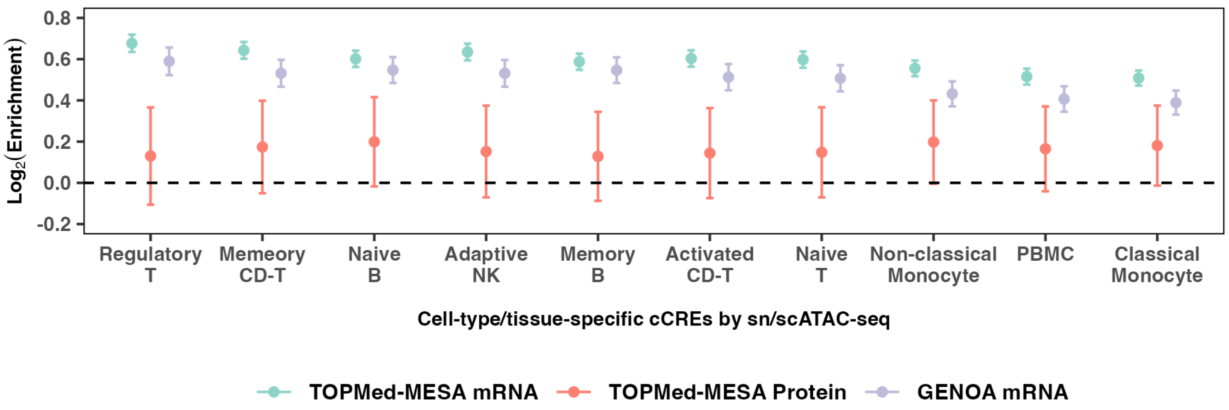


**Fig. S11: SuShiE inferred-PIPs of *cis*-molQTLs are enriched in different cell-type/tissue-specific cCREs by sn/scATAC-seq**

Across all three studies, *cis*-molQTLs' PIPs inferred by SuShiE are enriched in all 10 cell-type/tissue-specific cCREs by sn/sc ATAC-seq. Specifically, the mRNA expression of TOPMed-MESA and GENOA showed significant enrichment but non-significance for the protein expression of TOPMed-MESA because of the low number of genes identified with pQTLs (n=573). PBMC is measured with scATAC-seq in Satpathy et al.[1], while the rest cCREs are measured with snATAC-seq in Chiou et al.[2] The error bar is a 95% confidence interval.

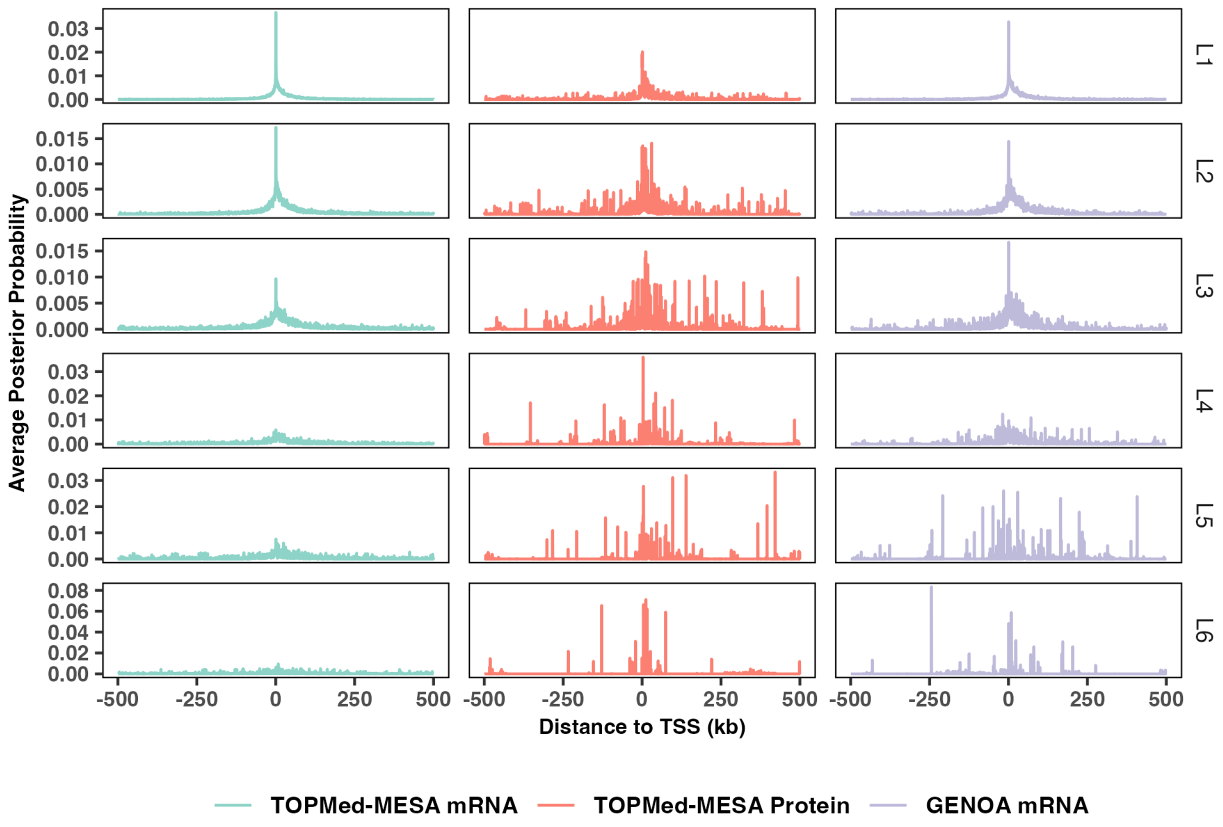


**Fig. S12: The relationship between the posterior probability inferred by SuShiE for shared effect from L1 to L6 and their distance to TSS of each gene**

The posterior probabilities ($\alpha_l$) of *cis*-molQTLs inferred by SuShiE from L1 to L3 are mainly enriched around the TSS region of genes, consistent with the findings that most genes exhibited 1-3 *cis*-molQTLs. We also observed relatively smaller enrichment all over the gene window when expanding to L4, L5, and L6. We grouped SNPs into 500-bp-long bins and computed the average of posterior probability. There are 2,000 bins to cover a one-million-bp-long genomic window around the gene.

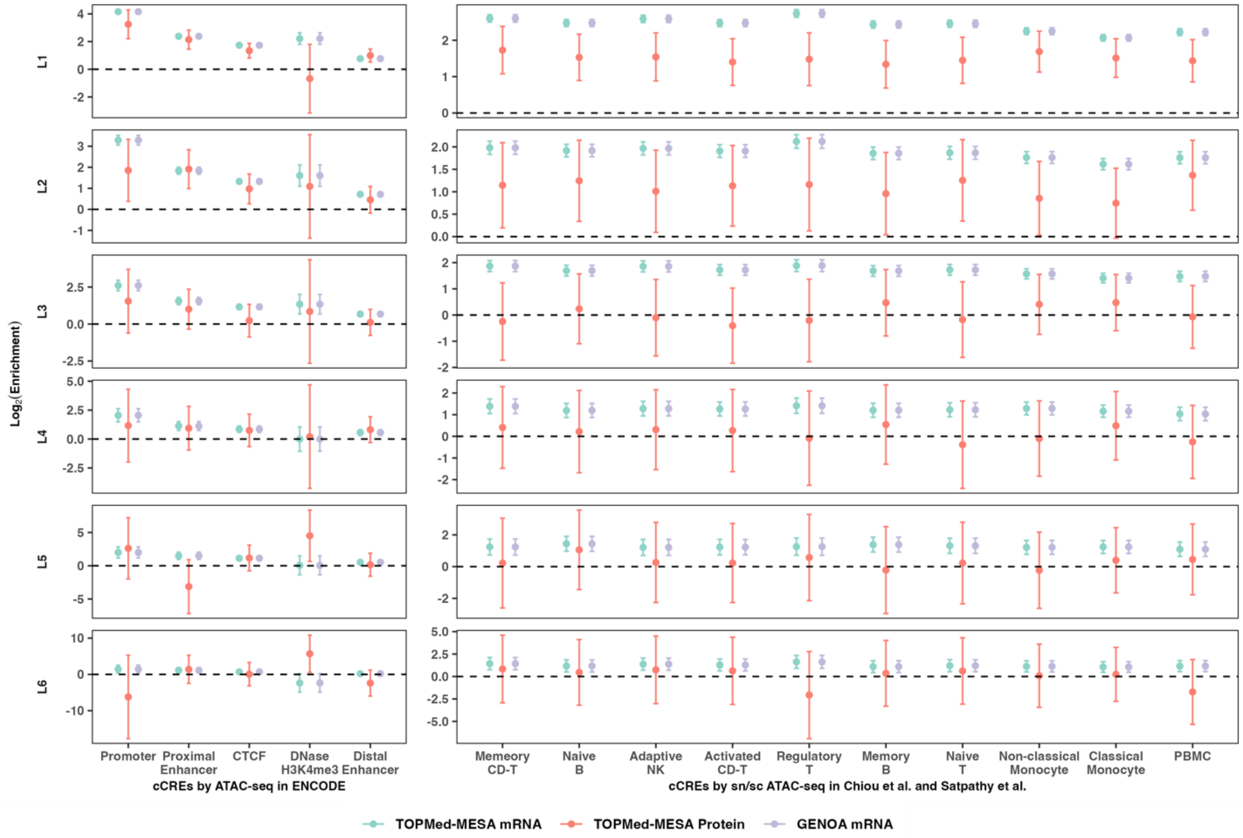


**Fig. S13: The functional enrichment decreased as the proportion of *cis*-molQTLs explained by each shared effect decreased**

The first three effects are enriched in all regulatory annotations with higher enrichment scores compared to using the PIPs (Fig. 3C, 3D). The enrichment decreased from L1 to L6 because the proportion of *cis*-molQTLs explained by each shared effect decreased. The mRNA expression of TOPMed-MESA and GENOA showed significant enrichment in most annotations but non-significance for the protein expression of TOPMed-MESA because of the low number of genes identified. The promoter, proximal enhancer, CTCF, distal enhancer, and DNase-H3K4me3 are measured in ENCODE; PBMC is measured with scATAC-seq in Satpathy et al., and the rest cCREs are measured with snATAC-seq in Chiou et al. The error bar is a 95% confidence interval.

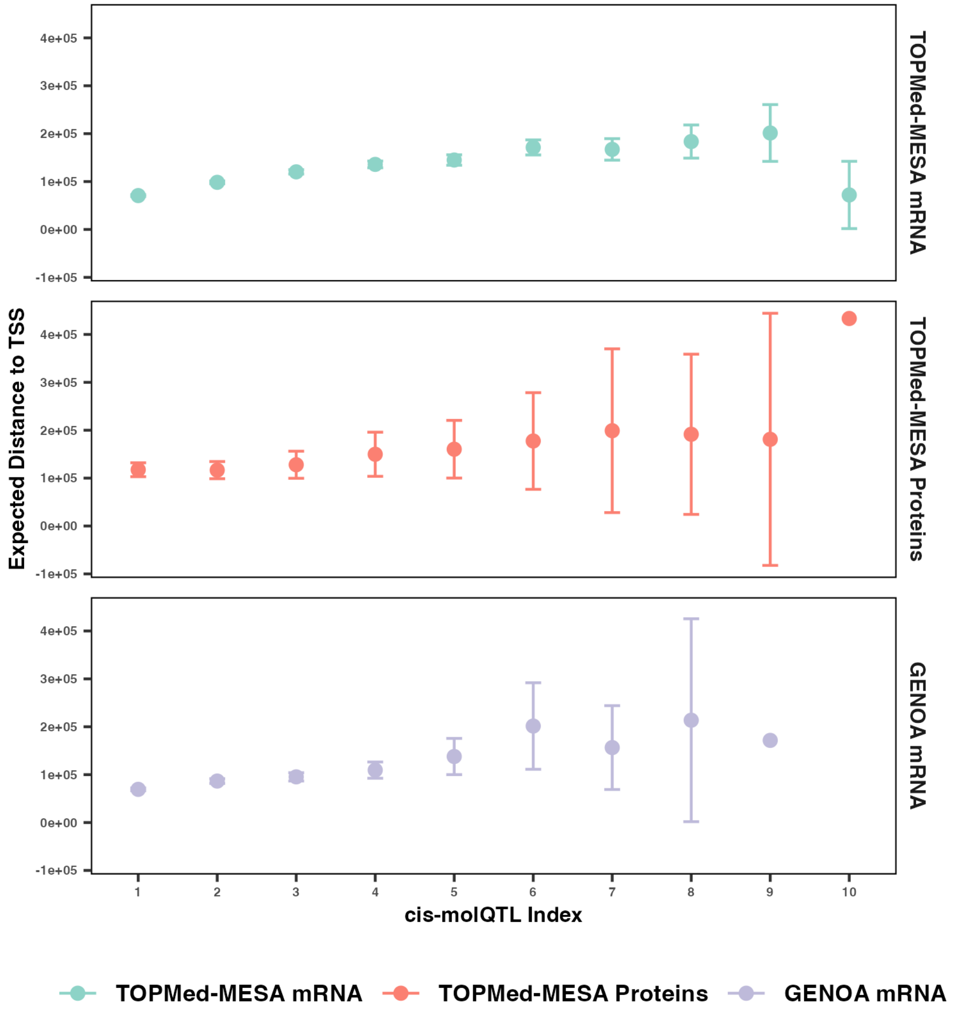


**Fig. S14: The expected distance between *cis*-molQTLs and TSS increased as the proportion of *cis*-molQTLs explained by each shared effect decreased**

We computed the expected distance between cis-molQTLs and TSS (see **Methods**) within each *cis*-molQTL index (L index). For example, we computed the average expected distance of the SNPs to the TSS, who are included in the first credible set (L=1). The x-axis is the corresponding *cis*-molQTL index, suggesting the proportion of *cis*-molQTLs explained from highest to lowest. The error bar is a 95% confidence interval.

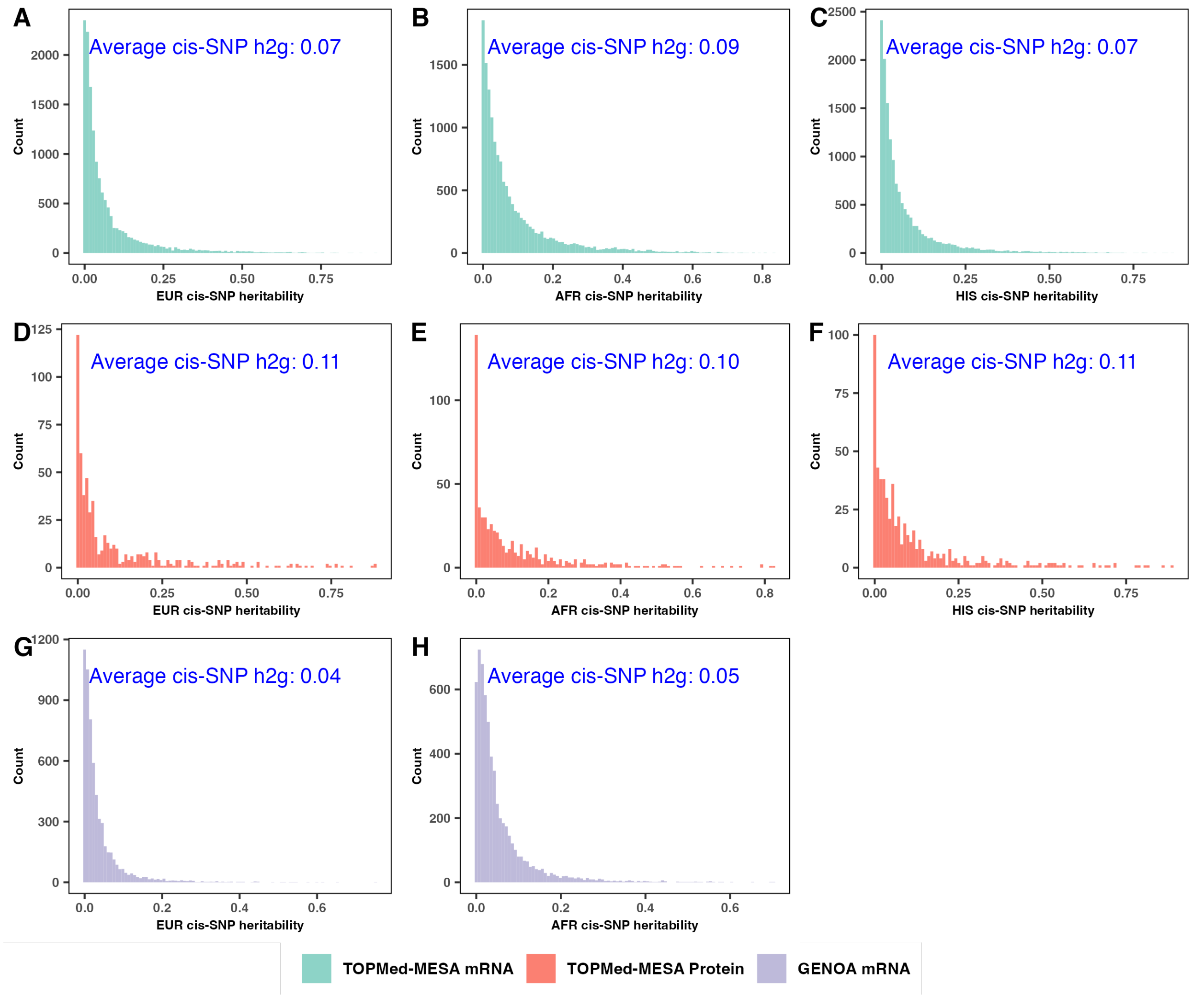


**Fig. S15: The average *cis*-SNP heritability estimates of molecular traits are significantly non-zero for each ancestry and each study.**

We used limix to compute the *cis*-SNP heritability for each ancestry of each e/pGene (h2g; see **Codes Availability**). We observed that for each ancestry of each study, the average h2g is statistically non-zero (P<1e-200 for all cases).

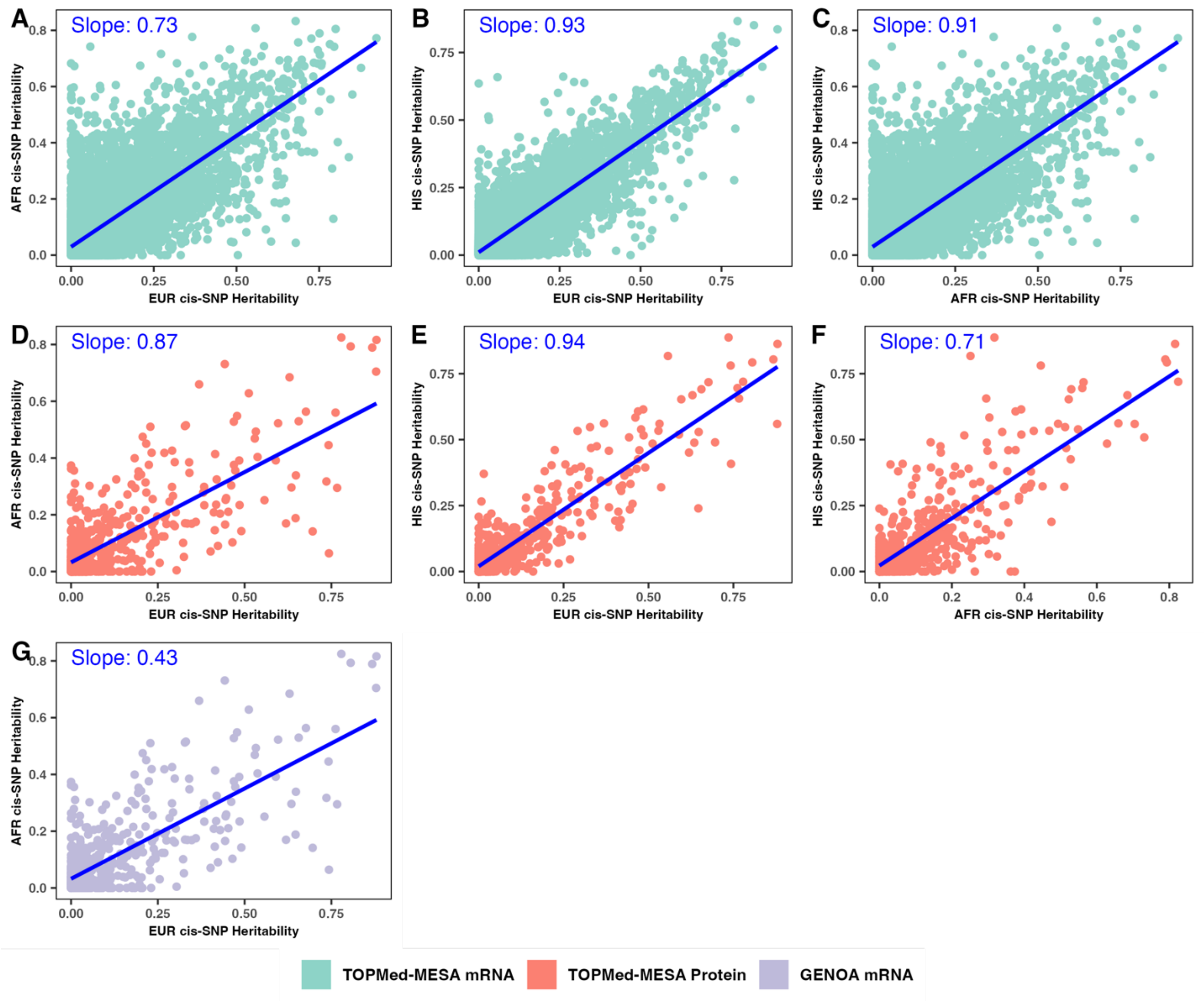


**Fig. S16: The *cis*-SNP heritability estimates of molecular traits are highly correlated across ancestries.**

We used limix to compute the *cis*-SNP heritability for each ancestry of each e/pGene (h2g; see **Codes Availability**). We observed that for each pair of ancestry, the heritabilities' correlations are highly correlated (P<1e-200 for all cases compared to 0).

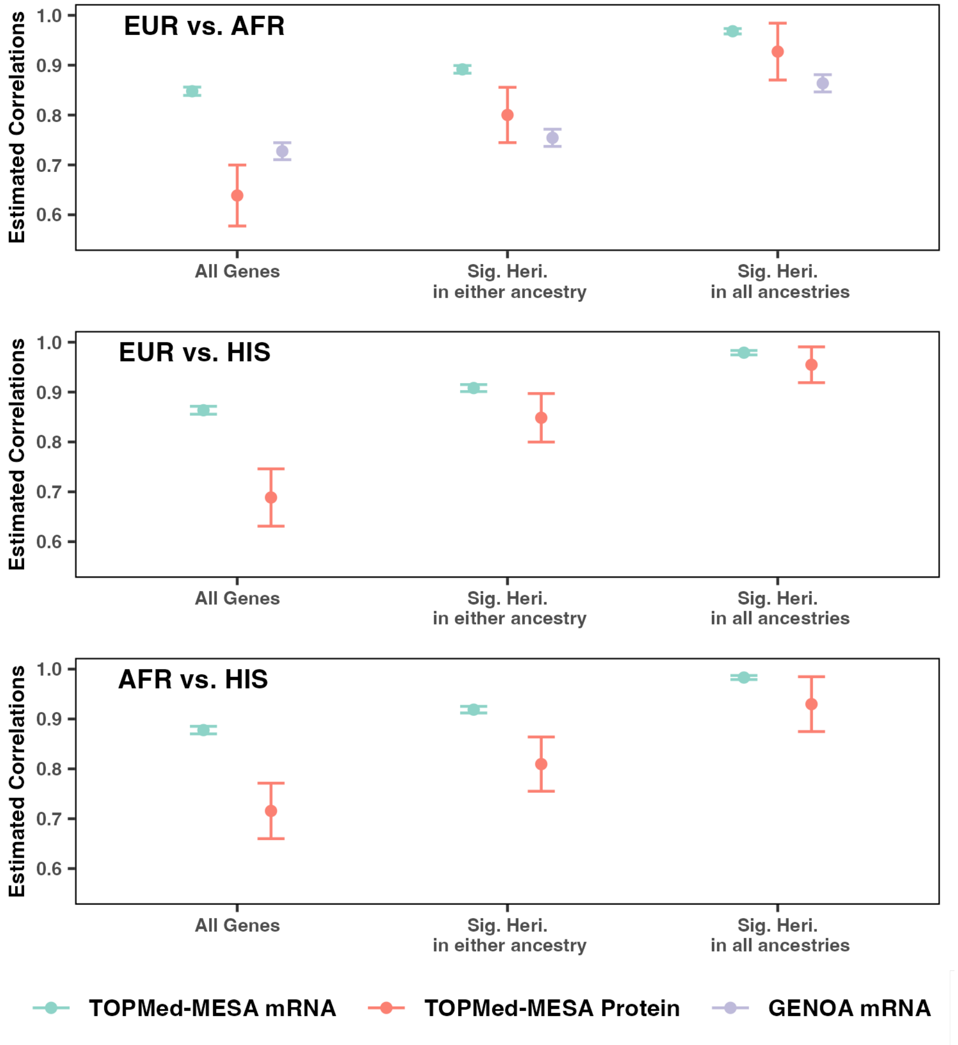


**Fig. S17: SuShiE estimated ancestry-specific effect sizes are highly correlated when focusing on genes that are significantly heritable in all ancestries**

By focusing on the primary shared effect (L1) of all genes, SuShiE estimated *cis*-molQTL average correlations of 0.81, 0.86, and 0.87 for EUR-AFR, EUR-HIS, and AFR-HIS, respectively. When reducing the genes that exhibited significant heritable genes in at least one ancestry, we observed average correlations increased to 0.85, 0.91, and 0.92, and then further increased to 0.94, 0.98, and 0.99 when focusing on genes whose heritabilities are significant in all ancestries. The error bar is a 95% confidence interval.

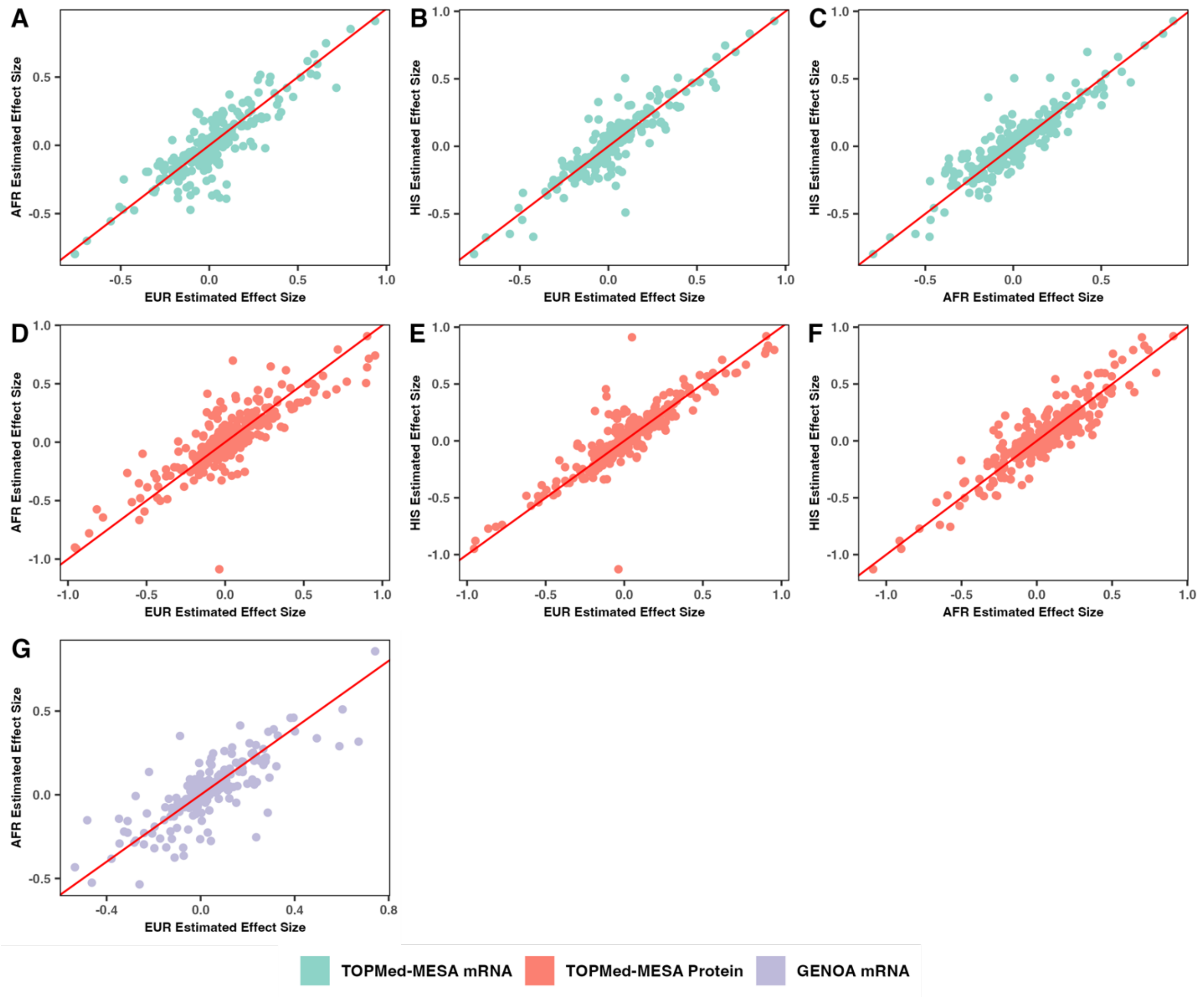


**Fig. S18: The ancestry-specific effect sizes inferred by SuShiE are highly correlated across ancestries**

We randomly picked 100 genes that are significantly heritable in all ancestries and plotted the scatter for their ancestry-specific effect sizes. The red line is the identical line ($y = x$).

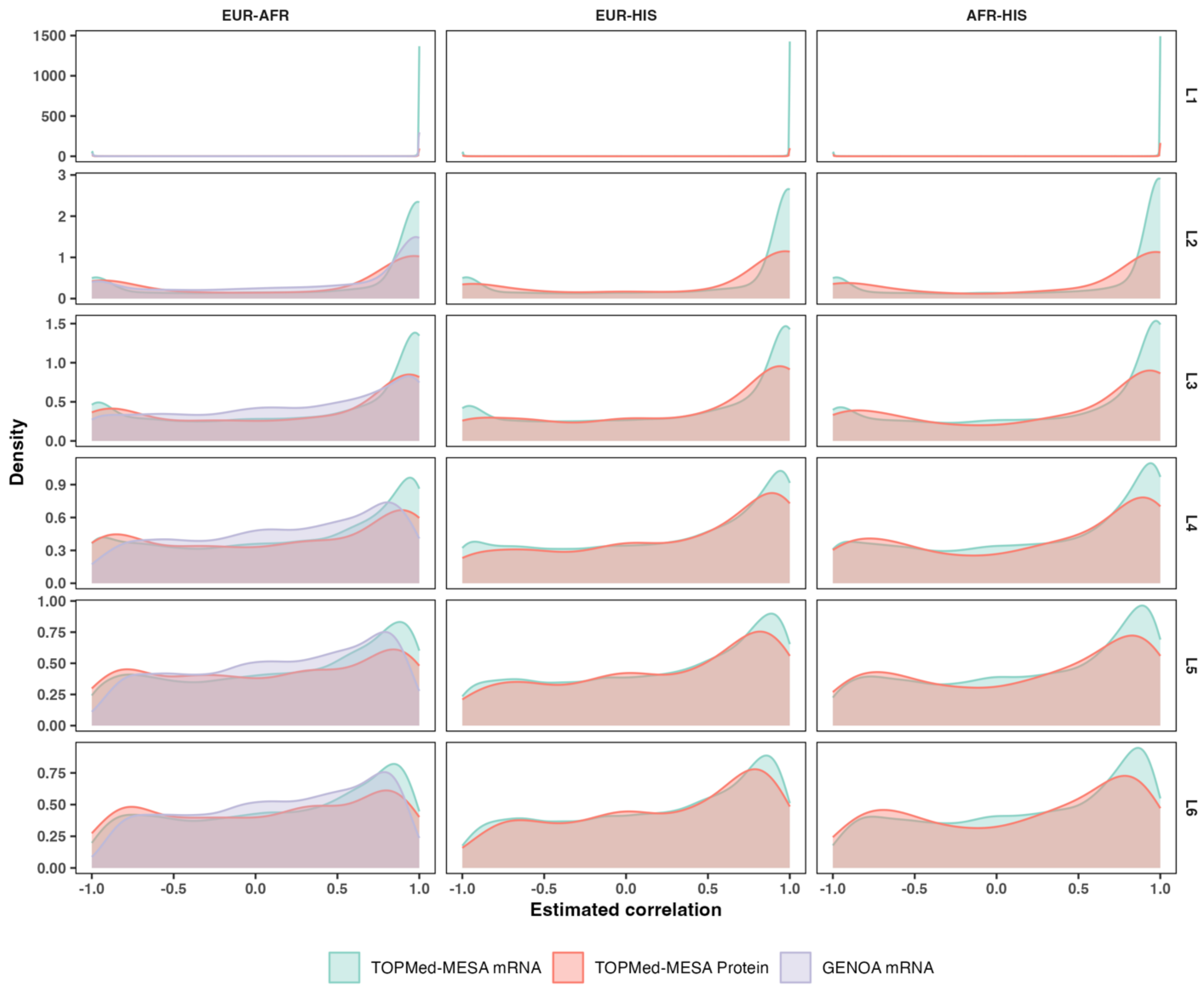


**Fig. S19: The *cis*-molQTL effect size correlations across ancestries estimated by SuShiE can be less than one**

The density plot of the *cis*-molQTL effect size correlations estimated by SuShiE for EUR-AFR, EUR-HIS, and AFR-HIS pairs from TOPMed-MESA mRNA, TOPMed-MESA Protein, and GENOA mRNA datasets. The figures show the first six single shared effects (from L1 to L6). These figures suggest that some molecules can exhibit ancestry-specific effect sizes (i.e., *cis*-molQTL effect size heterogeneity across ancestries).

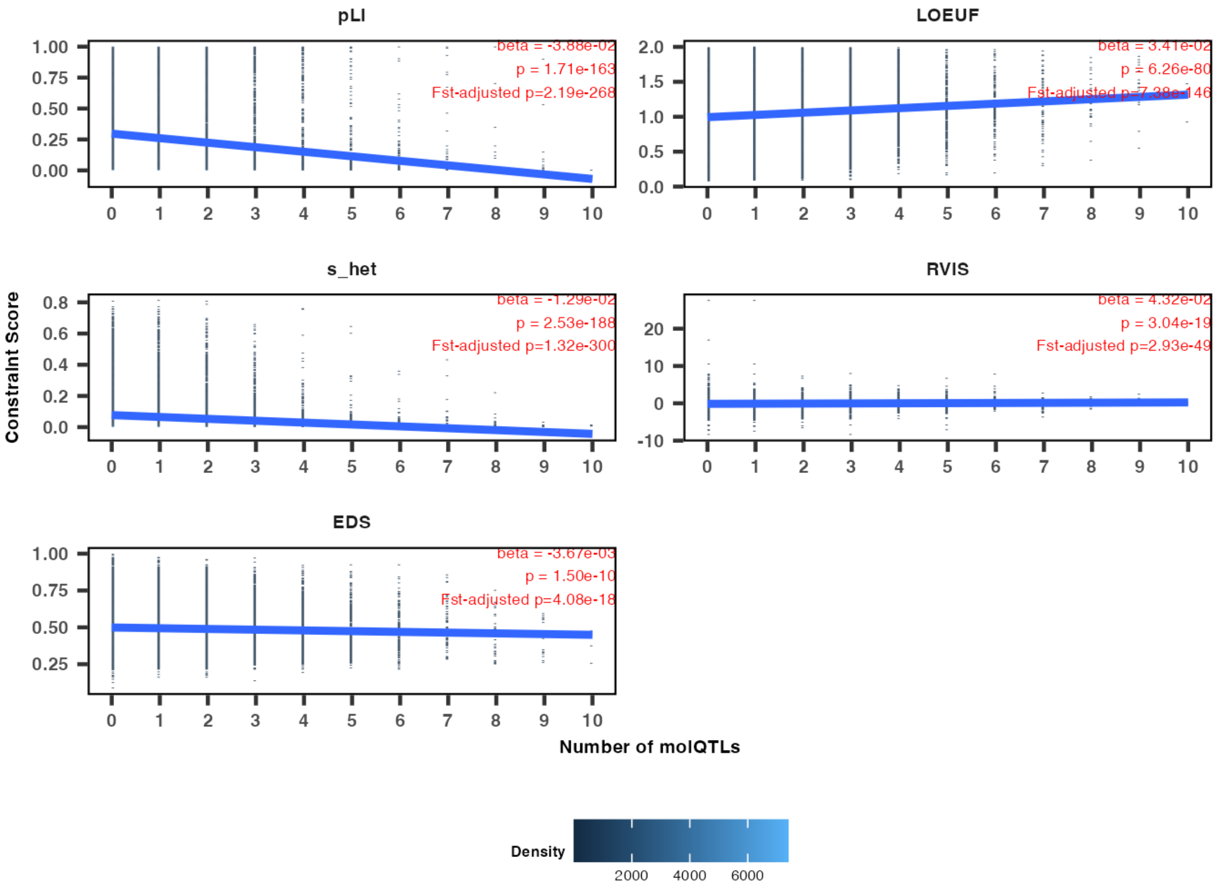


**Fig. S20: Gene constraint scores are negatively associated with their number of *cis*-molQTLs**

The 2d density plot between the number of *cis*-molQTLs (x-axis) and pLI, LOEUF, $s_{het}$, RVIS, and EDS as measures of loss-of-function intolerance (y-axis) across all three studies  (see **Methods**). The blue line is the linear regression estimate. A higher value of pLI, $s_{het}$, and, EDS is taken to indicate stronger constraint, while a lower value of LOEUF and RVIS is suggestive of more constraint. The beta is calculated after adjusting for study (i.e., TOPMed MESA mRNA, TOPMed MESA proteins, and GENOA mRNA). The p-value is calculated using bootstrap and after adjusting for $F_{st}$.

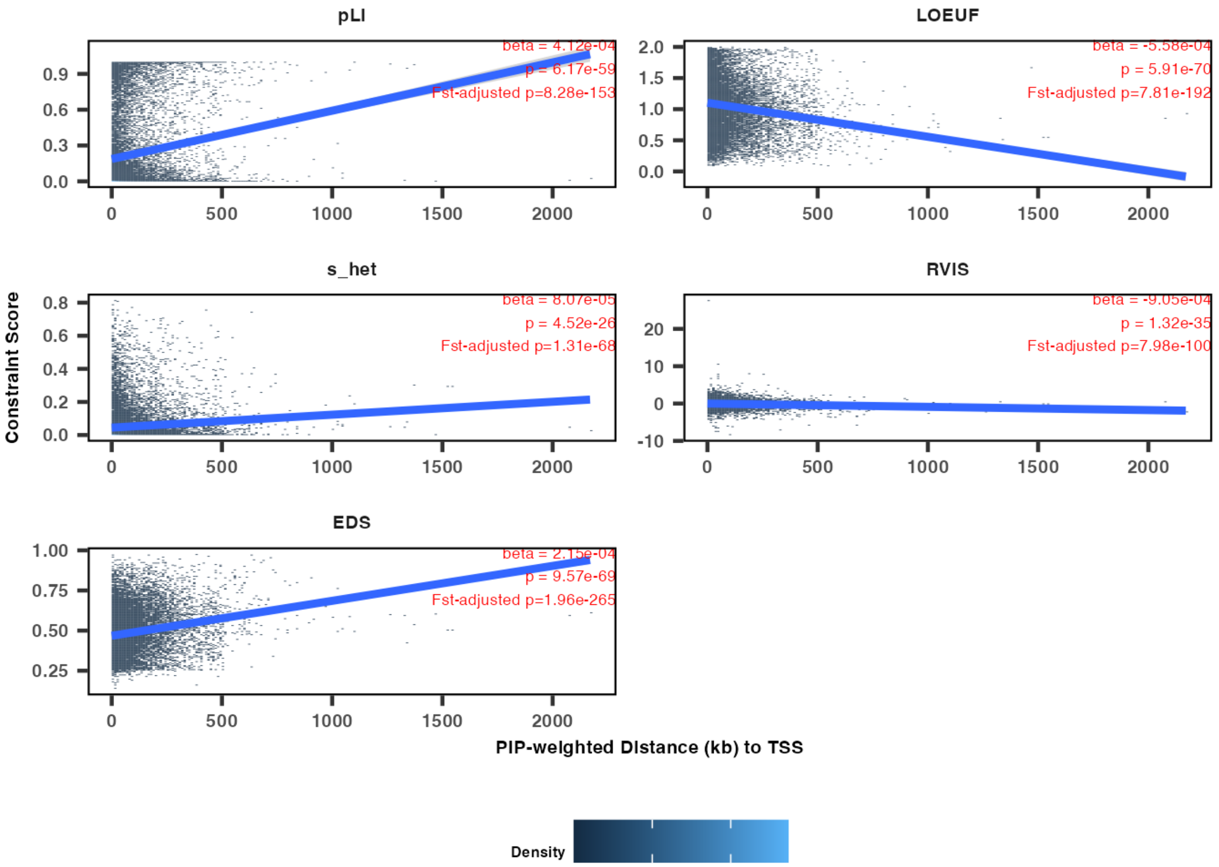


**Fig. S21: Gene constraint scores are positively associated with the expected distance between their *cis*-molQTLs and TSS**

The 2d density plot between *cis*-molQTLs distance to the gene TSS (x-axis) and pLI, LOEUF, $s_{het}$, RVIS, and EDS as measures of loss-of-function intolerance (y-axis) across all three studies (see **Methods**). The blue line is the linear regression estimate. A higher value of pLI, $s_{het}$, and, EDS is taken to indicate stronger constraint, while a lower value of LOEUF and RVIS is suggestive of more constraint. The beta is calculated after adjusting for study (i.e., TOPMed MESA mRNA, TOPMed MESA proteins, and GENOA mRNA). The p-value is calculated using bootstrap and after adjusting for $F_{st}$.

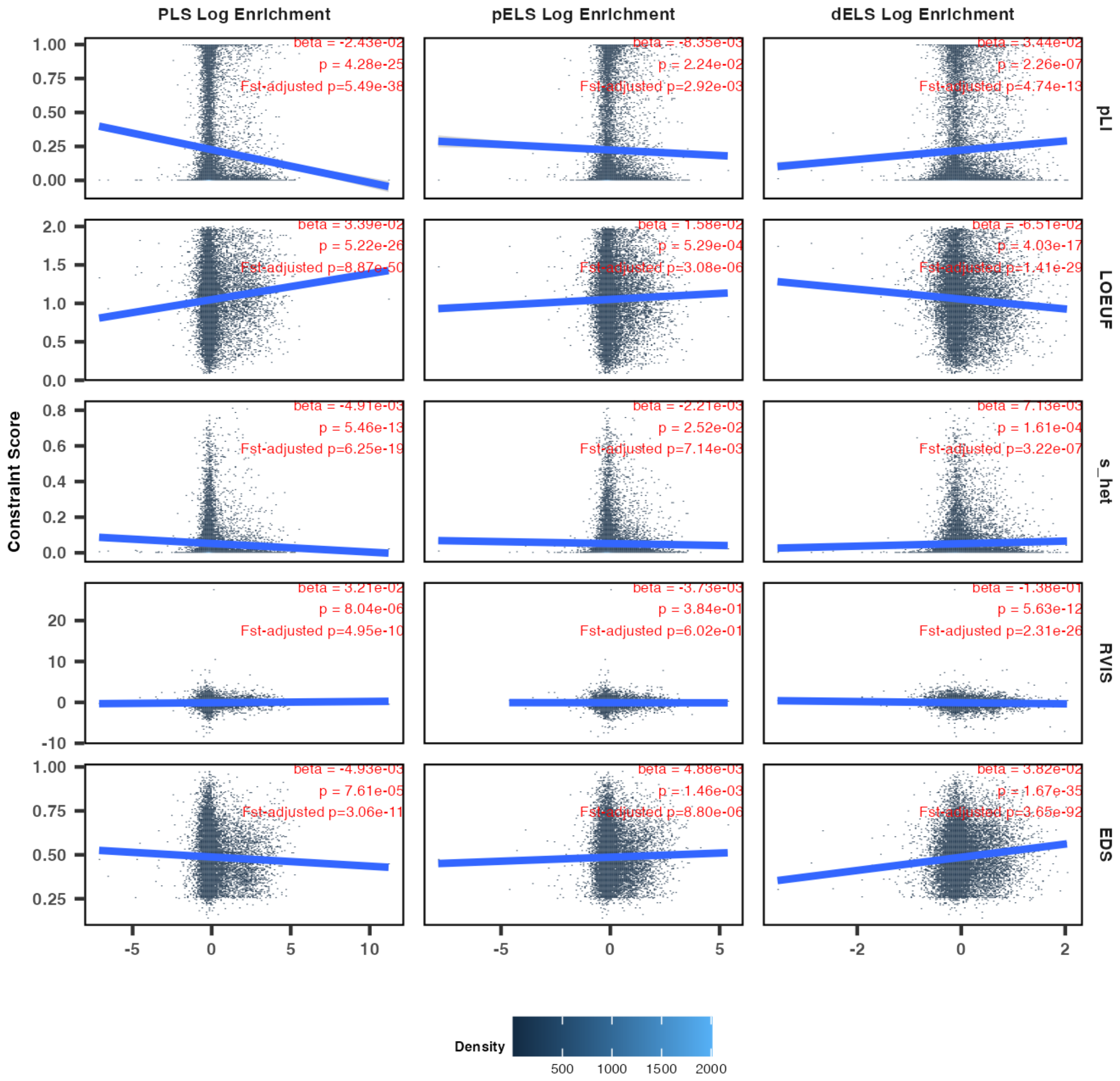


**Fig. S22: Gene constraint scores are associated with their cis-molQTL functional enrichment**

The 2d density plot between functional enrichment estimates calculated using their *cis*-molQTL PIPs (x-axis) and pLI, LOEUF, $s_{het}$, RVIS, and EDS as measures of loss-of-function intolerance (y-axis) across all three studies (see **Methods**). Each column is the promoter (PLS), proximal enhancer (pELS), and distal enhancer (dELS) annotation measured from from ENCODE[3]. The blue line is the linear regression estimate. A higher value of pLI, $s_{het}$, and, EDS is taken to indicate stronger constraint, while a lower value of LOEUF and RVIS is suggestive of more constraint. The beta is calculated after adjusting for study (i.e., TOPMed MESA mRNA, TOPMed MESA proteins, and GENOA mRNA). The p-value is calculated using bootstrap and after adjusting for $F_{st}$. The PLS and pELS enrichments are negatively associated with their gene constraint scores. The dELS enrichment is negatively associated with their gene constraint scores.

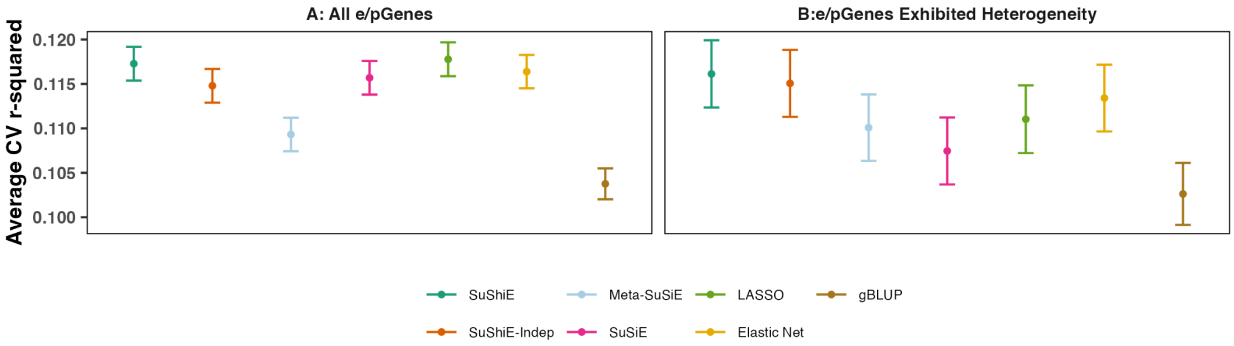


**Fig. S23: SuShiE outputs better or comparable prediction accuracy compared to alternative methods**

Point estimates for the cross-validation coefficient of determination (cv-$r^2$). Across all the e/pGenes (A), SuShiE output higher cv-$r^2$ compared to SuShiE-Indep, Meta-SuSiE, SuSiE, Elastic Net, and gBLUP (P=3.45e-2, 2.67e-9, 1.23e-1, 2.56e-1, and 2.81e-25). SuShiE output comparable cv-$r^2$ relative to LASSO (P=6.43e-1). When focusing on e/pGenes exhibited estimated cis-molQTL effect size correlation < 0.9 (B), SuShiE output higher cv-$r^2$ compared to all methods: SuShiE-Indep, Meta-SuSiE, SuSiE, LASSO, Elastic Net, and gBLUP (P=3.48e-1, 1.28e-2, 7.01e-4, 3.08e-2, 1.58e-1, and 1.23e-7). The reported P value is one-sided. The error bar is a 95% confidence interval.

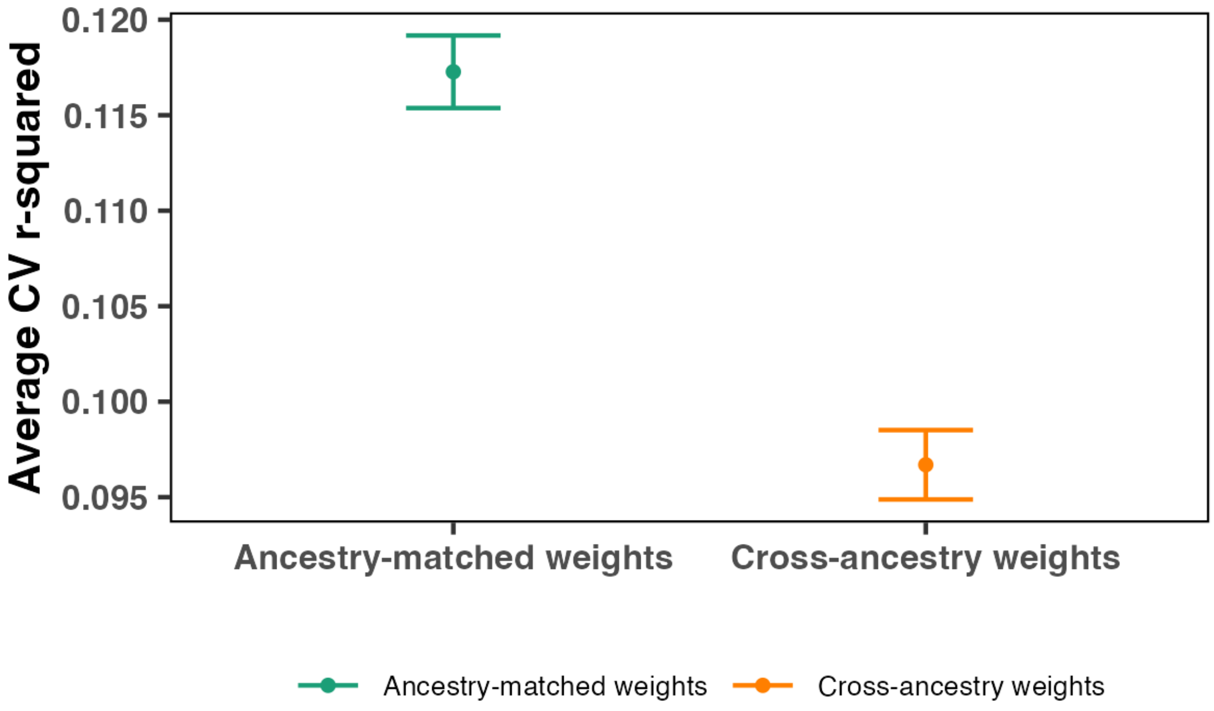


**Fig. S24: Ancestry-matched prediction accuracy is higher than cross-ancestry prediction accuracy**

Point estimates for the cross-validation coefficient of determination (cv-$r^2$). Across all the e/pGenes, we performed cross-ancestry prediction (e.g., predicting mRNA expression of AFR using EUR weights; see **Methods**) using SuShiE-based prediction weights. The ancestry-matched weights (SuShiE-based) produces higher prediction accuracy compared to cross-ancestry prediction (1.71e-53). The reported P value is one-sided. The error bar is a 95% confidence interval.

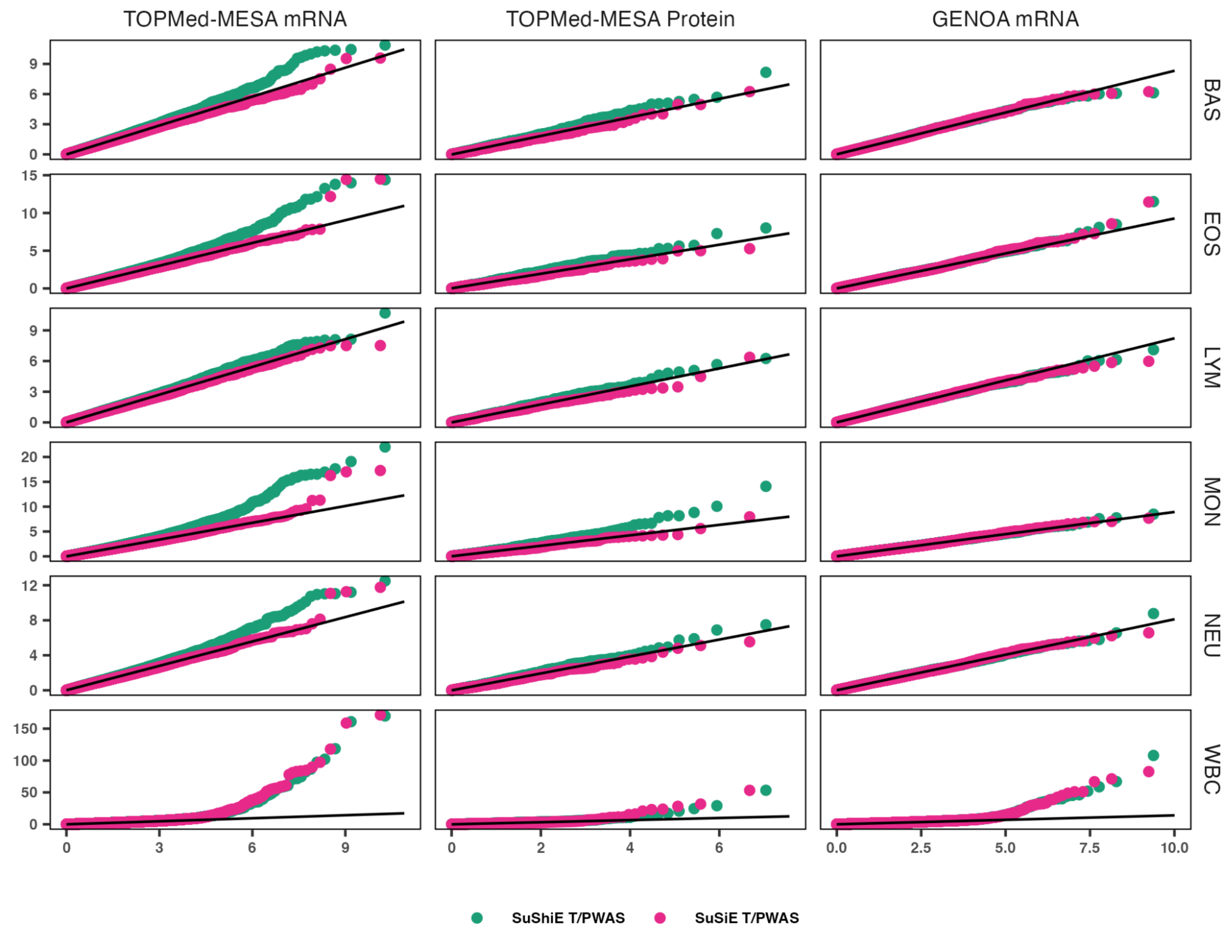


**Fig. S25: The QQ plot of P value for SuShiE and SuSiE-based T/PWAS**

The P value is negative log-transformed and then drawn based on exponential distribution.
