## Supplementary Notes for "Improved multi-ancestry fine-mapping identifies *cis*-regulatory variants underlying molecular traits and disease risk"

### 1. Generative model

Here, we describe the statistical model underlying SuShiE. SuShiE assumes *cis*-molQTLs are present in *all* ancestries (defined as shared *cis*-molQTLs) while allowing for effect sizes at causal *cis*-molQTLs to covary across ancestries a-priori, in contrast to previous multi-ancestry approaches. These assumptions provide enough flexibility to model a variety of *cis*-genetic architectures across ancestries, including cases when effects are present only in a subset of ancestries. For instance, when effects are observed only in a subset of ancestries, prior variances can be shrunk towards zero to effectively allow for *ancestry-specific* causal *cis*-molQTLs.

For the $i^{\text{th}}$ of $k \geq 1$ ancestries, we model the quantitative level of an observed molecular trait $\mathbf{g}_i \in \mathbb{R}^{n_i}$ in $n_i$ individuals as a linear combination of the normalized genotype matrix $\mathbf{X}_i \in \mathsf{Mat}_{n_i \times p}(\mathbb{R})$ and the vector of *cis*-molQTL effects $\boldsymbol{\beta}_i$ at $p$ SNPs according to the following generative model,

$$\mathbf{g}_i = \mathbf{X}_i \boldsymbol{\beta}_i + \boldsymbol{\epsilon}_i \quad (i = 1, \ldots, k)$$

where $\boldsymbol{\epsilon}_i \sim \mathcal{N}(\mathbf{0}, \sigma^2_{e,i}\mathbf{I}_{n_i})$ is environmental noise. To impose sparsity on $\boldsymbol{\beta}_i$ and reflect a *shared* genetic architecture across $\boldsymbol{\beta}_1, \ldots, \boldsymbol{\beta}_k$, we extend the Sum of Single-Effects (i.e., SuSiE) model [1] as,

$$\text{(SuShiE)}\quad \begin{aligned} \boldsymbol{\beta}_i &= \sum_{\ell=1}^{L} \boldsymbol{\beta}_{\ell,i} & (i = 1, \ldots, k) \\ \boldsymbol{\beta}_{\ell,i} &= b_{\ell,i} \cdot \boldsymbol{\gamma}_\ell & (\ell = 1, \ldots, L \text{ and } i = 1 \ldots, k) \\ \boldsymbol{\gamma}_\ell &\overset{\text{iid}}{\sim} \text{Multi}(1, \boldsymbol{\pi}) & (\ell = 1, \ldots, L) \\ \mathbf{b}_\ell = \begin{bmatrix} b_{\ell,1} \\ \vdots \\ b_{\ell,k} \end{bmatrix} &\overset{\text{iid}}{\sim} \mathcal{N}(\mathbf{0}, \mathbf{C}_\ell) & (\ell = 1, \ldots, L), \end{aligned}$$

where $\boldsymbol{\gamma}_\ell$ is a binary $p \times 1$ indicator vector with precisely one non-zero entry specifying which (single) SNP is the shared *cis*-molQTL effect with prior probability $\boldsymbol{\pi}$, the $b_{\ell,i}$ are the *ancestry-specific* effect sizes which, for each fixed $\ell$, we have assumed have a joint distribution $\mathbf{b}_\ell \sim \mathcal{N}(\mathbf{0}, \mathbf{C}_\ell)$. We use the term *ancestry-specific* effect sizes to refer to the effect sizes of the *same* genetic variants for *different* ancestries.

The hyperparameters of the SuShiE model are the prior $k \times k$ covariance matrices $\mathbf{C}_1, \ldots, \mathbf{C}_\ell$, the prior probability for a SNP to be causal (i.e. have a non-zero effect) $\boldsymbol{\pi}$, and the environmental, or residual, variance parameters $\sigma^2_{e,1}, \ldots, \sigma^2_{e,k}$.

Under SuShiE, the complete data likelihood (i.e., joint distribution) has the following form

$$\text{(1)}\quad \begin{aligned} \Pr(\boldsymbol{\beta}, \mathbf{g} \mid \mathbf{X}, \mathbf{C}, \boldsymbol{\pi}, \boldsymbol{\sigma}^2_e) &= \Pr(\mathbf{g} \mid \boldsymbol{\beta}, \mathbf{X}, \mathbf{C}, \boldsymbol{\pi}, \boldsymbol{\sigma}^2_e) \Pr(\boldsymbol{\beta} \mid \mathbf{C}, \boldsymbol{\pi}) \\ &= \left( \prod_{i=1}^{k} \Pr(\mathbf{g}_i \mid \boldsymbol{\beta}_i, \mathbf{X}_i, \sigma^2_{e,i}) \right) \left( \prod_{\ell=1}^{L} \Pr(\boldsymbol{\beta}_{\ell,1}, \ldots, \boldsymbol{\beta}_{\ell,k} \mid \mathbf{C}_\ell, \boldsymbol{\pi}) \right) \\ &= \left( \prod_{i=1}^{k} \mathcal{N}(\mathbf{g}_i \mid \mathbf{X}_i \boldsymbol{\beta}_i, \sigma^2_{e,i}\mathbf{I}_{n_i}) \right) \left( \prod_{\ell=1}^{L} \mathcal{N}(\mathbf{b}_\ell \mid \mathbf{0}, \mathbf{C}_\ell) \text{Multi}(\boldsymbol{\gamma}_\ell \mid 1, \boldsymbol{\pi}) \right), \end{aligned}$$

where we have overloaded notation to reflect $\mathbf{g} = \{\mathbf{g}_1, \ldots, \mathbf{g}_k\}$, $\mathbf{X} = \{\mathbf{X}_1, \ldots, \mathbf{X}_k\}$, $\boldsymbol{\beta} = \{\boldsymbol{\beta}_1, \ldots, \boldsymbol{\beta}_k\}$, $\boldsymbol{\sigma}^2_e = \{\sigma^2_{e,1}, \ldots, \sigma^2_{e,k}\}$, and $\mathbf{C} = \{\mathbf{C}_1, \ldots, \mathbf{C}_\ell\}$. Similarly, the complete-data *log*-likelihood is,

$$\text{(2)}\quad \begin{aligned} \ell(\boldsymbol{\beta}, \mathbf{g} \mid \mathbf{X}, \mathbf{C}, \boldsymbol{\pi}, \boldsymbol{\sigma}^2_e) &= \log \Pr(\boldsymbol{\beta}, \mathbf{g} \mid \mathbf{X}, \mathbf{C}, \boldsymbol{\pi}, \boldsymbol{\sigma}^2_e) \\ &= \sum_{i=1}^{k} \log \mathcal{N}(\mathbf{g}_i \mid \mathbf{X}_i \boldsymbol{\beta}_i, \sigma^2_{e,i}\mathbf{I}_{n_i}) + \sum_{\ell=1}^{L} \log \mathcal{N}(\mathbf{b}_\ell \mid \mathbf{0}, \mathbf{C}_\ell) + \sum_{\ell=1}^{L} \log \text{Multi}(\boldsymbol{\gamma}_\ell \mid 1, \boldsymbol{\pi}). \end{aligned}$$

### 2. Variational Inference

To infer the *cis*-molQTL effects under SuShiE, we aim to infer the posterior distribution $\Pr(\boldsymbol{\beta} \mid \mathbf{g}, \mathbf{X}, \mathbf{C}, \boldsymbol{\pi}, \boldsymbol{\sigma}^2_e)$. This problem is, as stated, intractable due to non-conjugacy with the prior distributions. Instead, we will approach this problem through variational Bayesian methods [2].

Briefly, the goal of variational Bayesian methods is to find an approximating posterior distribution $Q(\boldsymbol{\beta})$ which minimizes a statistical divergence (analogous to a distance) between it and the posterior $\Pr(\boldsymbol{\beta} \mid \mathbf{g}, \mathbf{X}, \mathbf{C}, \boldsymbol{\pi}, \boldsymbol{\sigma}^2_e)$. As is common, we employ the Kullback-Leibler divergence and, in doing so, we can compute[1]

$$\begin{aligned} D_{\mathsf{KL}} \left[ Q(\boldsymbol{\beta}) \middle\| \Pr(\boldsymbol{\beta} \mid \mathbf{g}, \mathbf{X}, \mathbf{C}, \boldsymbol{\pi}, \boldsymbol{\sigma}^2_e) \right] &= \mathbb{E}_Q \left[ \log \frac{Q(\boldsymbol{\beta})}{\Pr(\boldsymbol{\beta} \mid \mathbf{g}, \mathbf{X}, \mathbf{C}, \boldsymbol{\pi}, \boldsymbol{\sigma}^2_e)} \right] \\ &= \mathbb{E}_Q \left[ \log \frac{Q(\boldsymbol{\beta})}{\Pr(\boldsymbol{\beta}, \mathbf{g} \mid \mathbf{X}, \mathbf{C}, \boldsymbol{\pi}, \boldsymbol{\sigma}^2_e)} + \log \Pr(\mathbf{g} \mid \mathbf{X}, \mathbf{C}, \boldsymbol{\pi}, \boldsymbol{\sigma}^2_e) \right] \\ &= \mathbb{E}_Q \left[ \log \frac{Q(\boldsymbol{\beta})}{\Pr(\boldsymbol{\beta}, \mathbf{g} \mid \mathbf{X}, \mathbf{C}, \boldsymbol{\pi}, \boldsymbol{\sigma}^2_e)} \right] + \log \Pr(\mathbf{g} \mid \mathbf{X}, \mathbf{C}, \boldsymbol{\pi}, \boldsymbol{\sigma}^2_e), \end{aligned}$$

with the last equation holding because the marginal log-likelihood does not depend upon $\boldsymbol{\beta}$. Rearranging, we have

$$\begin{aligned} &\log \Pr(\mathbf{g} \mid \mathbf{X}, \mathbf{C}, \boldsymbol{\pi}, \boldsymbol{\sigma}^2_e) \\ &= D_{\mathsf{KL}} \left[ Q(\boldsymbol{\beta}) \middle\| \Pr(\boldsymbol{\beta} \mid \mathbf{g}, \mathbf{X}, \mathbf{C}, \boldsymbol{\pi}, \boldsymbol{\sigma}^2_e) \right] + \underbrace{\mathbb{E}_Q \left[ \log \frac{\Pr(\boldsymbol{\beta}, \mathbf{g} \mid \mathbf{X}, \mathbf{C}, \boldsymbol{\pi}, \boldsymbol{\sigma}^2_e)}{Q(\boldsymbol{\beta})} \right]}_{\mathsf{ELBO}}, \end{aligned}$$

[1] While slightly unwieldy, we denote $\mathbb{E}_Q[\cdot], \mathbb{V}_Q[\cdot]$ to be the expectation and variance with respect to $\boldsymbol{\beta}$ under the approximating distribution $Q$ to make explicit what we consider random, and under what sampling assumptions expectations are derived.

where the second term is the evidence lower bound (ELBO) and depends upon the variational distribution $Q(\boldsymbol{\beta})$. The ELBO has earned this name because the KL-divergence is always non-negative, so that the inequality

$$\log \Pr(\mathbf{g} \mid \mathbf{X}, \mathbf{C}, \boldsymbol{\pi}, \boldsymbol{\sigma}_e^2) \geq \mathsf{ELBO}$$

holds.

Notice, furthermore, that the sum of the ELBO with the KL-divergence between $Q$ and $P$ is equal to the marginal log-likelihood, independent of the form of the approximating variational distribution $Q$. This constraint implies that maximizing the ELBO is equivalent to minimizing $D_{\mathsf{KL}}[Q\|P]$. Maximizing the ELBO will now be our goal.

In order to limit the universe of possible forms that $Q$ may take, we impose an additional mean-field assumption[2]. Namely, as in the SuSiE model, the SuShiE model assumes that each of the $L$ *shared* effects $\boldsymbol{\beta}_\ell$ are mutually independent under $Q$. Formally, this implies

$$Q(\boldsymbol{\beta}_1, \ldots, \boldsymbol{\beta}_k) = \prod_{\ell=1}^{L} Q(\boldsymbol{\beta}_\ell) = \prod_{\ell=1}^{L} Q(\mathbf{b}_\ell, \gamma_\ell), \tag{3}$$

where $\boldsymbol{\beta}_\ell$, as an argument of the distributions in equation (3), must be interpreted as the pair $(\mathbf{b}_\ell, \gamma_\ell)$.

### 3. Optimal Update Form and Variational Parameters

While equation (3) specifies the structural assumptions underlying $Q$, we have yet to identify the functional form and associated parameters required for inference. Here, we derive the *optimal* form for $Q$ and identify its variational parameters using coordinate ascent variational inference (i.e., CAVI). See the review [2] for more details. The reader should take note, however, that we will denote by $\mathbb{E}_Q\left[\ell(\boldsymbol{\beta}, \mathbf{g} \mid \mathbf{X}, \mathbf{C}, \boldsymbol{\pi}, \boldsymbol{\sigma}_e^2) \mid \mathbf{b}_\ell, \gamma_\ell\right]$ the expectation $\mathbb{E}_{\neg\ell}\left[\ell(\boldsymbol{\beta}, \mathbf{g} \mid \mathbf{X}, \mathbf{C}, \boldsymbol{\pi}, \boldsymbol{\sigma}_e^2)\right]$ of equation 18 in [2].

To facilitate the computation, we will adopt the following notation. Note that, below, by diag we will mean the *block* diagonal matrix formed by, and ordered as in, the matrices of its argument.

$$\begin{aligned}
n &= \sum_{i=1}^{k} n_i \\
\mathbf{r}_\ell &= \begin{bmatrix} \mathbf{g}_1 - \mathbf{X}_1\left(\sum_{\ell' \neq \ell} \boldsymbol{\beta}_{\ell',1}\right) \\ \vdots \\ \mathbf{g}_k - \mathbf{X}_k\left(\sum_{\ell' \neq \ell} \boldsymbol{\beta}_{\ell',k}\right) \end{bmatrix} \in \mathbb{R}^n \\
\bar{\mathbf{r}}_\ell &= \mathbb{E}_Q[\mathbf{r}_\ell] \\
\mathbf{Z}_j &= \operatorname{diag}((\mathbf{X}_1)_j, \ldots, (\mathbf{X}_k)_j) \in \mathsf{Mat}_{n \times k}(\mathbb{R}) \\
\mathbf{D} &= \operatorname{diag}(\sigma_{e,1}^2, \ldots, \sigma_{e,k}^2) \in \mathsf{Mat}_{k \times k}(\mathbb{R}) \\
\mathbf{g} &= \begin{bmatrix} \mathbf{g}_1 \\ \vdots \\ \mathbf{g}_k \end{bmatrix} \\
\boldsymbol{\beta}_{i \mid \gamma_{\ell,j}=1} &= b_{\ell,i}\mathbf{e}_j + \sum_{\ell' \neq \ell} \boldsymbol{\beta}_{\ell',i},
\end{aligned}$$

where $(\mathbf{X}_i)_j$ is the column vector at the $j^{\text{th}}$ SNP of the $i^{\text{th}}$ ancestry and $\mathbf{e}_j$ is the standard basis vector having as its only non-zero entry the value 1 in the $j^{\text{th}}$ position with its dimension understood by context.

We are now prepared to begin the computation. Whenever possible, we will *first* integrate over the discrete variables $\gamma_{\ell,j}$. We will absorb constants which are *independent* of the value of $\mathbf{b}_\ell$ and each $\gamma_{\ell,j}$ into a term $O(1)$. Using equation (2), we find

$$\begin{aligned}
\log Q(\mathbf{b}_\ell, \gamma_\ell) &= \mathbb{E}_Q\left[\ell(\boldsymbol{\beta}, \mathbf{g} \mid \mathbf{X}, \mathbf{C}, \boldsymbol{\pi}, \boldsymbol{\sigma}_e^2) \,\middle|\, \mathbf{b}_\ell, \gamma_\ell\right] + O(1) \\
&= \mathbb{E}_Q\left[\sum_{i=1}^{k} \log \mathcal{N}(\mathbf{g}_i \mid \mathbf{X}_i\boldsymbol{\beta}_i, \sigma_{e,i}^2 \mathbf{I}_{n_i}) + \sum_{\ell'=1}^{L} \log \mathcal{N}(\mathbf{b}_{\ell'} \mid \mathbf{0}, \mathbf{C}_{\ell'}) \,\middle|\, \mathbf{b}_\ell, \gamma_\ell\right] + \log \operatorname{Multi}(\gamma_\ell \mid 1, \boldsymbol{\pi}) + O(1) \\
&= \sum_{i=1}^{k} \mathbb{E}_Q\left[\log \mathcal{N}(\mathbf{g}_i \mid \mathbf{X}_i\boldsymbol{\beta}_i, \sigma_{e,i}^2 \mathbf{I}_{n_i}) \,\middle|\, \mathbf{b}_\ell, \gamma_\ell\right] - \frac{1}{2}\mathbf{b}_\ell^{\mathsf{T}}\mathbf{C}_\ell^{-1}\mathbf{b}_\ell + \log \operatorname{Multi}(\gamma_\ell \mid 1, \boldsymbol{\pi}) + O(1) \\
n &= \sum_{i=1}^{k} -\frac{1}{2\sigma_{e,i}^2}\mathbb{E}_Q\left[(\mathbf{g}_i - \mathbf{X}_i\boldsymbol{\beta}_i)^{\mathsf{T}}(\mathbf{g}_i - \mathbf{X}_i\boldsymbol{\beta}_i) \mid \mathbf{b}_\ell, \gamma_\ell\right] - \frac{1}{2}\mathbf{b}_\ell^{\mathsf{T}}\mathbf{C}_\ell^{-1}\mathbf{b}_\ell + \log \operatorname{Multi}(\gamma_\ell \mid 1, \boldsymbol{\pi}) + O(1)
\end{aligned}$$

$$= \underbrace{\sum_{i=1}^{k} -\frac{1}{2\sigma_{e,i}^2}\mathbb{E}_Q\left[(\mathbf{g}_i - \mathbf{X}_i\boldsymbol{\beta}_i)^\mathsf{T}(\mathbf{g}_i - \mathbf{X}_i\boldsymbol{\beta}_i) \mid \mathbf{b}_\ell, \gamma_\ell\right]}_{(*)} - \frac{1}{2}\mathbf{b}_\ell^\mathsf{T}\mathbf{C}_\ell^{-1}\mathbf{b}_\ell + \sum_{j=1}^{p}\gamma_{\ell,j}\log(\pi_j) + O(1).$$

At this point, only $(*)$ remains to be analyzed—towards this end, we may exploit discreteness of $\gamma_\ell$. Discreteness of $\gamma_\ell$ implies that $\sum_{j=1}^{p}\gamma_{\ell,j} \equiv 1$ is identically 1, and so

$$(\mathbf{g}_i - \mathbf{X}_i\boldsymbol{\beta}_i)^\mathsf{T}(\mathbf{g}_i - \mathbf{X}_i\boldsymbol{\beta}_i) = \sum_{j=1}^{p}\gamma_{\ell,j}(\mathbf{g}_i - \mathbf{X}_i\boldsymbol{\beta}_i)^\mathsf{T}(\mathbf{g}_i - \mathbf{X}_i\boldsymbol{\beta}_i),$$

but, on the other hand, if $\gamma_{\ell,j} = 1$, we may write

$$\mathbf{g}_i - \mathbf{X}_i\boldsymbol{\beta}_{i\,|\,\gamma_{\ell,j}=1} = \mathbf{r}_{\ell,i} - b_{\ell,i}(\mathbf{X}_i)_j$$

and therefore

$$(\mathbf{g}_i - \mathbf{X}_i\boldsymbol{\beta}_{i\,|\,\gamma_{\ell,j}=1})^\mathsf{T}(\mathbf{g}_i - \mathbf{X}_i\boldsymbol{\beta}_{i\,|\,\gamma_{\ell,j}=1}) = \mathbf{r}_{\ell,i}^\mathsf{T}\mathbf{r}_{\ell,i} - 2\mathbf{r}_{\ell,i}^\mathsf{T}b_{\ell,i}(\mathbf{X}_i)_j + b_{\ell,i}^2(\mathbf{X}_i)_j^\mathsf{T}(\mathbf{X}_i)_j.$$

Finally, observe that

$$\begin{aligned}\mathbb{E}_Q\left[\gamma_{\ell,j}(\mathbf{g}_i - \mathbf{X}_i\boldsymbol{\beta}_i)^\mathsf{T}(\mathbf{g}_i - \mathbf{X}_i\boldsymbol{\beta}_i)\;\middle|\;\mathbf{b}_\ell, \gamma_\ell\right] &= \mathbb{E}_Q\left[\gamma_{\ell,j}(\mathbf{g}_i - \mathbf{X}_i\boldsymbol{\beta}_{i\,|\,\gamma_{\ell,j}=1})^\mathsf{T}(\mathbf{g}_i - \mathbf{X}_i\boldsymbol{\beta}_{i\,|\,\gamma_{\ell,j}=1})\;\middle|\;\mathbf{b}_\ell, \gamma_\ell\right]\\ &= \gamma_{\ell,j}\mathbb{E}_Q\left[(\mathbf{g}_i - \mathbf{X}_i\boldsymbol{\beta}_{i\,|\,\gamma_{\ell,j}=1})^\mathsf{T}(\mathbf{g}_i - \mathbf{X}_i\boldsymbol{\beta}_{i\,|\,\gamma_{\ell,j}=1})\;\middle|\;\mathbf{b}_\ell, \gamma_\ell\right],\end{aligned}$$

since $\gamma_\ell$ is not integrated out in the expectation. Putting this together, it follows that $(*)$ may be written as follows, using the matrices defined above.

$$\begin{aligned}(*) &= \sum_{j=1}^{p}\gamma_{\ell,j}\left[\sum_{i=1}^{k} -\frac{1}{2\sigma_{e,i}^2}\left(\mathbb{E}_Q\left[\mathbf{r}_{\ell,i}^\mathsf{T}\mathbf{r}_{\ell,i}\right] - 2\mathbb{E}_Q\left[b_{\ell,i}\mathbf{r}_{\ell,i}^\mathsf{T}(\mathbf{X}_i)_j\;\middle|\;\mathbf{b}_\ell\right] + \mathbb{E}_Q\left[b_{\ell,i}^2(\mathbf{X}_i)_j^\mathsf{T}(\mathbf{X}_i)_j\;\middle|\;\mathbf{b}_\ell\right]\right)\right]\\
&= \sum_{j=1}^{p}\gamma_{\ell,j}\left[\sum_{i=1}^{k}\left(-\frac{1}{2\sigma_{e,i}^2}\mathbb{E}_Q\left[\mathbf{r}_{\ell,i}^\mathsf{T}\mathbf{r}_{\ell,i}\right] - \frac{1}{2\sigma_{e,i}^2}\left(-2\bar{\mathbf{r}}_{\ell,i}^\mathsf{T}(\mathbf{X}_i)_j b_{\ell,i} + b_{\ell,i}^2(\mathbf{X}_i)_j^\mathsf{T}(\mathbf{X}_i)_j\right)\right)\right]\\
&= \sum_{j=1}^{p}\gamma_{\ell,j}\left[\sum_{i=1}^{k} -\frac{1}{2\sigma_{e,i}^2}\mathbb{E}_Q\left[\mathbf{r}_{\ell,i}^\mathsf{T}\mathbf{r}_{\ell,i}\right]\right] + \sum_{j=1}^{p}\gamma_{\ell,j}\left[\underbrace{\sum_{i=1}^{k}\frac{1}{2\sigma_{e,i}^2}(2\bar{\mathbf{r}}_{\ell,i}^\mathsf{T}(\mathbf{X}_i)_j b_{\ell,i})}_{=\bar{\mathbf{r}}_\ell^\mathsf{T}\mathbf{Z}_j\mathbf{D}^{-1}\mathbf{b}_\ell} + \overbrace{\sum_{i'=1}^{k} -\frac{1}{2\sigma_{i',e}^2}b_{i',\ell}^2(\mathbf{X}_{i'})_j^\mathsf{T}(\mathbf{X}_{i'})_j}^{=-\frac{1}{2}\mathbf{b}_\ell^\mathsf{T}(\mathbf{Z}_j^\mathsf{T}\mathbf{Z}_j\mathbf{D}^{-1})\mathbf{b}_\ell}\right]\\
&= \sum_{j=1}^{p}\gamma_{\ell,j}\left[\sum_{i=1}^{k} -\frac{1}{2\sigma_{e,i}^2}\mathbb{E}_Q\left[\mathbf{r}_{\ell,i}^\mathsf{T}\mathbf{r}_{\ell,i}\right]\right] + \sum_{j=1}^{p}\gamma_{\ell,j}\left[\bar{\mathbf{r}}_\ell^\mathsf{T}\mathbf{Z}_j\mathbf{D}^{-1}\mathbf{b}_\ell - \frac{1}{2}\mathbf{b}_\ell^\mathsf{T}(\mathbf{Z}_j^\mathsf{T}\mathbf{Z}_j\mathbf{D}^{-1})\mathbf{b}_\ell\right].\end{aligned}$$

Notice the term $\sum_{j=1}^{p}\gamma_{\ell,j}\left[\sum_{i=1}^{k}\left(-\frac{1}{2\sigma_{e,i}^2}\mathbb{E}_Q\left[\mathbf{r}_{\ell,i}^\mathsf{T}\mathbf{r}_{\ell,i}\right]\right)\right]$ is independent of $j$, and so since $\sum_{j=1}^{p}\gamma_{\ell,j} \equiv 1$, we find

$$\sum_{j=1}^{p}\gamma_{\ell,j}\sum_{i=1}^{k} -\frac{1}{2\sigma_{e,i}^2}\mathbb{E}_Q[\mathbf{r}_{\ell,i}^\mathsf{T}\mathbf{r}_{\ell,i}] = \sum_{i=1}^{k} -\frac{1}{2\sigma_{e,i}^2}\mathbb{E}_Q[\mathbf{r}_{\ell,i}^\mathsf{T}\mathbf{r}_{\ell,i}],$$

which we may absorb into the constant term $O(1)$, since it is independent of the value of $\mathbf{b}_\ell$ and $\gamma_\ell$. Thus, we may write

$$\log Q(\mathbf{b}_\ell, \gamma_\ell) = \sum_{j=1}^{p}\gamma_{\ell,j}\left[-\frac{1}{2}\mathbf{b}_\ell^\mathsf{T}(\mathbf{Z}_j^\mathsf{T}\mathbf{Z}_j\mathbf{D}^{-1})\mathbf{b}_\ell + \bar{\mathbf{r}}_\ell^\mathsf{T}\mathbf{Z}_j\mathbf{D}^{-1}\mathbf{b}_\ell + \log\pi_j\right] - \frac{1}{2}\mathbf{b}_\ell^\mathsf{T}\mathbf{C}_\ell^{-1}\mathbf{b}_\ell + O(1)$$

Using the same trick $\sum_j \gamma_{\ell,j} \equiv 1$ again, we may write $-\frac{1}{2}\mathbf{b}_\ell^\mathsf{T}\mathbf{C}_\ell^{-1}\mathbf{b}_\ell = \sum_j -\frac{1}{2}\gamma_{\ell,j}\mathbf{b}_\ell^\mathsf{T}\mathbf{C}_\ell^{-1}\mathbf{b}_\ell$ and thereby write

$$\log Q(\mathbf{b}_\ell, \gamma_\ell) = \sum_{j=1}^{p}\gamma_{\ell,j}\left[-\frac{1}{2}\mathbf{b}_\ell^\mathsf{T}(\mathbf{Z}_j^\mathsf{T}\mathbf{Z}_j\mathbf{D}^{-1})\mathbf{b}_\ell + \bar{\mathbf{r}}_\ell^\mathsf{T}\mathbf{Z}_j\mathbf{D}^{-1}\mathbf{b}_\ell - \frac{1}{2}\mathbf{b}_\ell^\mathsf{T}\mathbf{C}_\ell^{-1}\mathbf{b}_\ell + \log\pi_j\right] + O(1).$$

We may now collect terms now as follows,

$$\bar{\mathbf{r}}_\ell^\mathsf{T}\mathbf{Z}_j\mathbf{D}^{-1}\mathbf{b}_\ell - \frac{1}{2}\mathbf{b}_\ell^\mathsf{T}(\mathbf{Z}_j^\mathsf{T}\mathbf{Z}_j\mathbf{D}^{-1})\mathbf{b}_\ell - \frac{1}{2}\mathbf{b}_\ell^\mathsf{T}\mathbf{C}_\ell^{-1}\mathbf{b}_\ell = \bar{\mathbf{r}}_\ell^\mathsf{T}\mathbf{Z}_j\mathbf{D}^{-1}\mathbf{b}_\ell - \frac{1}{2}\mathbf{b}_\ell^\mathsf{T}\underbrace{\left(\mathbf{Z}_j^\mathsf{T}\mathbf{Z}_j\mathbf{D}^{-1} + \mathbf{C}_\ell^{-1}\right)}_{\boldsymbol{\Sigma}_{\ell,j}^{-1}}\mathbf{b}_\ell$$

$$= \underbrace{\bar{\mathbf{r}}_\ell^\intercal \mathbf{Z}_j \mathbf{D}^{-1} \mathbf{b}_\ell \boldsymbol{\Sigma}_{\ell,j}}_{\boldsymbol{\mu}_{\ell,j}^\intercal} \boldsymbol{\Sigma}_{\ell,j}^{-1} \mathbf{b}_\ell - \frac{1}{2} \mathbf{b}_\ell^\intercal \boldsymbol{\Sigma}_{\ell,j}^{-1} \mathbf{b}_\ell = \boldsymbol{\mu}_{\ell,j}^\intercal \boldsymbol{\Sigma}_{\ell,j}^{-1} \mathbf{b}_\ell - \frac{1}{2} \mathbf{b}_\ell^\intercal \boldsymbol{\Sigma}_{\ell,j}^{-1} \mathbf{b}_\ell$$

$$= -\frac{1}{2} (\mathbf{b}_\ell - \boldsymbol{\mu}_{\ell,j})^\intercal \boldsymbol{\Sigma}_{\ell,j}^{-1} (\mathbf{b}_\ell - \boldsymbol{\mu}_{\ell,j}) + \frac{1}{2} \boldsymbol{\mu}_{\ell,j}^\intercal \boldsymbol{\Sigma}_{\ell,j}^{-1} \boldsymbol{\mu}_{\ell,j}.$$

In the last step, we have added and subtracted $-\frac{1}{2} \boldsymbol{\mu}_{\ell,j}^\intercal \boldsymbol{\Sigma}_{\ell,j}^{-1} \boldsymbol{\mu}_{\ell,j}$. Thus,

$$\log Q(\mathbf{b}_\ell, \gamma_\ell) = \sum_{j=1}^p \gamma_{\ell,j} \left[ -\frac{1}{2} (\mathbf{b}_\ell - \boldsymbol{\mu}_{\ell,j})^\intercal \boldsymbol{\Sigma}_{\ell,j}^{-1} (\mathbf{b}_\ell - \boldsymbol{\mu}_{\ell,j}) + \frac{1}{2} \boldsymbol{\mu}_{\ell,j}^\intercal \boldsymbol{\Sigma}_{\ell,j}^{-1} \boldsymbol{\mu}_{\ell,j} + \log \pi_j \right] + O(1).$$

If this is to be a distribution, it must integrate to 1 in $\mathbf{b}_\ell$ and $\gamma_\ell$. Exponentiating,

$$Q(\mathbf{b}_\ell, \gamma_\ell) = \tilde{C} \prod_{j=1}^p \exp\left( \gamma_{\ell,j} \left( -\frac{1}{2} (\mathbf{b}_\ell - \boldsymbol{\mu}_{\ell,j})^\intercal \boldsymbol{\Sigma}_{\ell,j}^{-1} (\mathbf{b}_\ell - \boldsymbol{\mu}_{\ell,j}) \right) \right) \exp\left( \gamma_{\ell,j} \left( \frac{1}{2} \boldsymbol{\mu}_{\ell,j}^\intercal \boldsymbol{\Sigma}_{\ell,j}^{-1} \boldsymbol{\mu}_{\ell,j} + \log \pi_j \right) \right),$$

for some normalizing constant $\tilde{C}$. Thus, integrating over $\mathbf{b}_\ell$ and $\gamma_\ell$, we must have the equality

$$1 = \tilde{C} \sum_{j=1}^p \exp\left( \log(\sqrt{(2\pi)^k \det \boldsymbol{\Sigma}_{\ell,j}}) + \frac{1}{2} \boldsymbol{\mu}_{\ell,j}^\intercal \boldsymbol{\Sigma}_{\ell,j}^{-1} \boldsymbol{\mu}_{\ell,j} + \log \pi_j \right).$$

Noticing that

$$\tilde{C} \sum_{j=1}^p \exp\left( \log(\sqrt{(2\pi)^k \det \boldsymbol{\Sigma}_{\ell,j}}) + \frac{1}{2} \boldsymbol{\mu}_{\ell,j}^\intercal \boldsymbol{\Sigma}_{\ell,j}^{-1} \boldsymbol{\mu}_{\ell,j} + \log \pi_j \right) = \tilde{C} \sum_{j=1}^p \exp\left( \log \pi_j - \log \mathcal{N}(\boldsymbol{\mu}_{\ell,j} \mid \mathbf{0}, \boldsymbol{\Sigma}_{\ell,j}) \right)$$

we find

$$\tilde{C} = \frac{1}{\sum_{j=1}^p \exp\left( \log \pi_j - \log \mathcal{N}(\boldsymbol{\mu}_{\ell,j} \mid \mathbf{0}, \boldsymbol{\Sigma}_{\ell,j}) \right)}.$$

Thus, we have found

$$Q(\mathbf{b}_\ell, \gamma_\ell) = \frac{\prod_{j=1}^p \exp\left( \gamma_{\ell,j} \left( -\frac{1}{2} (\mathbf{b}_\ell - \boldsymbol{\mu}_{\ell,j})^\intercal \boldsymbol{\Sigma}_{\ell,j}^{-1} (\mathbf{b}_\ell - \boldsymbol{\mu}_{\ell,j}) \right) \right) \exp\left( \gamma_{\ell,j} \left( \frac{1}{2} \boldsymbol{\mu}_{\ell,j}^\intercal \boldsymbol{\Sigma}_{\ell,j}^{-1} \boldsymbol{\mu}_{\ell,j} + \log \pi_j \right) \right)}{\sum_{j=1}^p \exp\left( \log \pi_j - \log \mathcal{N}(\boldsymbol{\mu}_{\ell,j} \mid \mathbf{0}, \boldsymbol{\Sigma}_{\ell,j}) \right)}.$$

which, multiplying by

$$1 \equiv \prod_{j=1}^p \left( \frac{\sqrt{(2\pi)^k \det \boldsymbol{\Sigma}_{\ell,j}}}{\sqrt{(2\pi)^k \det \boldsymbol{\Sigma}_{\ell,j}}} \right)^{\gamma_{\ell,j}}$$

we may group and rewrite

$$Q(\mathbf{b}_\ell, \gamma_\ell) = \prod_{j=1}^p \left( \frac{1}{\sqrt{(2\pi)^k \det \boldsymbol{\Sigma}_{\ell,j}}} \exp\left( \left( -\frac{1}{2} (\mathbf{b}_\ell - \boldsymbol{\mu}_{\ell,j})^\intercal \boldsymbol{\Sigma}_{\ell,j}^{-1} (\mathbf{b}_\ell - \boldsymbol{\mu}_{\ell,j}) \right) \right) \right)^{\gamma_{\ell,j}}$$
$$\times \prod_{j=1}^p \frac{\left( \exp\left( \log(\sqrt{(2\pi)^k \det \Sigma_{\ell,j}}) + \left( \frac{1}{2} \boldsymbol{\mu}_{\ell,j}^\intercal \boldsymbol{\Sigma}_{\ell,j}^{-1} \boldsymbol{\mu}_{\ell,j} + \log \pi_j \right) \right) \right)^{\gamma_{\ell,j}}}{\sum_{j=1}^p \exp\left( \log \pi_j - \mathcal{N}(\boldsymbol{\mu}_{\ell,j} \mid \mathbf{0}, \boldsymbol{\Sigma}_{\ell,j}) \right)},$$

which shows

$$Q(\mathbf{b}_\ell, \gamma_\ell) := \left( \prod_{j=1}^p \mathcal{N}(\mathbf{b}_\ell \mid \boldsymbol{\mu}_{\ell,j}, \boldsymbol{\Sigma}_{\ell,j})^{\gamma_{\ell,j}} \right) \mathrm{Multi}(\gamma_\ell \mid 1, \boldsymbol{\alpha}_\ell), \tag{4}$$

where $\boldsymbol{\alpha}_{\ell,j} = \left( \texttt{softmax}(\log \pi_i - \log \mathcal{N}(\boldsymbol{\mu}_{\ell,j} \mid \mathbf{0}, \boldsymbol{\Sigma}_{\ell,j})) \right)_j$. In particular,

$$Q(\mathbf{b}_\ell \mid \gamma_{\ell,j} = 1) := \mathcal{N}(\mathbf{b}_\ell \mid \boldsymbol{\mu}_{\ell,j}, \boldsymbol{\Sigma}_{\ell,j}) \quad \text{and} \quad Q(\gamma_\ell) := \mathrm{Multi}(1, \boldsymbol{\alpha}_\ell).$$

These distributions have the following interpretations under the SuShiE model. For the $\ell^{\text{th}}$ effect, $Q(\gamma_{\ell,j} = 1) = \boldsymbol{\alpha}_{\ell,j}$ is the posterior probability for SNP $j$ to be causal and $Q(\mathbf{b}_\ell \mid \gamma_{\ell,j} = 1)$ is the posterior distribution of the ancestry-specific effect sizes given that SNP $j$ is causal.

Moreover, under the mean-field hypothesis we have placed on $Q$, this computation shows

$$Q(\boldsymbol{\beta}) = \prod_{\ell=1}^L Q(\boldsymbol{\beta}_\ell) = \prod_{\ell=1}^L \left[ \left( \prod_{j=1}^p \mathcal{N}(\mathbf{b}_\ell \mid \boldsymbol{\mu}_{\ell,j}, \boldsymbol{\Sigma}_{\ell,j})^{\gamma_{\ell,j}} \right) \mathrm{Multi}(\gamma_\ell \mid 1, \boldsymbol{\alpha}_\ell) \right]. \tag{5}$$

This computation—of course—requires us to assume that $\mathbf{\Sigma}_{\ell,j}^{-1}$ (equivalently, $\mathbf{\Sigma}_{\ell,j}$) is symmetric and positive-definite. Since a sum of symmetric positive-definite matrices is once again symmetric positive-definite, it suffices to show that $\mathbf{Z}_j\mathbf{D}^{-1}\mathbf{Z}_j^\intercal$ and $\mathbf{C}_\ell^{-1}$ are symmetric positive definite. By assumption, $\mathbf{C}_\ell$ (hence, $\mathbf{C}_\ell^{-1}$) is symmetric positive definite. $\mathbf{Z}_j\mathbf{D}^{-1}\mathbf{Z}_j^\intercal$ is symmetric positive-definite due to positive-definiteness of $\mathbf{D}^{-1}$ and the fact that $\mathbf{Z}_j\mathbf{D}^{-1}\mathbf{Z}_j^\intercal$ can be re-written as a block-diagonal matrix of quadratic forms.

Here, we briefly present our algorithm for inferring variational parameters given genotype and molecular phenotype data, which is similar to the Iterative Bayesian Stepwise Selection algorithm introduced in Wang et al. [1].

---

**Algorithm 1** SuShiE

---

**Require:** Genotypic data $\mathbf{X}_{N_i\times P}$ and phenotypic data $\mathbf{y}_{N_i\times 1}$ for $i = 1,\ldots,k$ ancestries
**Require:** Number of single shared effects $L$
**Require:** Initialize the estimator $\overline{\mathbf{b}}_{\ell,i}$ for $\ell = 1,\cdots,L$ and $i = 1,\ldots,k$ for the posterior mean of $\mathbf{b}_{\ell,i}$
**Require:** Initialize hyperparameters $\mathbf{C}_\ell$, for $\ell = 1,\cdots,L$; $\sigma^2_{e,i}$ for $i = 1,\ldots,k$; $\boldsymbol{\pi} \in \mathbb{R}^{p\times 1}$
**Require:** Function to compute posterior distribution for variational parameters: $F$
**Require:** Function to update hyperparameter $\mathbf{C}$ and $\sigma^2_e$: $F_{\mathbf{C}}$ and $F_{\sigma^2_e}$
**Require:** Function to compute ELBO: $F_{\text{ELBO}}$
**Ensure:** ELBO increase

**repeat**
  **for** $i$ in $1,\cdots,k$ **do**
    $\bar{\mathbf{r}}_i = \mathbf{y}_i - \mathbf{X}_i \sum_{\ell=1}^{L} \overline{\mathbf{b}}_{\ell,i}$
  **end for**
  **for** $\ell$ in $1,\cdots,L$ **do**
    **for** $i$ in $1,\cdots,k$ **do**
      $\bar{\mathbf{r}}_{\ell,i} = \bar{\mathbf{r}}_i + \mathbf{X}\overline{\mathbf{b}}_{\ell,i}$
    **end for**
    $\mathbf{C}'_\ell \leftarrow F_{\mathbf{C}}(\bar{\mathbf{r}}_{\ell,i}, \mathbf{X}_i, \mathbf{C}_\ell, \sigma^2_{e,i}, \boldsymbol{\pi} \mid \text{for } i = 1,\ldots,k)$
    $(\boldsymbol{\alpha}_\ell, \boldsymbol{\mu}_\ell, \mathbf{\Sigma}_\ell) \leftarrow F(\bar{\mathbf{r}}_{\ell,i}, \mathbf{X}_i, \mathbf{C}'_\ell, \sigma^2_{e,i}, \boldsymbol{\pi} \mid \text{for } i = 1,\ldots,k)$
    **for** $i$ in $1,\cdots,k$ **do**
      $\overline{\mathbf{b}}_{\ell,i} \leftarrow \boldsymbol{\mu}_{\ell,i} \circ \boldsymbol{\alpha}_\ell$
      $\overline{\mathbf{b}}^2_{\ell,i} \leftarrow \text{diag}((\mathbf{d}_{\ell,i} + \boldsymbol{\mu}_{\ell,i} \circ \boldsymbol{\mu}_{\ell,i}) \circ \boldsymbol{\alpha}_\ell)$ where $\mathbf{d}_{\ell,i} = ((\mathbf{\Sigma}_{\ell,1})_{ii}, \ldots, (\mathbf{\Sigma}_{\ell,p})_{ii})$ and $p$ is SNP number
      $\bar{\mathbf{r}}_{\ell,i} = \bar{\mathbf{r}}_i - \mathbf{X}\overline{\mathbf{b}}_{\ell,i}$
    **end for**
  **end for**
  **for** $i$ in $1,\cdots,k$ **do**
    $\sigma^2_{e,i} \leftarrow F_{\sigma^2_e}(\mathbf{X}_i, \mathbf{y}_i, \overline{\mathbf{b}}_{\ell,i}, \overline{\mathbf{b}}^2_{\ell,i} \mid \text{for } \ell = 1,\cdots,L)$
  **end for**
  $ELBO \leftarrow F_{\text{ELBO}}$
**until** convergence criterion satisfied

---

See below for details on computing the ELBO as well as updating hyperparameters.

### 4. Computation of the ELBO

Here, we leverage the definition of approximating distributions $Q(\boldsymbol{\beta})$ and the assumptions of the SuShiE model to expand the ELBO. While evaluation of the ELBO is not necessary to perform variational inference under $Q$, it is useful for checking convergence, and serves as the objective for updating hyper-parameters. Recall that

$$\text{ELBO} = \mathbb{E}_Q\left[\log \frac{\Pr(\boldsymbol{\beta}, \mathbf{g} \mid \mathbf{X}, \mathbf{C}, \boldsymbol{\pi}, \boldsymbol{\sigma}^2_e)}{Q(\boldsymbol{\beta})}\right].$$

Using equation (1) and equation (2), we may compute

$$
\begin{aligned}
\mathsf{ELBO} &= \mathbb{E}_Q\left[\log \frac{\prod_{i=1}^{k} \mathcal{N}(\mathbf{g}_i \mid \mathbf{X}_i\boldsymbol{\beta}_i, \sigma_{e,i}^2\mathbf{I}) \Pr(\boldsymbol{\beta} \mid \mathbf{C}, \boldsymbol{\pi})}{Q(\boldsymbol{\beta})}\right] \\
&= \mathbb{E}_Q\left[\left(\sum_{i=1}^{k} \log \mathcal{N}(\mathbf{g}_i \mid \mathbf{X}_i\boldsymbol{\beta}_i, \sigma_{e,i}^2\mathbf{I})\right) + \log \frac{\Pr(\boldsymbol{\beta} \mid \mathbf{C}, \boldsymbol{\pi})}{Q(\boldsymbol{\beta})}\right] \\
&= \sum_{i=1}^{k} \mathbb{E}_Q\left[\log \mathcal{N}(\mathbf{g}_i \mid \mathbf{X}_i\boldsymbol{\beta}_i, \sigma_{e,i}^2\mathbf{I})\right] - \underbrace{\mathbb{E}_Q\left[\log \frac{Q(\boldsymbol{\beta})}{\Pr(\boldsymbol{\beta} \mid \mathbf{C}, \boldsymbol{\pi})}\right]}_{(*)} \qquad (6) \\
&= \left(\sum_{i=1}^{k} -\frac{n_i}{2}\log(2\pi\sigma_{e,i}^2) - \sum_{i=1}^{k} \frac{1}{2\sigma_{e,i}^2} \underbrace{\mathbb{E}_Q\left[(\mathbf{g}_i - \mathbf{X}_i\boldsymbol{\beta}_i)^{\mathsf{T}}(\mathbf{g}_i - \mathbf{X}_i\boldsymbol{\beta}_i)\right]}_{(**)}\right) - \underbrace{\mathbb{E}_Q\left[\log \frac{Q(\boldsymbol{\beta})}{\Pr(\boldsymbol{\beta} \mid \mathbf{C}, \boldsymbol{\pi})}\right]}_{(*)}.
\end{aligned}
$$

Evidently, only the expectations $\mathbb{E}_Q\left[(\mathbf{g}_i - \mathbf{X}_i\boldsymbol{\beta}_i)^{\mathsf{T}}(\mathbf{g}_i - \mathbf{X}_i\boldsymbol{\beta}_i)\right]$ and $\mathbb{E}_Q\left[\log \frac{Q(\boldsymbol{\beta})}{\Pr(\boldsymbol{\beta} \mid \mathbf{C}, \boldsymbol{\pi})}\right]$ remain to be computed, which we have labeled by $(**)$ and $(*)$, respectively. We will compute $(*)$ first and then $(**)$.

We may expand the term $(*)$ in equation (6). In order to do this, we will exploit equation (3) to integrate *first* over the discrete variables $\gamma_\ell$ wherever they appear.

(7)

$$
\begin{aligned}
(*) &= \mathbb{E}_Q\left[\log \frac{\prod_{\ell=1}^{L} Q(\mathbf{b}_\ell, \gamma_\ell)}{\prod_{\ell=1}^{L} \mathcal{N}(\mathbf{b}_\ell \mid \mathbf{0}, \mathbf{C})\mathrm{Multi}(\gamma_\ell \mid 1, \boldsymbol{\pi})}\right] = \mathbb{E}_Q\left[\log \frac{\prod_{\ell=1}^{L} Q(\mathbf{b}_\ell \mid \gamma_\ell)Q(\gamma_\ell)}{\prod_{\ell=1}^{L} \mathcal{N}(\mathbf{b}_\ell \mid \mathbf{0}, \mathbf{C})\mathrm{Multi}(\gamma_\ell \mid 1, \boldsymbol{\pi})}\right] \\
&= \sum_{\ell=1}^{L} \mathbb{E}_Q\left[\log \frac{Q(\mathbf{b}_\ell \mid \gamma_\ell)Q(\gamma_\ell)}{\mathcal{N}(\mathbf{b}_\ell \mid \mathbf{0}, \mathbf{C}_\ell)\mathrm{Multi}(\gamma_\ell \mid 1, \boldsymbol{\pi})}\right] = \sum_{\ell=1}^{L}\sum_{j=1}^{p} Q(\gamma_{\ell,j} = 1)\mathbb{E}_Q\left[\log \frac{Q(\mathbf{b}_\ell \mid \gamma_{\ell,j} = 1)Q(\gamma_{\ell,j} = 1)}{\mathcal{N}(\mathbf{b}_\ell \mid \mathbf{0}, \mathbf{C}_\ell)\mathrm{Multi}(\gamma_{\ell,j} = 1 \mid \boldsymbol{\pi})}\right] \\
&= \sum_{\ell=1}^{L}\sum_{j=1}^{p} Q(\gamma_{\ell,j} = 1)\mathbb{E}_Q\left[\log \frac{Q(\mathbf{b}_\ell \mid \gamma_{\ell,j} = 1)}{\mathcal{N}(\mathbf{b}_\ell \mid \mathbf{0}, \mathbf{C}_\ell)}\right] + \sum_{\ell=1}^{L}\sum_{j=1}^{p} Q(\gamma_{\ell,j} = 1)\left[\log \frac{Q(\gamma_{\ell,j} = 1)}{\mathrm{Multi}(\gamma_{\ell,j} = 1 \mid \boldsymbol{\pi})}\right] \\
&= \sum_{\ell=1}^{L}\sum_{j=1}^{p} Q(\gamma_{\ell,j} = 1)D_{\mathsf{KL}}\left[Q(\mathbf{b}_\ell \mid \gamma_{\ell,j} = 1)\middle\|\mathcal{N}(\mathbf{b}_\ell \mid \mathbf{0}, \mathbf{C}_\ell)\right] + \sum_{\ell=1}^{L} D_{\mathsf{KL}}\left[Q(\gamma_\ell)\|\Pr(\gamma_\ell)\right].
\end{aligned}
$$

The term $D_{\mathsf{KL}}\left[Q(\mathbf{b}_\ell \mid \gamma_{\ell,j} = 1)\middle\|\mathcal{N}(\mathbf{b}_\ell \mid \mathbf{0}, \mathbf{C}_\ell)\right]$ is the Kullback-Leibler divergence between two Guassians, and therefore has the form

$$
D_{\mathsf{KL}}\left[Q(\mathbf{b}_\ell \mid \gamma_{\ell,j} = 1)\middle\|\mathcal{N}(\mathbf{b}_\ell \mid \mathbf{0}, \mathbf{C}_\ell)\right] = \frac{1}{2}\left(\mathrm{tr}(\mathbf{C}_\ell^{-1}\boldsymbol{\Sigma}_{\ell,j}) - k + \boldsymbol{\mu}_{\ell,j}^{\mathsf{T}}\mathbf{C}_\ell^{-1}\boldsymbol{\mu}_{\ell,j} + \log\det\mathbf{C}_\ell - \log\det\boldsymbol{\Sigma}_{\ell,j}\right).
$$

Similarly, the term $D_{\mathsf{KL}}\left[Q(\gamma_\ell)\|\Pr(\gamma_\ell)\right]$ is the Kullback-Leibler divergence between two discrete distributions over $p$ states, and takes the form $\sum_{j=1}^{p} \boldsymbol{\alpha}_{\ell,j}\log\left(\boldsymbol{\alpha}_{\ell,j}/\boldsymbol{\pi}_j\right)$.

It now only remains to compute $(**)$ in equation (6). Before proceeding, we will need to collect the first and second moment of the posterior distribution for $\boldsymbol{\beta}_{\ell,i} = b_{\ell,i}\gamma_\ell$ (the posterior distribution of the $\ell^{\text{th}}$ effect for ancestry $i$). For this, it is easier to write $Q(\boldsymbol{\beta}_{\ell,i}) = Q(b_{\ell,i}, \gamma_\ell) = Q(b_{\ell,i} \mid \gamma_\ell)Q(\gamma_\ell)$ and integrate over $\gamma_\ell$ first.

$$
\begin{aligned}
\mathbb{E}_Q\left[\boldsymbol{\beta}_{\ell,i}\right] &= \mathbb{E}_Q\left[b_{\ell,i}\gamma_\ell\right] = \boldsymbol{\mu}_{\ell,i} \circ \boldsymbol{\alpha}_\ell \\
\mathbb{E}_Q\left[\boldsymbol{\beta}_{\ell,i}\boldsymbol{\beta}_{\ell,i}^{\mathsf{T}}\right] &= \mathbb{E}_Q\left[b_{\ell,i}^2\gamma_\ell\gamma_\ell^{\mathsf{T}}\right] = \mathrm{diag}((\mathbf{d}_{\ell,i} + \boldsymbol{\mu}_{\ell,i} \circ \boldsymbol{\mu}_{\ell,i}) \circ \boldsymbol{\alpha}_\ell)
\end{aligned} \qquad (8)
$$

where $\boldsymbol{\mu}_{\ell,i} = [(\boldsymbol{\mu}_{\ell,1})_i, \ldots, (\boldsymbol{\mu}_{\ell,p})_i]$ is the $p \times 1$ vector of re-arranged posterior effect-size expectations for each SNP in ancestry $i$, $\boldsymbol{\alpha}_\ell$ is the $p \times 1$ vector whose $j^{\text{th}}$ entry is the posterior probability SNP $j$ is causal for effect $\ell$. Recall that $\boldsymbol{\Sigma}_{\ell,j}$ is a $k \times k$ matrix for SNP $j$ at the $\ell$ effect for $k$ ancestries. Here, $\mathbf{d}_{\ell,i}$ in equation (8) is a $p$ vector whose $j^{\text{th}}$ entry is the $i^{\text{th}}$ diagonal entry in $\boldsymbol{\Sigma}_{\ell,j}$ across all $1 \leq j \leq p$ SNPs. In other words,

$$
\mathbf{d}_{\ell,i} = ((\boldsymbol{\Sigma}_{\ell,1})_{ii}, \ldots, (\boldsymbol{\Sigma}_{\ell,p})_{ii}).
$$

Notice that $\boldsymbol{\beta}_i^\mathsf{T}\mathbf{X}_i^\mathsf{T}\mathbf{X}_i\boldsymbol{\beta}_i$ is a scalar so that $\boldsymbol{\beta}_i^\mathsf{T}\mathbf{X}_i^\mathsf{T}\mathbf{X}_i\boldsymbol{\beta}_i = \operatorname{tr}\left[\boldsymbol{\beta}_i^\mathsf{T}\mathbf{X}_i^\mathsf{T}\mathbf{X}_i\boldsymbol{\beta}_i\right]$ and, thus, with the cyclic property of trace, linearity of expectation, we have the following computation.

$$\begin{aligned}
&\mathbb{E}_Q\left[(\mathbf{g}_i - \mathbf{X}_i\boldsymbol{\beta}_i)^\mathsf{T}(\mathbf{g}_i - \mathbf{X}_i\boldsymbol{\beta}_i)\right] = \mathbb{E}_Q\left[\mathbf{g}_i^\mathsf{T}\mathbf{g}_i - 2\mathbf{g}_i^\mathsf{T}\mathbf{X}_i\boldsymbol{\beta}_i + (\mathbf{X}_i\boldsymbol{\beta}_i)^\mathsf{T}\mathbf{X}_i\boldsymbol{\beta}_i\right]\\
&= \mathbf{g}_i^\mathsf{T}\mathbf{g}_i - 2\mathbf{g}_i^\mathsf{T}\mathbf{X}_i\mathbb{E}_Q\left[\boldsymbol{\beta}_i\right] + \mathbb{E}_Q\left[\boldsymbol{\beta}_i^\mathsf{T}\mathbf{X}_i^\mathsf{T}\mathbf{X}_i\boldsymbol{\beta}_i\right]\\
&= \mathbf{g}_i^\mathsf{T}\mathbf{g}_i - 2\mathbf{g}_i^\mathsf{T}\mathbf{X}_i\mathbb{E}_Q[\boldsymbol{\beta}_i] + \mathbb{E}_Q\left[\operatorname{tr}\left[\mathbf{X}_i^\mathsf{T}\mathbf{X}_i\boldsymbol{\beta}_i\boldsymbol{\beta}_i^\mathsf{T}\right]\right]\\
&= \mathbf{g}_i^\mathsf{T}\mathbf{g}_i - 2\mathbf{g}_i^\mathsf{T}\mathbf{X}_i\mathbb{E}_Q\left[\boldsymbol{\beta}_i\right] + \operatorname{tr}\left[\mathbf{X}_i^\mathsf{T}\mathbf{X}_i\mathbb{V}_Q\left[\boldsymbol{\beta}_i\right]\right] + \mathbb{E}_Q\left[\boldsymbol{\beta}_i\right]^\mathsf{T}\mathbf{X}_i^\mathsf{T}\mathbf{X}_i\mathbb{E}_Q\left[\boldsymbol{\beta}_i\right]\\
&= (\mathbf{g}_i - \mathbf{X}_i\mathbb{E}_Q\left[\boldsymbol{\beta}_i\right])^\mathsf{T}(\mathbf{g}_i - \mathbf{X}_i\mathbb{E}_Q\left[\boldsymbol{\beta}_i\right]) + \operatorname{tr}\left[\mathbf{X}_i^\mathsf{T}\mathbf{X}_i\mathbb{V}_Q\left[\boldsymbol{\beta}_i\right]\right]\\
&= (\mathbf{g}_i - \mathbf{X}_i\mathbb{E}_Q\left[\boldsymbol{\beta}_i\right])^\mathsf{T}(\mathbf{g}_i - \mathbf{X}_i\mathbb{E}_Q\left[\boldsymbol{\beta}_i\right]) + \operatorname{tr}\left[\mathbf{X}_i^\mathsf{T}\mathbf{X}_i\mathbb{V}_Q\left[\sum_{\ell=1}^{L}\boldsymbol{\beta}_{\ell,i}\right]\right]\\
&= (\mathbf{g}_i - \mathbf{X}_i\mathbb{E}_Q\left[\boldsymbol{\beta}_i\right])^\mathsf{T}(\mathbf{g}_i - \mathbf{X}_i\mathbb{E}_Q\left[\boldsymbol{\beta}_i\right]) + \sum_{\ell=1}^{L}\operatorname{tr}\left[\mathbf{X}_i^\mathsf{T}\mathbf{X}_i\mathbb{V}_Q\left[\boldsymbol{\beta}_{\ell,i}\right]\right]\\
&= (\mathbf{g}_i - \mathbf{X}_i\mathbb{E}_Q\left[\boldsymbol{\beta}_i\right])^\mathsf{T}(\mathbf{g}_i - \mathbf{X}_i\mathbb{E}_Q\left[\boldsymbol{\beta}_i\right]) + \sum_{\ell=1}^{L}\operatorname{tr}\left[\mathbf{X}_i^\mathsf{T}\mathbf{X}_i\left(\mathbb{E}_Q\left[\boldsymbol{\beta}_{\ell,i}\boldsymbol{\beta}_{\ell,i}^\mathsf{T}\right] - \mathbb{E}_Q\left[\boldsymbol{\beta}_{\ell,i}\right]\mathbb{E}_Q[\boldsymbol{\beta}_{\ell,i}]^\mathsf{T}\right)\right]\\
&= (\mathbf{g}_i - \mathbf{X}_i\mathbb{E}_Q\left[\boldsymbol{\beta}_i\right])^\mathsf{T}(\mathbf{g}_i - \mathbf{X}_i\mathbb{E}_Q\left[\boldsymbol{\beta}_i\right]) + \sum_{\ell=1}^{L}\operatorname{tr}\left[\mathbf{X}_i^\mathsf{T}\mathbb{E}_Q[\boldsymbol{\beta}_{\ell,i}\boldsymbol{\beta}_{\ell,i}^\mathsf{T}]\mathbf{X}_i\right] - \sum_{\ell=1}^{L}\mathbb{E}_Q[\boldsymbol{\beta}_{\ell,i}]^\mathsf{T}\mathbf{X}_i^\mathsf{T}\mathbf{X}_i\mathbb{E}_Q[\boldsymbol{\beta}_{\ell,i}].
\end{aligned} \tag{9}$$

We note that this is the expectation of a residual sum of squares (RSS), where Wang et al. briefly derived this in their original paper. We denote this as $\mathbb{E}_Q[\text{RSS}]_i$ for ancestry $i$. In particular, note that $\mathbb{E}_Q[\text{RSS}]_i \geq 0$ for all $i$ [1].

The full ELBO may now be written as a function of the $\boldsymbol{\mu}_{\ell,j}$ and $\boldsymbol{\Sigma}_{\ell,j}$ as follows.

$$\begin{aligned}
\mathsf{ELBO}(\boldsymbol{\mu}, \boldsymbol{\Sigma}) &= \sum_{i=1}^{k} -\frac{n_i}{2}\log(2\pi\sigma_{e,i}^2) - \frac{1}{2\sigma_{e,i}^2}\left(\mathbf{g}_i - \sum_{\ell=1}^{L}\mathbf{X}_i(\boldsymbol{\mu}_{\ell,i}\circ\boldsymbol{\alpha}_\ell)\right)^\mathsf{T}\left(\mathbf{g}_i - \sum_{\ell=1}^{L}\mathbf{X}_i(\boldsymbol{\mu}_{\ell,i}\circ\boldsymbol{\alpha}_\ell)\right)\\
&+ \sum_{i=1}^{k}\sum_{j=1}^{p}(\mathbf{X}_i)_j^\mathsf{T}(\mathbf{X}_i)_j\left(\sum_{\ell=1}^{L}\operatorname{diag}((\mathbf{d}_{\ell,i} + \boldsymbol{\mu}_{\ell,i}\circ\boldsymbol{\mu}_{\ell,i})\circ\boldsymbol{\alpha}_\ell)\right)_j\\
&- \sum_{i=1}^{k}\sum_{\ell=1}^{L}(\boldsymbol{\mu}_{\ell,i}\circ\boldsymbol{\alpha}_\ell)^\mathsf{T}\mathbf{X}_i^\mathsf{T}\mathbf{X}_i(\boldsymbol{\mu}_{\ell,i}\circ\boldsymbol{\alpha}_\ell)\\
&- \sum_{\ell=1}^{L}\sum_{j=1}^{p}\frac{\boldsymbol{\alpha}_{\ell,j}}{2}\left(\operatorname{tr}(\mathbf{C}_\ell^{-1}\boldsymbol{\Sigma}_{\ell,j}) - k + \boldsymbol{\mu}_{\ell,j}^\mathsf{T}\mathbf{C}_\ell^{-1}\boldsymbol{\mu}_{\ell,j} + \log\det\mathbf{C}_\ell - \log\det\boldsymbol{\Sigma}_{\ell,j} + 2\log\left(\frac{\boldsymbol{\alpha}_{\ell,j}}{\boldsymbol{\pi}_j}\right)\right).
\end{aligned} \tag{10}$$

### 5. ESTIMATING HYPERPARAMETERS

We may use empirical Bayes to update our hyperparameters $\sigma_{e,i}^2$ ($i = 1, \ldots, k$) and $\mathbf{C}_\ell$ ($\ell = 1, \ldots, L$) by finding the corresponding estimation that maximizes the ELBO.

5.1. **Updating environmental/residual variance.** Let us now consider $\sigma_{e,i}^2$ to be a variable so that the ELBO is a function of the $\boldsymbol{\mu}_{\ell,i}$, $\boldsymbol{\Sigma}_{\ell,i}$ and $\sigma_{e,i}^2$. We will maximize the ELBO with respect to $\sigma_{e,i}^2$ while keeping all other parameters fixed.

From equations (6) and (9), we may compute

$$\begin{aligned}
\frac{\partial\,\mathsf{ELBO}}{\partial\,\sigma_{e,i}^2} &= \frac{\partial}{\partial\sigma_{e,i}^2}\mathbb{E}_Q\left[\log\mathcal{N}(\mathbf{g}_i - \mathbf{X}_i\boldsymbol{\beta}_i \mid \mathbf{0}, \sigma_{e,i}^2\mathbf{I}_{n_i})\right]\\
&= \frac{\partial}{\partial\sigma_{e,i}^2}\left[-\frac{n_i}{2}\log(2\pi\sigma_{e,i}^2) - \frac{1}{2\sigma_{e,i}^2}\mathbb{E}[\text{RSS}]_i\right]\\
&= -\frac{n_i}{2\sigma_{e,i}^2} + \frac{1}{2(\sigma_{e,i}^2)^2}\mathbb{E}[\text{RSS}]_i \Rightarrow\\
\widehat{\sigma}_{i,e}^2 &= \frac{\mathbb{E}[\text{RSS}]_i}{n_i}.
\end{aligned} \tag{11}$$

It is prudent to show that this actually maximizes the ELBO when the $\boldsymbol{\mu}_{\ell,i}$ and $\boldsymbol{\Sigma}_{\ell,i}$ are held fixed. For this, one simply observes that for values $\sigma_{e,i}^2 < \widehat{\sigma}_{i,e}^2$, $\dfrac{\partial\,\mathsf{ELBO}}{\partial\,\sigma_{e,i}^2} > 0$, while for values $\sigma_{e,i}^2 > \widehat{\sigma}_{i,e}^2$, $\dfrac{\partial\,\mathsf{ELBO}}{\partial\,\sigma_{e,i}^2} < 0$.

5.2. **Updating prior for effect size covariance matrix.** We will infer the prior effect covariance $\mathbf{C}_\ell$ by considering the ELBO to also be a function of the symmetric positive-definite matrix $\mathbf{C}_\ell$—similar to above—and maximizing the ELBO in $\mathbf{C}_\ell$. Recall that the set of symmetric positive-definite matrices form an (open) convex subspace of $\mathbb{R}^{\binom{k+1}{2}} = \mathbb{R}^{k(k+1)/2}$. However, since the ELBO will not necessarily be concave in $\mathbf{C}_\ell$, we will need to take a different approach to this optimization problem than we used before. We will be able to directly prove the existence and uniqueness of the point $\widehat{\mathbf{C}}_\ell$ at which the ELBO attains its maximum as a function $\mathbf{C}_\ell$.

To begin, we will extract the part of the ELBO that depends on $\mathbf{C}_\ell$. From equations (6, 7), the only place the ELBO depends on the parameter $\mathbf{C}_\ell$ is in term

$$-\sum_{j=1}^{p} Q(\gamma_{\ell,j}=1) D_{\mathsf{KL}}\left[Q(\mathbf{b}_\ell \mid \gamma_{\ell,j}=1) \middle\| \mathcal{N}(\mathbf{b}_\ell \mid \mathbf{0}, \mathbf{C}_\ell)\right].$$

Thus, to maximize the ELBO, we need only maximize this term or, equivalently, *minimize* its negative

$$\sum_{j=1}^{p} Q(\gamma_{\ell,j}=1) D_{\mathsf{KL}}\left[Q(\mathbf{b}_\ell \mid \gamma_{\ell,j}=1) \middle\| \mathcal{N}(\mathbf{b}_\ell \mid \mathbf{0}, \mathbf{C}_\ell)\right],$$

which we will now focus on. From equation (10), we observe this may be written as

$$\sum_{j=1}^{p} \frac{Q(\gamma_{\ell,j}=1)}{2}\left(\mathrm{tr}(\mathbf{C}_\ell^{-1}\boldsymbol{\Sigma}_{\ell,j}) - k + \boldsymbol{\mu}_{\ell,j}^{\mathsf{T}}\mathbf{C}_\ell^{-1}\boldsymbol{\mu}_{\ell,j} + \log\det\mathbf{C}_\ell - \log\det\boldsymbol{\Sigma}_{\ell,j}\right).$$

Once again extracting the terms which depend on $\mathbf{C}_\ell$, we are left with

$$\sum_{j=1}^{p} \frac{Q(\gamma_{\ell,j}=1)}{2}\left(\mathrm{tr}(\mathbf{C}_\ell^{-1}\boldsymbol{\Sigma}_{\ell,j}) + \boldsymbol{\mu}_{\ell,j}^{\mathsf{T}}\mathbf{C}_\ell^{-1}\boldsymbol{\mu}_{\ell,j} + \log\det\mathbf{C}_\ell\right),$$

which, since $\sum_{j=1}^{p} \frac{Q(\gamma_{\ell,j}=1)}{2} = \frac{1}{2}$, we may write as

$$\frac{1}{2}\log\det\mathbf{C}_\ell + \sum_{j=1}^{p} \frac{Q(\gamma_{\ell,j}=1)}{2}\left(\mathrm{tr}(\mathbf{C}_\ell^{-1}\boldsymbol{\Sigma}_{\ell,j}) + \boldsymbol{\mu}_{\ell,j}^{\mathsf{T}}\mathbf{C}_\ell^{-1}\boldsymbol{\mu}_{\ell,j}\right).$$

By properties of the trace, we may collect terms in the sum to form the quantity we wish to minimize

$$\frac{1}{2}\left(\log\det\mathbf{C}_\ell + \mathrm{tr}(\mathbf{C}_\ell^{-1}\widehat{\mathbf{C}}_\ell)\right), \tag{12}$$

where

$$\widehat{\mathbf{C}}_\ell = \sum_{j=1}^{p} Q(\gamma_{\ell,j}=1)(\boldsymbol{\Sigma}_{\ell,j} + \boldsymbol{\mu}_{\ell,j}\boldsymbol{\mu}_{\ell,j}^{\mathsf{T}}).$$

We claim that $\widehat{\mathbf{C}}_\ell$ is a symmetric positive-definite matrix and so is, in addition, invertible. $\widehat{\mathbf{C}}_\ell$ is symmetric as a linear combination of symmetric matrices; it is positive-definite because $\boldsymbol{\Sigma}_{\ell,j}$ is positive-definite and the matrices $\boldsymbol{\mu}_{\ell,j}\boldsymbol{\mu}_{\ell,j}^{\mathsf{T}}$ are positive semidefinite, so that $\widehat{\mathbf{C}}_\ell$ is, in particular, a convex combination of the positive-definite matrices $\boldsymbol{\Sigma}_{\ell,j} + \boldsymbol{\mu}_{\ell,j}\boldsymbol{\mu}_{\ell,j}^{\mathsf{T}}$.

We may now recognize a KL-divergence hiding in equation (12). There is an equality

$$\frac{1}{2}\left(\log\det\mathbf{C}_\ell + \mathrm{tr}(\mathbf{C}_\ell^{-1}\widehat{\mathbf{C}}_\ell)\right) = D_{\mathsf{KL}}\left[\mathcal{N}(\mathbf{0}, \mathbf{C}_\ell) \middle\| \mathcal{N}(\mathbf{0}, \widehat{\mathbf{C}}_\ell)\right] + \frac{1}{2}\left(k + \log\det\widehat{\mathbf{C}}_\ell\right),$$

and right-hand side of this equation only depends on $\mathbf{C}_\ell$ through the KL-divergence. It follows that equation (12) attains its unique minimum precisely at the point $\mathbf{C}_\ell = \widehat{\mathbf{C}}_\ell$. This implies that the ELBO, as a function of $\mathbf{C}_\ell$ alone, attains its unique *maximum* precisely at the point $\mathbf{C}_\ell = \widehat{\mathbf{C}}_\ell$.
